# Artificial Intelligence for Cardiac Biomarkers After Myocardial Infarction: A Systematic Review and a Leakage-Aware Modeling Framework

**DOI:** 10.64898/2026.04.28.26351958

**Authors:** Natalia Piórkowska, Agnieszka Olejnik, Lech Madeyski, Aleksandra Musz, Wiktor Kuliczkowski, Andrzej Mysiak, Aleksandra Żyłka, Iwona Bil-Lula

**Affiliations:** Faculty of Information and Communication Technology, Wroclaw University of Science and Technology, Wrocław, Poland; Department of Medical Laboratory Diagnostics, Division of Clinical Chemistry and Laboratory Haematology, Faculty of Pharmacy, Wroclaw Medical University, Wroclaw, Poland; Student Scientific Association at the Cardiac Perfusion Laboratory, Department of Medical Laboratory Diagnostics, Division of Clinical Chemistry and Laboratory Haematology, Faculty of Pharmacy, Wroclaw Medical University, Wroclaw, Poland; Institute of Heart Diseases, Wroclaw Medical University, Wroclaw, Poland; Independent Researcher; Specialized Immunology Laboratory, Clinical Department of Nephrology, Transplantation Medicine and Internal Diseases, Institute of Internal Diseases, Wroclaw Medical University, Poland

**Keywords:** myocardial infarction, cardiac biomarkers, artificial intelligence, machine learning, risk stratification, prediction modeling

## Abstract

**Aims:** To systematically evaluate how artificial intelligence and machine-learning (AI/ML) methods are applied to cardiac biomarkers after myocardial infarction (MI), identify recurring methodological limitations, and operationalize a leakage-aware modelling workflow in a proof-of-concept post-MI dataset using a controlled proxy classification task.

**Methods and results:** A PRISMA 2020-compliant systematic review of studies published between 2015 and 2025 identified 120 eligible studies from 1,389 records. Most studies used multimodal inputs combining biomarkers with clinical or functional variables (109/120, 90.8%) and focused on prediction or prognostic modelling (89/120, 74.2%). Logistic or regularized regression (76/120, 63.3%) and Random Forest (69/120, 57.5%) were the most frequently used approaches. Internal validation predominated, whereas independent external validation was reported in only 44/120 studies (36.7%). Area under the receiver operating characteristic curve (ROC-AUC) was reported in 114/120 studies (95.0%), while calibration analyses and decision-curve analysis remained limited. Formal explainability methods were used inconsistently, and public code availability was uncommon.

To translate these observations into a practical framework, we implemented a leakage-aware machine-learning workflow in a proof-of-concept dataset of 152 patients with MI and 117 variables. The analytical task was defined as a binary classification problem (STEMI vs NSTEMI), used intentionally as a methodological proxy rather than a clinically relevant prognostic endpoint. Three predefined feature-set variants were benchmarked using nested cross-validation. The FULL variant achieved near-perfect discrimination [ROC-AUC 0.9988 (95% CI 0.9925-1.000)], the CLINICAL variant showed modest performance [0.6025 (0.4463-0.7450)], and the BIOMARKERS variant yielded strong discrimination with low dimensionality [0.9300 (0.8537-0.9863)]. Permutation-based falsification testing reduced performance towards chance level, supporting the procedural integrity of the workflow.

**Conclusions:** AI/ML research on cardiac biomarkers after MI is expanding rapidly but remains limited by heterogeneous methodology, insufficient external validation, incomplete interpretability, and weak reproducibility practices. A leakage-aware framework integrating explicit feature governance, nested validation, calibration assessment, robustness analyses, and falsification testing may improve the credibility and translational relevance of biomarker-based cardiovascular AI studies. However, the proof-of-concept case study is intended as a methodological demonstration and does not represent prognostic modelling of post-MI outcomes.

**Translational Perspective:** AI models using cardiac biomarkers after MI often report strong discrimination, but their clinical value is undermined by limited external validation, incomplete calibration assessment, and poor transparency. Our systematic review identifies these recurrent weaknesses, while the proof-of-concept case study demonstrates how a leakage-aware workflow can distinguish clinically plausible signals from structurally inflated performance under controlled analytical conditions. The use of a proxy classification task highlights methodological behavior rather than clinical prognosis, underscoring the need for future studies to validate such frameworks on clinically meaningful post-MI outcomes. Integrating explicit feature governance, nested validation, calibration, decision-analytic assessment, and falsification testing may help move biomarker-based cardiovascular AI from promising retrospective performance towards more reproducible and clinically trustworthy prediction models.

## 1. Introduction

Cardiovascular diseases remain the leading cause of morbidity and mortality worldwide^1^, with myocardial infarction (MI) representing one of the most critical events in this spectrum^2,3^. Precise and prompt diagnosis plays a crucial role in improving patient survival after MI. Current diagnostic methods combine assessment of clinical symptoms, electrocardiographic (ECG) findings, and measurement of circulating cardiac biomarkers. In cases of ST-segment elevation MI (STEMI), the presence of ST-segment elevation on ECG typically leads to immediate medical intervention. In contrast, diagnosing non-ST segment elevation MI (NSTEMI) often relies on changes in biomarker levels, particularly in patients with atypical clinical manifestations^4^. Among these biomarkers, cardiac troponin I (cTnI) and T (cTnT) are regarded as the gold standard because of their superior sensitivity and specificity. Nevertheless, none of the currently used indicators perfectly fulfills the requirements for an ideal biomarker that ensures both early and accurate MI detection^4^. Therefore, discovering novel biomarkers may help minimize diagnostic delays and enable the prompt implementation of appropriate therapeutic strategies. Despite substantial advances in acute management, risk stratification and long-term prognosis after MI remain challenging due to the multifactorial nature of post-infarction remodeling, inflammation, and comorbidities^3,5^. Accurate prediction of adverse outcomes - such as heart failure, arrhythmias, or recurrent ischemic events—relies on the integration of diverse clinical, biochemical, and functional biomarkers^6,7^. However, conventional statistical approaches often fail to capture complex nonlinear interactions among these markers, limiting their predictive power in individualized risk assessment^8–12^.

In recent years, artificial intelligence (AI) and machine learning (ML) have emerged as transformative tools in cardiovascular medicine, enabling the development of models capable of learning intricate patterns from multidimensional datasets^9,13–15^. These approaches have shown promise in predicting mortality^14,16^, left ventricular dysfunction^17,18^, and rehospitalization after MI^19,20^ by integrating heterogeneous data types - from serum biomarkers and ECG features to clinical parameters. Compared with traditional regression models, AI/ML methods such as random forests, support vector machines (SVM), and deep learning (DL) architectures can uncover latent relationships and interactions not apparent through linear modeling^9,10,12,21^.

The growing application of AI/ML to cardiac biomarkers reflects a paradigm shift from descriptive analyses towards predictive and precision medicine^14,22^. Recent advancements in biomarker research underscore the promising role of microRNAs, heart-type fatty acid-binding protein, and troponins measurable in urine or saliva as emerging tools for the early detection of MI^4^. Notably, a metabolomic analysis identified a metabolite profile that may have potential in the differentiation of STEMI and NSTEMI^23^. Biochemical markers such as high-sensitivity troponins, NT-proBNP, C-reactive protein (CRP), fibroblast growth factor 23 (FGF23), and galectin-3 - combined with functional and clinical data - have been used to develop models for post-MI risk stratification^24^. Yet, despite encouraging results, methodological heterogeneity across studies has led to inconsistent findings and limited reproducibility^9,15,25^. Key elements such as feature preprocessing, validation strategy, interpretability, and reporting standards vary substantially, hindering cross-study comparability and clinical translation^15,26,27^.

Emerging evidence suggests that methodological heterogeneity - rather than data scarcity - has become a key barrier to progress in AI-driven cardiology^13,14,28^. Studies often differ in their data preprocessing, feature selection (e.g., recursive feature elimination, Boruta)^29^, and validation design (e.g., k-fold versus external validation)^30,31^. Moreover, reporting transparency remains suboptimal: only a minority of studies share their code or raw data, and external validation in independent cohorts is rarely performed^9,26,32^. This raises concerns regarding data leakage, overfitting, and limited generalizability^9,33,34^. Importantly, model interpretability—a prerequisite for clinical implementation - remains underreported^15^, despite the availability of explainability tools such as SHAP (SHapley Additive exPlanations) and permutation importance^35^.

While previous reviews have summarized applications of AI in cardiovascular research, few have provided a systematic and critical assessment focused specifically on the use of cardiac biomarkers in post-MI patients. Furthermore, methodological rigor has rarely been evaluated using established frameworks such as CHARMS, PROBAST, and TRIPOD-ML/AI. The absence of standardized methodological evaluation limits understanding of how AI/ML models are developed, validated, and reported, and prevents the identification of consistent best practices.

To address these gaps, we conducted a systematic review of studies applying AI/ML methods to cardiac biomarkers in patients after MI, covering studies published between 2015 and 2025. The review was pre-registered in PROSPERO to ensure transparency and reproducibility and followed the PRISMA 2020 reporting guidelines. The overarching goal was to provide a comprehensive synthesis of methodological trends, recurring limitations, and best practice recommendations for AI/ML modeling in post-MI prognosis.

Specifically, this review aimed to:

i. characterize the types of MI biomarkers (clinical, biochemical, functional) most frequently used in AI/ML models (RQ1);
ii. identify the most commonly applied AI/ML algorithms and evaluate their methodological quality (RQ2);
iii. assess validation strategies and their adequacy for clinical prediction (RQ3);
iv. summarize model performance metrics and evaluate their clinical relevance (RQ4);
v. examine model interpretability and its contribution to clinical decision-making insight (RQ5);
vi. identify recurring reporting limitations, including data and code availability (RQ6); and
vii. propose methodological recommendations for improving reproducibility, robustness, and clinical utility (RQ7).

Through this approach, we aim to map the current methodological landscape and establish a foundation for evidence-based recommendations to guide the design, validation, and reporting of AI/ML studies using cardiac biomarkers of MI. The findings are expected to inform both the cardiovascular research community and clinicians adopting AI-driven tools in clinical practice, promoting transparent, interpretable, and clinically meaningful model development.

## 2. Methods

This systematic review was conducted in accordance with the Preferred Reporting Items for Systematic Reviews and Meta-Analyses (PRISMA 2020) guidelines. The protocol was prospectively registered in the PROSPERO database (registration number: CRD420261348962) to ensure methodological transparency and reduce the risk of reporting bias.

The primary objective of the review was to identify and critically appraise studies applying AI and ML methods to the analysis of cardiac biomarkers - biochemical, functional, and clinical - in patients after MI. The review focused specifically on identifying (i) methodological trends, (ii) recurring methodological limitations, and (iii) best-practice recommendations for future AI-driven cardiovascular research.

### 2.1 Eligibility Criteria

Studies were eligible if they:

1. included adult patients with the history of MI,
2. applied AI or ML algorithms to the analysis of cardiac biomarkers (biochemical, functional, or clinical),
3. reported model performance and validation metrics.

Eligible study designs included original empirical research, such as retrospective, prospective, or registry-based studies.

Studies were excluded if they:

- focused exclusively on acute MI diagnosis without predictive modeling,
- lacked an identifiable AI/ML analytical component,
- were reviews, editorials, commentaries, or conference abstracts without full methodological data,
- involved animal or *in vitro* models,
- or provided insufficient methodological detail upon full-text assessment.

Only English-language full-text publications published between 1 January 2015 and 30 September 2025 were included. Preprints with reporting complete methodological results were eligible and were analyzed separately in sensitivity analyses.

The full set of eligibility criteria applied during study selection is summarized in Supplementary Table S1.

### 2.2 Information Sources

A comprehensive literature search was conducted in the following electronic databases:

- PubMed/MEDLINE
- Scopus
- Web of Science Core Collection
- IEEE Xplore
- ACM Digital Library

The search covered publications from 1 January 2015 to 30 September 2025. This time frame was selected to capture the rapid growth of ML and deep learning applications in biomedical research following major advances in modern AI architectures after 2015.

To increase retrieval coverage and minimize publication bias, the search was supplemented by:

- screening the first 200 results in Google Scholar, and
- manual examination of reference lists of all eligible articles.

Preprints from medRxiv, bioRxiv, and arXiv were also considered when methodological descriptions and results were available. Sensitivity analyses excluding preprints were conducted to evaluate their influence on the final synthesis.

All retrieved references were imported into Mendeley Reference Manager (Elsevier, version 2.115) for reference management and duplicate removal. Duplicate records were identified automatically and verified manually.

Detailed database-specific search strategies, including Boolean operators, MeSH terms, and field filters, are provided in Supplementary Table S2.

### 2.3 Search Strategy

The search strategy was designed to identify studies applying AI and ML techniques to cardiac biomarkers in post-MI populations.

The population of interest included adult patients after MI. The intervention consisted of AI/ML-based analysis of cardiac biomarkers. Comparators included traditional statistical approaches where applicable, and the outcomes focused on predictive model performance and interpretability.

The strategy combined PICO elements (Population, Intervention, Comparator, Outcome) with methodological guidance from the CHARMS framework (Checklist for Critical Appraisal and Data Extraction for Systematic Reviews of Prediction Modelling Studies).

To ensure comprehensive coverage of both qualitative and quantitative research domains, the SPIDER tool (Sample, Phenomenon of Interest, Design, Evaluation, Research type) was initially used to define broad inclusion domains, whereas the CHARMS framework guided the structure of data extraction and methodological appraisal.

### 2.4 Research Questions

The systematic review addressed the following research questions (RQ), which guided data extraction and synthesis:

RQ1. What types of cardiac biomarkers (clinical, biochemical, functional) are most commonly used in AI/ML models for patients after MI?

RQ2. Which AI and ML algorithms are applied to analyze cardiac biomarkers in post-MI populations?

RQ3. What model validation strategies are used, and what methodological limitations are associated with them?

RQ4. Which model evaluation metrics are reported, and whether they are appropriate for clinical prediction tasks?

RQ5. How do studies report model interpretability and explainability?

RQ6. What are the most common reporting and methodological limitations observed in the literature?

RQ7. Based on the available evidence, what methodological recommendations can be proposed for future AI-based biomarker studies in MI?

Full search strategies, data extraction templates, and lists of excluded studies with reasons are provided in the Supplementary Materials.

### 2.5 Case Study Dataset and Study Population

The proof-of-concept case study was conducted using a clinical biomarker dataset derived from patients with MI. The dataset comprised 152 individuals with confirmed acute MI and included 117 variables describing demographic characteristics, cardiovascular risk factors, clinical parameters, and circulating biomarkers.

Demographic and anthropometric variables included age, sex, body weight, height, and body mass index (BMI). Clinical variables captured major cardiovascular risk factors and comorbidities, including diabetes mellitus, smoking status and duration, and family history of cardiovascular disease. Additional information regarding pharmacological treatment and selected clinical outcomes was also available.

A key component of the dataset was a panel of circulating biomarkers associated with inflammation, extracellular matrix remodeling, and cardiovascular aging. These included soluble α-Klotho, fibroblast growth factor-23 (FGF-23), inflammatory cytokines such as tumor necrosis factor-α (TNF-α) and interleukin-6 (IL-6), and matrix-remodeling markers including matrix metalloproteinases (MMP-2 and MMP-9) and extracellular matrix metalloproteinase inducer (EMMPRIN). Biomarker concentrations were measured in plasma samples collected during hospitalization.

Prior to analysis, all patient identifiers were removed, and only anonymized records were used.

Importantly, the dataset was used exclusively to demonstrate the feasibility of applying AI approaches within a controlled methodological framework for biomarker-based cardiovascular research. The prediction task defined in this study (binary classification of STEMI vs NSTEMI) represents a diagnostic subtyping problem rather than a clinically relevant post-MI prognostic outcome. This task was intentionally selected as a methodological proxy to enable controlled evaluation of the leakage-aware modeling workflow under realistic data conditions, including heterogeneous feature composition and potential sources of information leakage.

Accordingly, the case study should be interpreted as a methodological illustration rather than a definitive clinical validation study, and its results are not intended to support prognostic inference regarding post-MI outcomes.

### 2.6 Selection Process

Two reviewers independently screened titles and abstracts of all retrieved records and subsequently assessed the full texts of potentially eligible studies according to predefined inclusion and exclusion criteria. Discrepancies were resolved through discussion and, when necessary, consultation with a third reviewer.

The study selection process followed PRISMA 2020 recommendations and is summarized in the PRISMA flow diagram (Figure 1).

**Figure 1.**
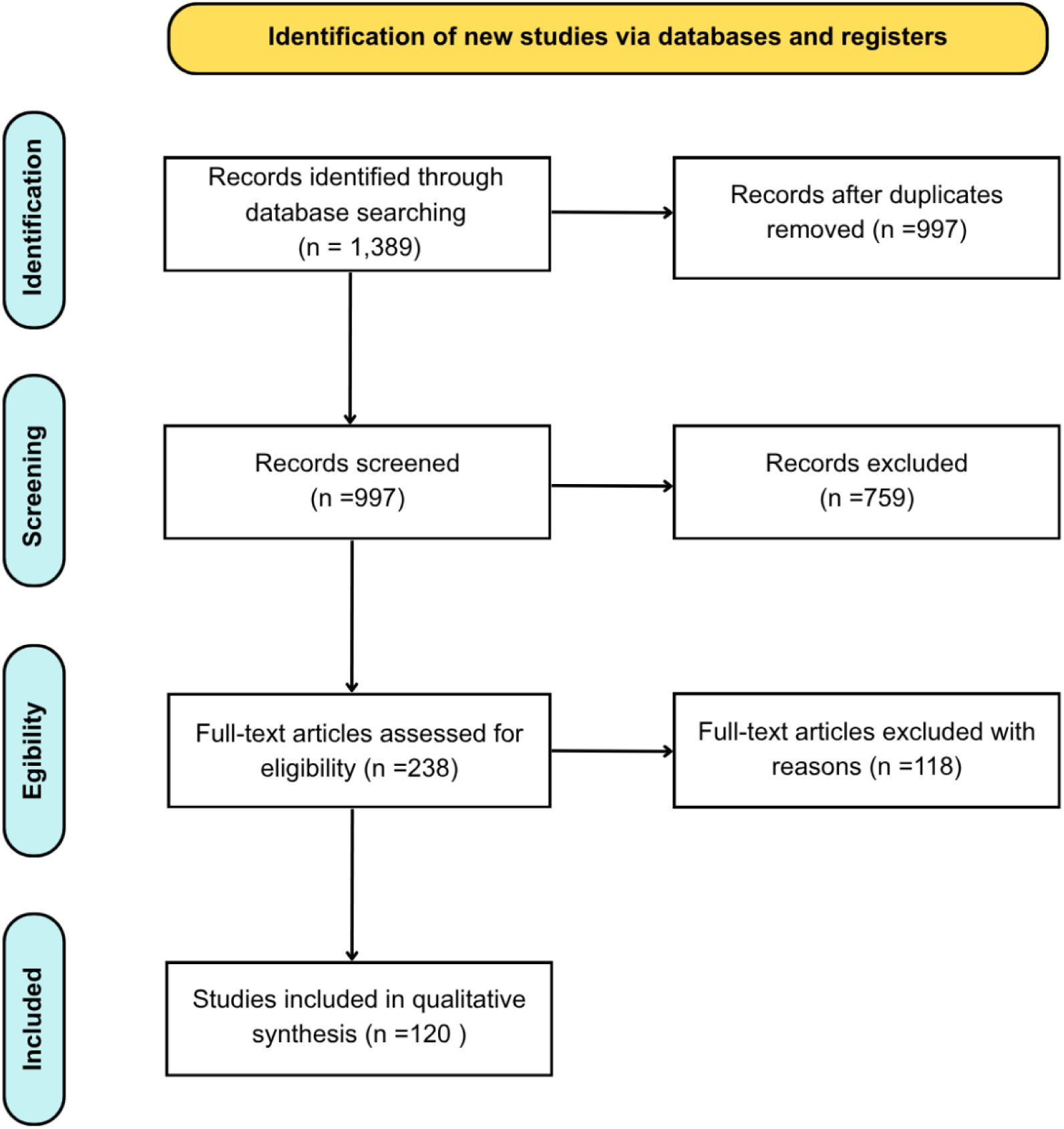
PRISMA flow diagram of the study selection process.

### 2.7 Data Extraction

Data extraction was conducted independently by two reviewers using a pre-specified CHARMS-based extraction form implemented using Microsoft Excel (template available in the Supplementary Material). Discrepancies between reviewers were resolved through discussion or adjudication by a third reviewer.

The following variables were extracted:

- population characteristics and biomarker categories (biochemical, functional, clinical)
- data sources and sample size (registries, clinical cohorts, electronic health records, biobanks)
- AI/ML algorithms and feature engineering strategies
- preprocessing and data normalization procedures
- model validation approaches
- model performance metrics
- interpretability and explainability methods
- availability of code, model weights, or supplementary data supporting reproducibility.

A complete list of all 120 included studies, together with country/centre and primary study objective, is provided in Supplementary Table S3.

### 2.8 Risk of Bias and Quality Assessment

The methodological quality of included studies was evaluated using the Prediction Model Risk of Bias Assessment Tool (PROBAST). Where applicable, selected elements from the emerging PROBAST-AI extension were incorporated to account for AI-specific methodological considerations.

The assessment covered four standard PROBAST domains:

- participants,
- predictors,
- outcomes,
- analysis.

Each domain was rated as low, high, or unclear risk of bias according to PROBAST guidance.

In addition to the core PROBAST domains, AI-specific aspects were explicitly evaluated, including data preprocessing procedures, feature selection strategies, validation design, and model interpretability. Particular attention was given to potential sources of information leakage, improper handling of predictors, and inconsistencies in model validation workflows.

Reporting transparency and completeness were further assessed using selected items derived from the TRIPOD-AI reporting checklist, focusing on the clarity of model development, validation, and reproducibility practices.

Risk-of-bias assessments were performed independently by two reviewers. Discrepancies were resolved through discussion and, when necessary, consultation with a third reviewer.

Given the scale and heterogeneity of included studies, results of the quality assessment are presented as aggregated domain-level summaries in Supplementary Table S4.

### 2.9 Reporting Bias and Certainty Assessment

Potential reporting and publication bias were addressed by including preprints and grey literature sources during the search process. When sufficient comparable studies were available, publication bias was explored using funnel plots and Egger’s regression tests where appropriate.

The certainty of evidence across outcomes was assessed using a modified GRADE framework adapted for prediction modeling studies. The evaluation considered risk of bias, consistency of findings, precision, directness of evidence, and potential publication bias.

Certainty assessments were conducted independently by two reviewers, with disagreements resolved through consensus. Results are summarized in Supplementary Table S5.

### 2.10 Effect Measures

Because of heterogeneity in study designs and reported outcomes, a range of model performance measures was extracted. Primary performance measures included discrimination metrics such as:

- area under the receiver operating characteristic curve (ROC-AUC),
- F1-score,
- Matthews correlation coefficient (MCC),
- accuracy and balanced accuracy.

When available, calibration metrics were also extracted, including calibration slope, Brier score, and Hosmer–Lemeshow statistics.

Decision-analytic measures (e.g., decision curve analysis and net benefit) and interpretability indices (e.g., SHAP values, permutation importance, and feature importance) were additionally recorded to evaluate clinical relevance and model explainability.

### 2.11 Data Synthesis

Given the expected methodological heterogeneity, both quantitative and narrative synthesis approaches were applied.

When studies reported comparable performance metrics and outcome definitions, meta-analysis using random-effects models was initially planned. However, due to substantial methodological heterogeneity across studies, a formal meta-analysis was not ultimately conducted.

For outcomes that were not directly comparable across studies (e.g., interpretability approaches or decision-analytic metrics), findings were synthesized narratively.

Subgroup analyses were conducted according to:

- biomarker category
- machine-learning model class
- validation strategy (internal vs external)
- data source (clinical cohorts vs registries).

Sensitivity analyses excluding preprints were performed to evaluate their potential impact on the overall conclusions.

## 3. Current Landscape of AI in Cardiac Biomarkers after MI

### 3.1 Overview of Included Studies

The systematic review identified 120 eligible studies from 1,389 records retrieved across multiple databases (Figure 1). After removal of duplicates and screening, 238 full-text articles were assessed and 120 met the inclusion criteria.

The characteristics of the included studies are summarized in Table 1, while the complete study-level inventory of all 120 included articles is provided in Supplementary Table S3. Most investigations relied on observational clinical datasets, frequently derived from electronic health records, hospital registries, or prospective cohorts. Retrospective designs predominated (86/120; 71.7%), whereas 43 studies (35.8%) included a prospective component. Cohort-based designs were reported in 71 studies (59.2%).

**Table 1.**
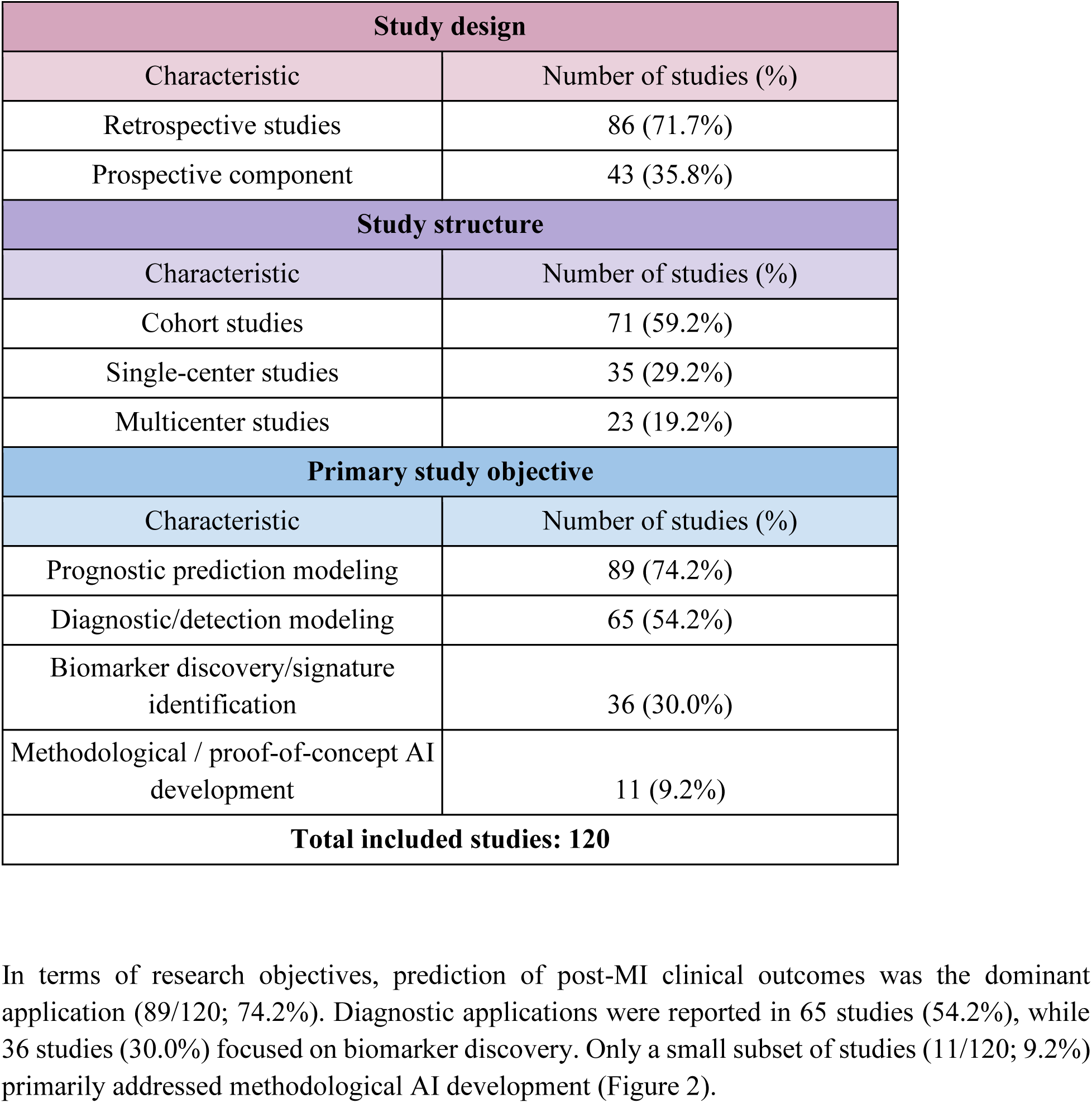
Characteristics of the studies included in the systematic review.

In terms of research objectives, prediction of post-MI clinical outcomes was the dominant application (89/120; 74.2%). Diagnostic applications were reported in 65 studies (54.2%), while 36 studies (30.0%) focused on biomarker discovery. Only a small subset of studies (11/120; 9.2%) primarily addressed methodological AI development (Figure 2).

**Figure 2.**
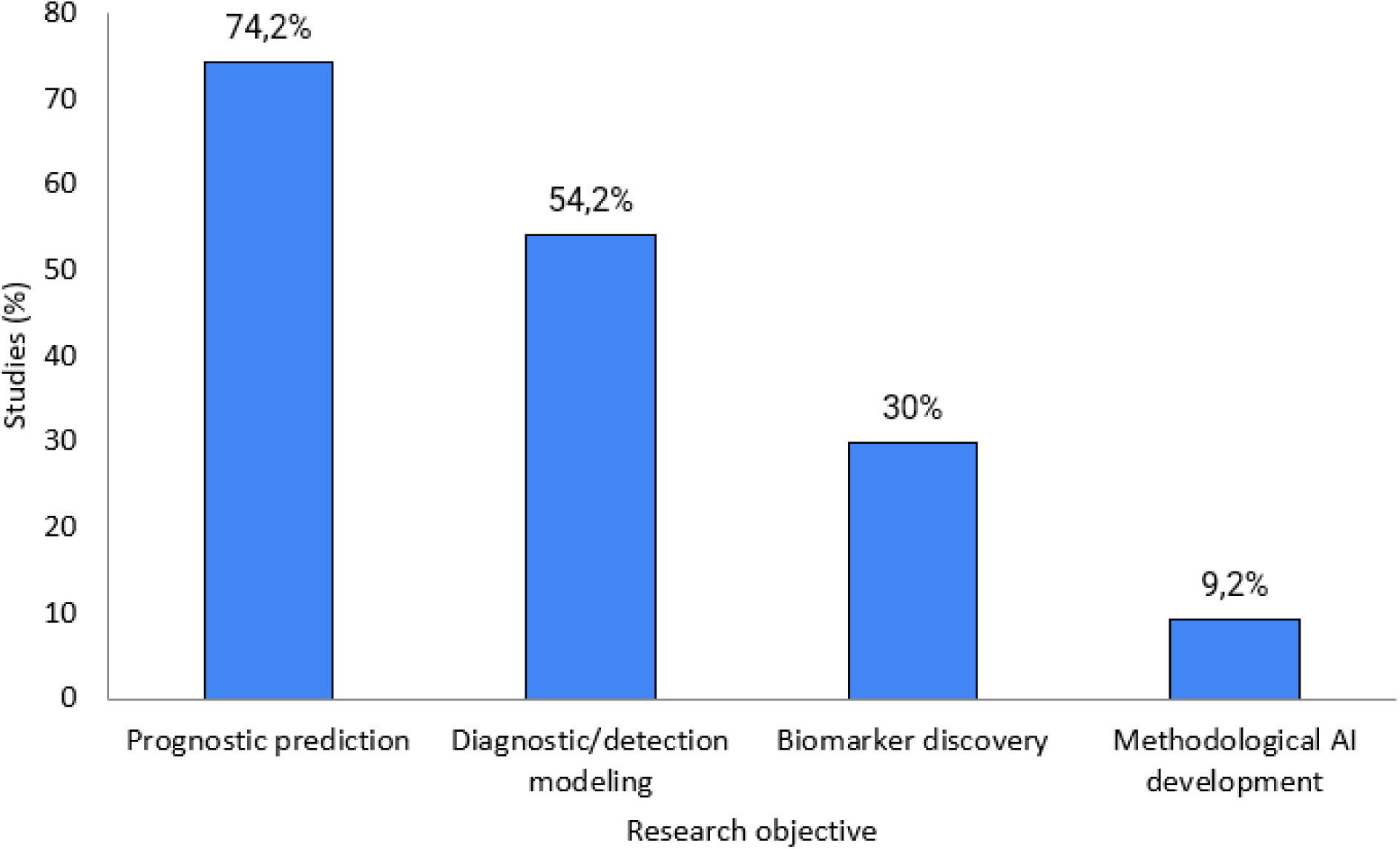
Distribution of research objectives in studies applying AI/ML to cardiac biomarkers after MI.

### 3.2 Types of Biomarkers Used in AI Models

Most studies implemented multimodal input structures combining biochemical biomarkers with clinical or physiological variables. Overall, 109 of 120 studies (90.8%) integrated biomarker measurements with demographic, clinical, imaging, or physiological features. The distribution of biomarker categories used in AI models is summarized in Table 2 and Figure 3.

**Figure 3.**
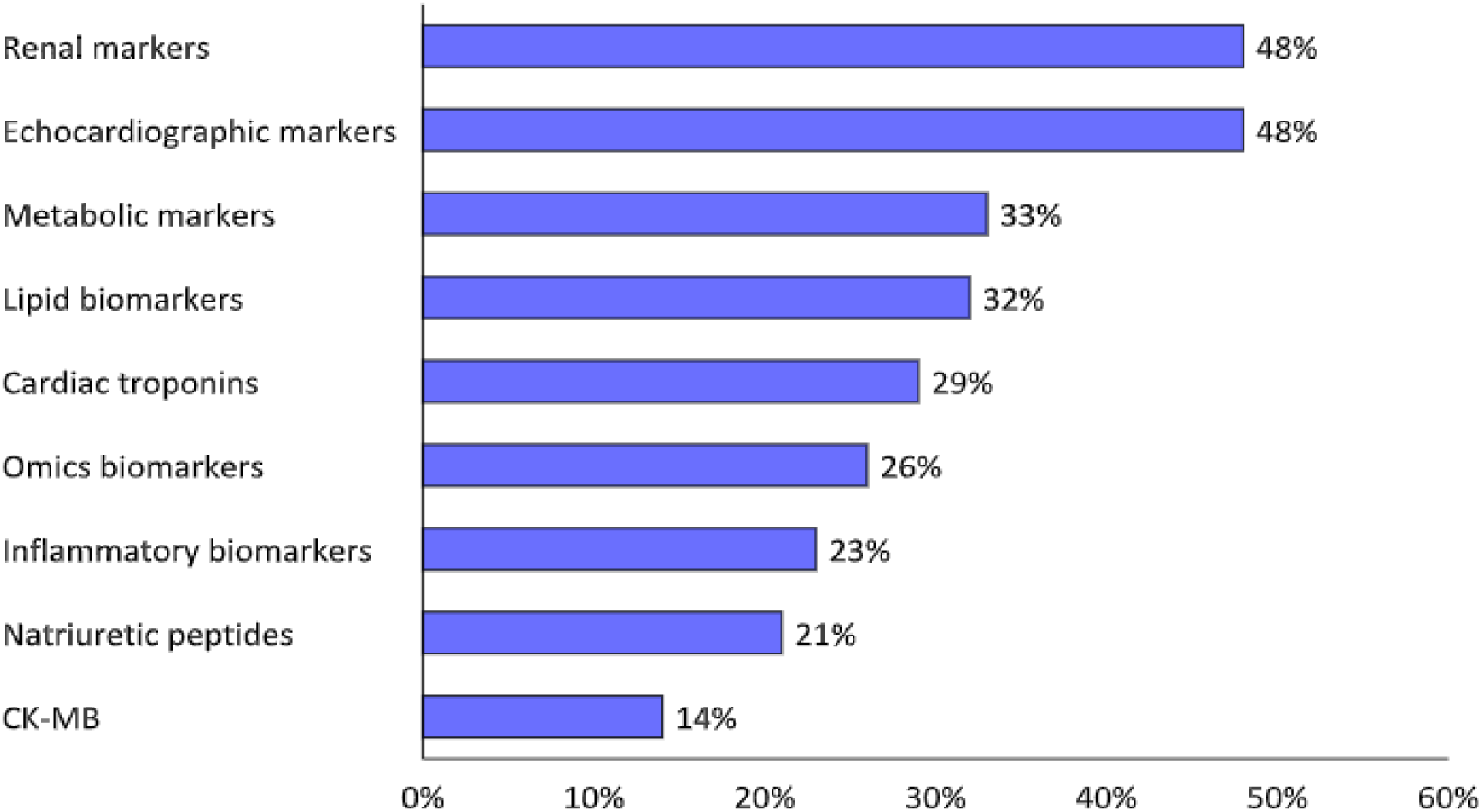
Frequency of biomarker categories used in AI models for post-MI prediction.

**Table 2.**
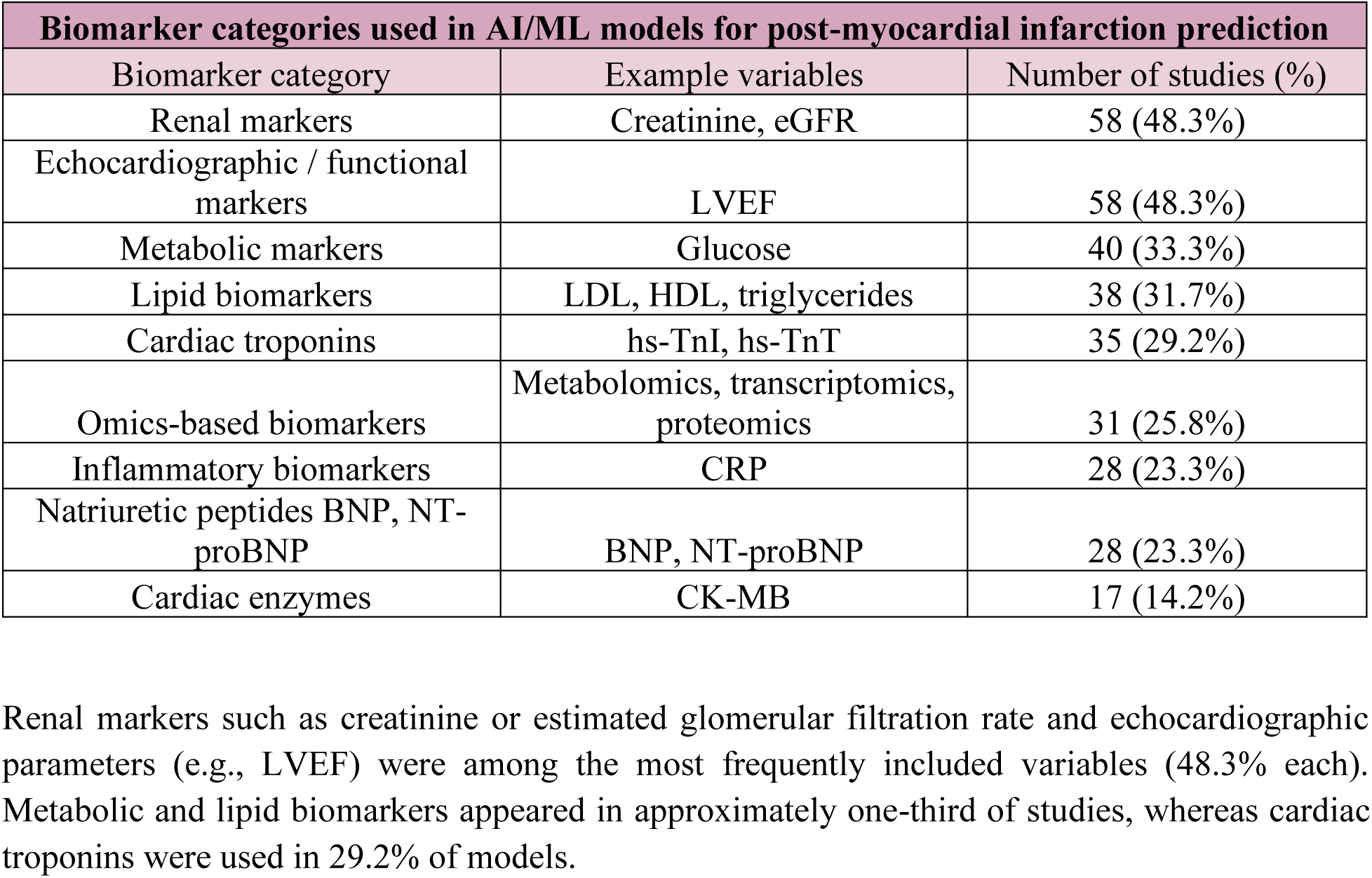
Biomarker categories used in AI/ML models for post-myocardial infarction prediction.

Renal markers such as creatinine or estimated glomerular filtration rate and echocardiographic parameters (e.g., LVEF) were among the most frequently included variables (48.3% each). Metabolic and lipid biomarkers appeared in approximately one-third of studies, whereas cardiac troponins were used in 29.2% of models.

Emerging omics-based biomarkers were incorporated in 25.8% of studies. Inflammatory markers, natriuretic peptides, and creatine kinase-MB were used less frequently.

Overall, the predominance of multimodal datasets reflects the multifactorial nature of cardiovascular risk after MI.

### 3.3 AI and ML Approaches

A wide range of ML algorithms was applied across the included studies (Table 3, Figure 4).

**Figure 4.**
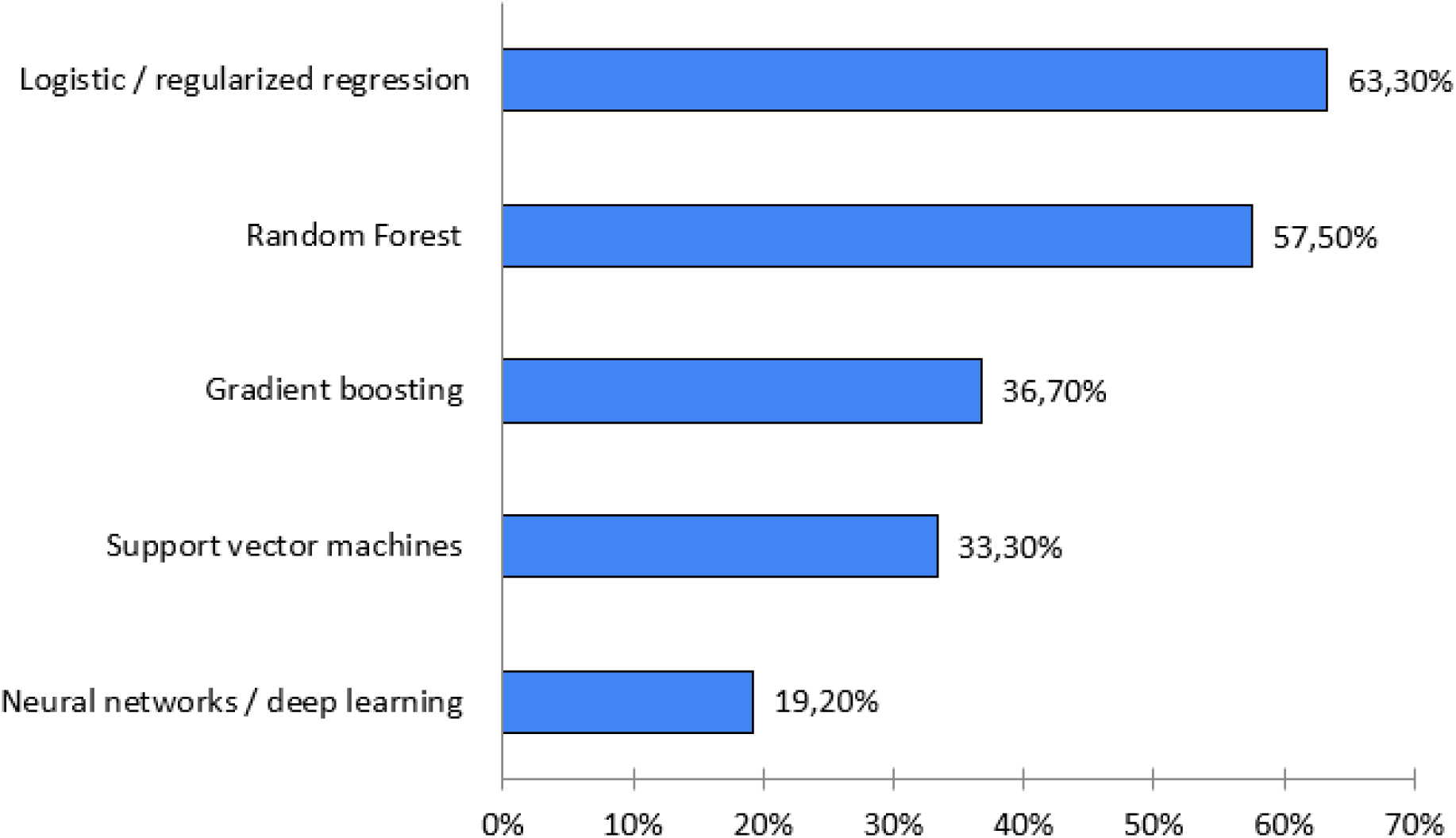
Frequency of AI/ML algorithm categories used across included studies.

**Table 3.** AI and ML approaches used in studies of cardiac biomarkers after MI.

| AI and machine learning approaches used in studies of cardiac biomarkers after myocardial infarction |  |  |
| --- | --- | --- |
| Algorithm category | Representative methods | Number of studies (%) |
| Logistic / regularized regression | Logistic regression, LASSO, ridge, elastic net | 76 (63.3%) |
| Random Forest | Random Forest | 69 (57.5%) |
| Gradient boosting | XGBoost, LightGBM, boosting-based ensembles | 44 (36.7%) |
| Support vector machines | Linear SVM, kernel SVM | 40 (33.3%) |
| Neural networks / deep learning | MLP, ANN, deep learning architectures | 23 (19.2%) |

Logistic or regularized regression models were the most frequently reported approaches (76/120; 63.3%), followed by Random Forest algorithms (69/120; 57.5%). Gradient boosting methods, including XGBoost and LightGBM, were reported in 44 studies (36.7%), while support vector machines were used in 40 studies (33.3%). Neural networks or deep learning architectures appeared less frequently (23/120; 19.2%).

Because many studies compared several algorithms within the same analysis, these categories are not mutually exclusive. Overall, the methodological landscape reflects a combination of traditional statistical approaches and modern machine learning algorithms applied predominantly to structured clinical datasets.

### 3.4 Model Validation Strategies

Validation approaches varied substantially across the reviewed studies (Table 4, Figure 5).

**Figure 5.**
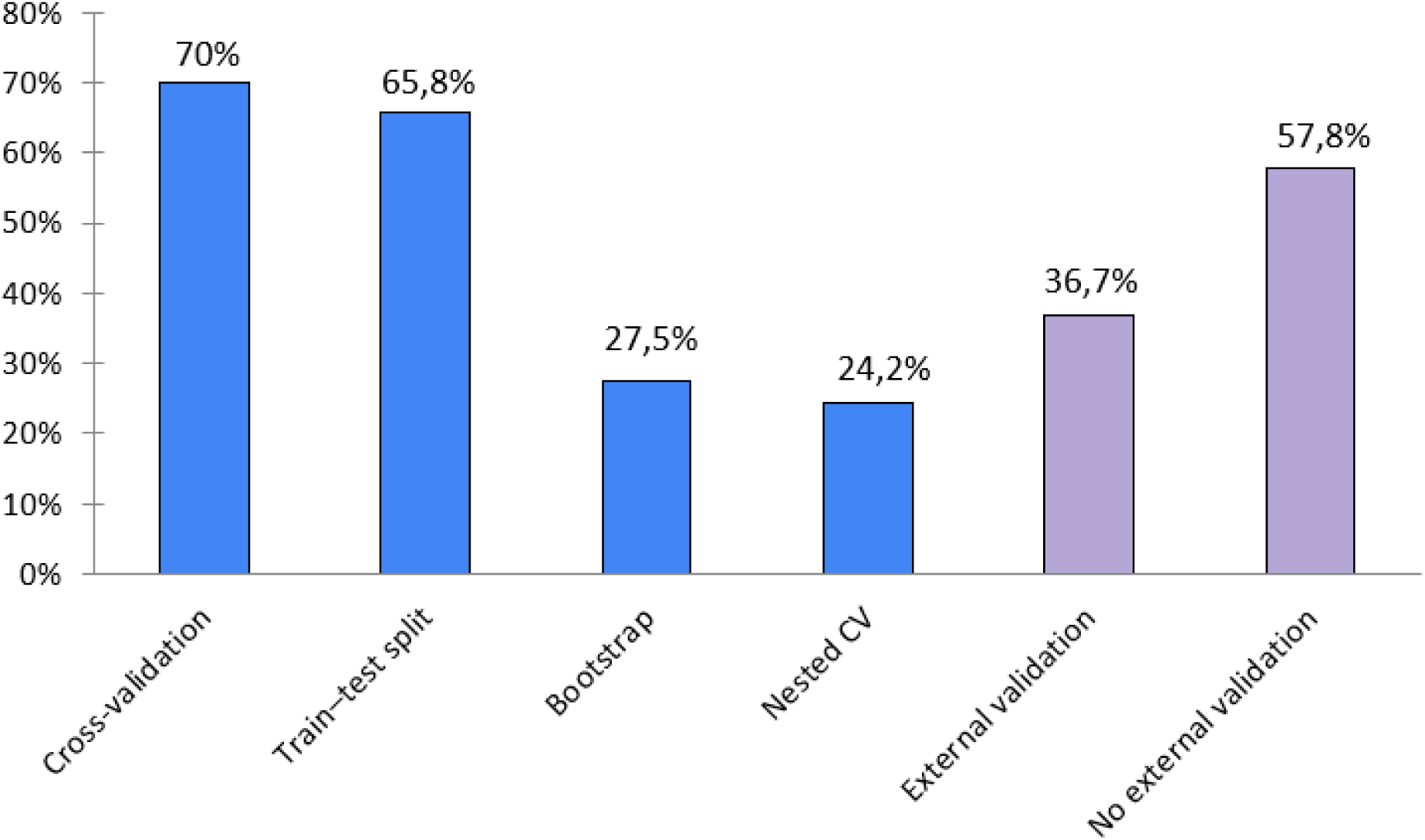
Distribution of internal and external validation strategies in AI studies of cardiac biomarkers after MI.

**Table 4.**
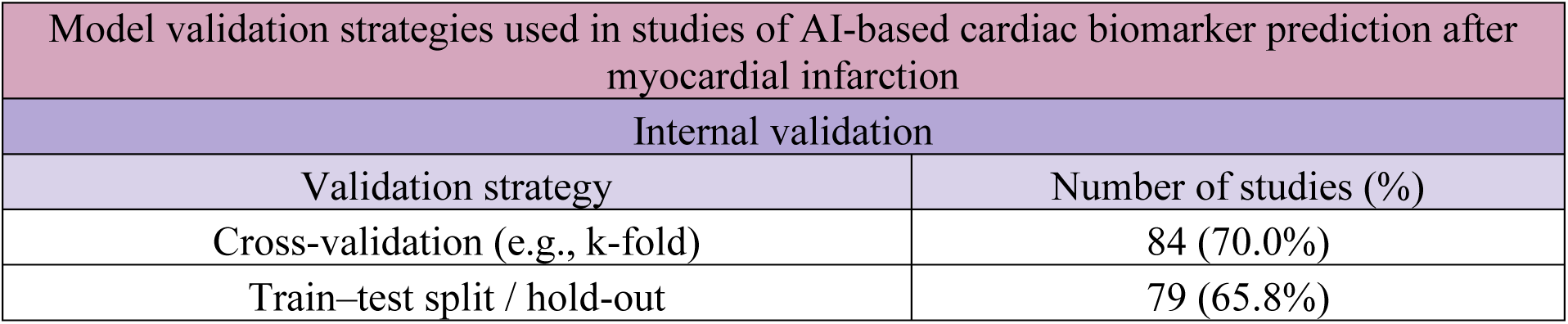
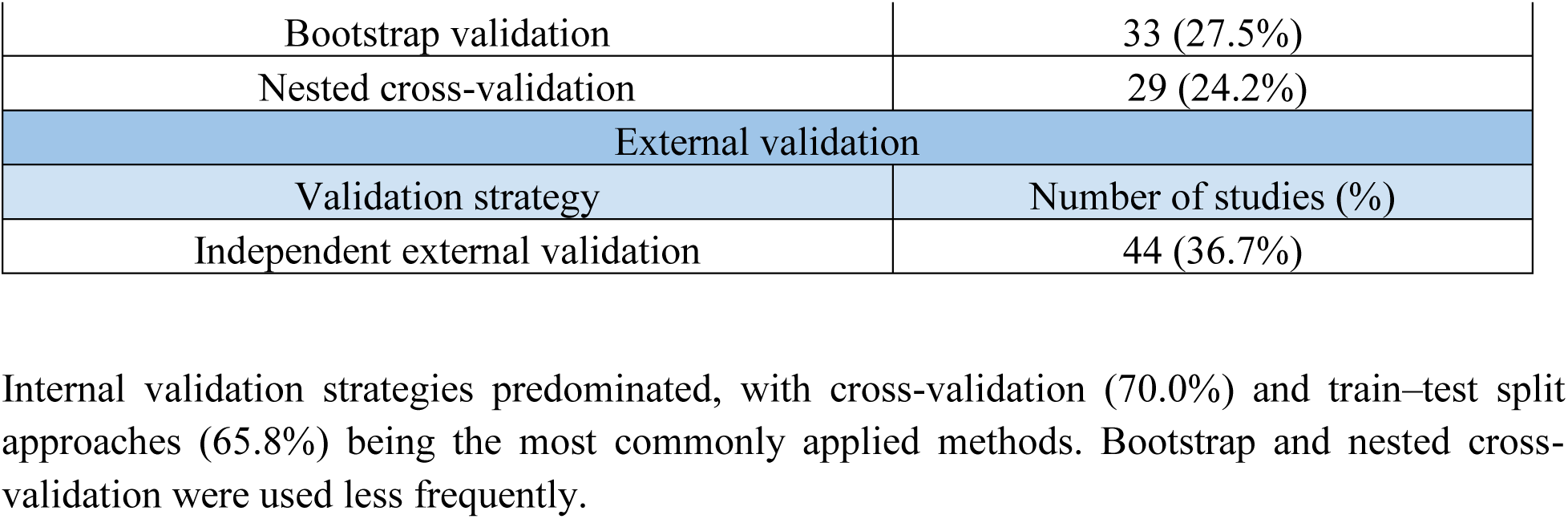
Model validation strategies used in studies of AI-based cardiac biomarker prediction after MI.

Internal validation strategies predominated, with cross-validation (70.0%) and train–test split approaches (65.8%) being the most commonly applied methods. Bootstrap and nested cross-validation were used less frequently.

In contrast, independent external validation was reported in only 44 of 120 studies (36.7%). Detailed methodological coding in a subset of studies indicated that the majority did not perform any external validation.

Among studies that implemented external validation, geographical or multicenter validation was the most common approach, followed by temporal and cross-dataset validation (Figure 6).

**Figure 6.**
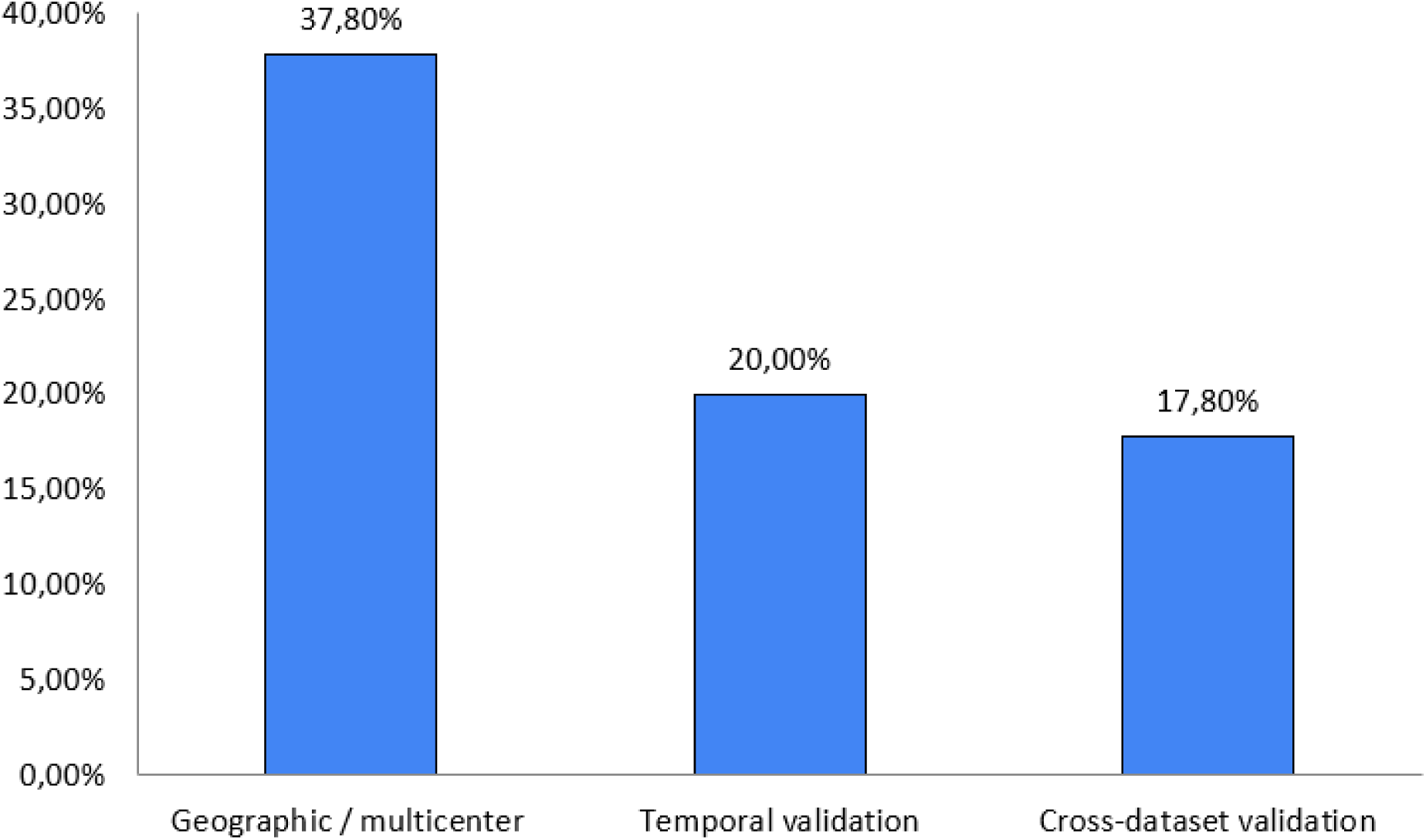
Types of external validation approaches reported across studies.

Overall, these findings indicate that while internal validation is widely adopted, rigorous evaluation of model generalizability across independent datasets remains limited.

### 3.5 Model Performance Metrics

Model performance was evaluated predominantly using discrimination-based metrics (Table 5, Figure 7).

**Figure 7.**
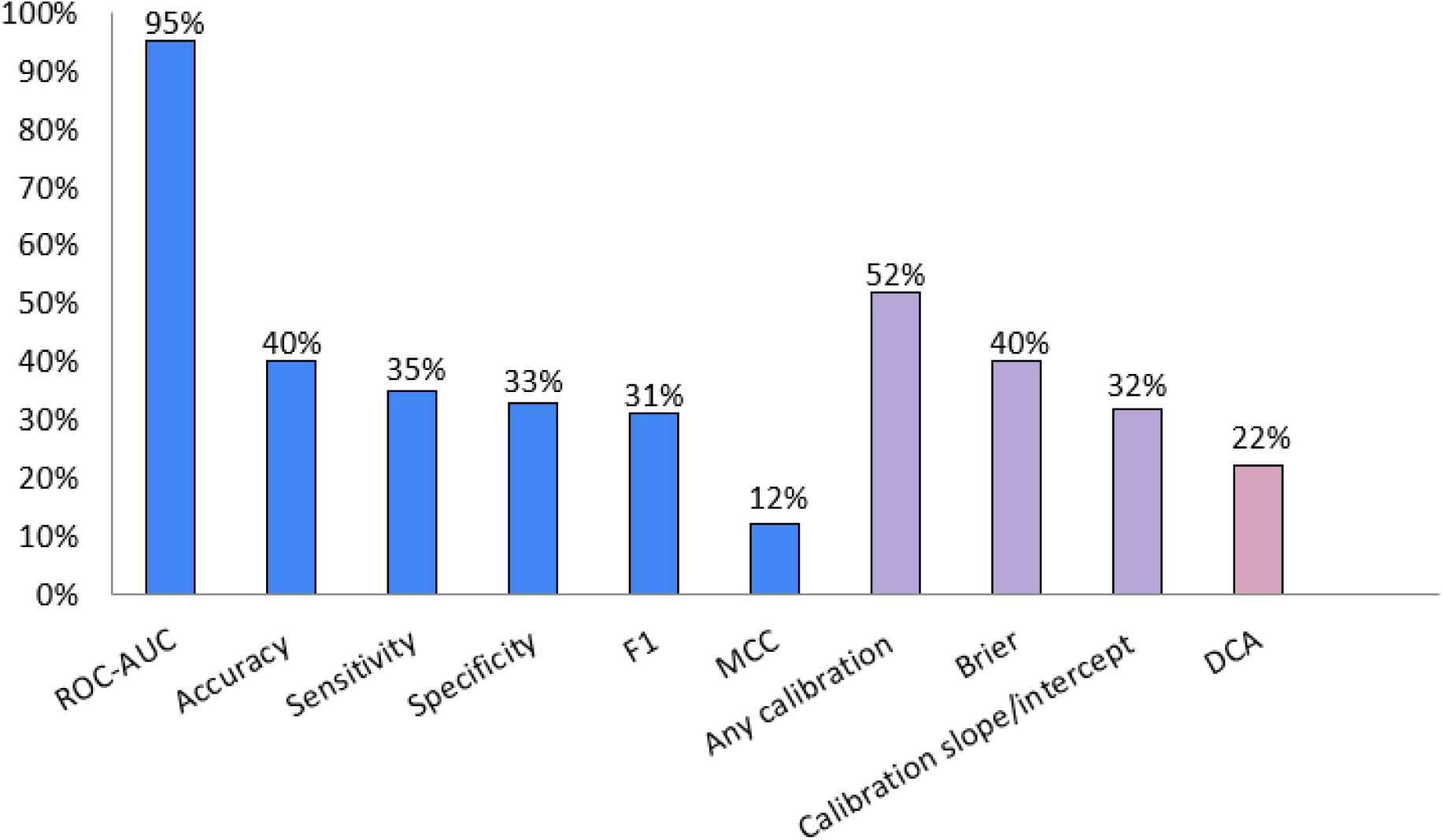
Reporting frequency of discrimination, calibration, and clinical utility metrics in AI studies of cardiac biomarkers.

**Table 5.** Model performance metrics reported in AI studies of cardiac biomarkers after MI.

| Discrimination metrics |  |  |
| --- | --- | --- |
| Metric category | Specific metric | Number of studies (%) |
| ROC-AUC | Area under ROC curve | 114 (95.0%) |
| Accuracy | Classification accuracy | 48 (40.0%) |
| Sensitivity | Recall (true positive rate) | 42 (35.0%) |
| Specificity | True negative rate | 40 (33.3%) |
| F1 score | Harmonic mean of precision and recall | 38 (31.7%) |
| Precision–recall metrics | PR-AUC or precision/recall | 28 (23.3%) |
| MCC | Matthews correlation coefficient | 15 (12.5%) |
| Calibration metrics |  |  |
| Metric category | Specific metric | Number of studies (%) |
| Any calibration analysis | Any reported method | 47 (52.2%) |
| Brier score | Probabilistic accuracy | 36 (40.0%) |
| Calibration slope/intercept | Calibration parameters | 29 (32.2%) |
| Clinical utility |  |  |
| Metric category | Specific metric | Number of studies (%) |
| Decision curve analysis | Net benefit analysis | 20 (22.2%) |

The most commonly reported measure was ROC-AUC, appearing in 95.0% of studies. Other metrics such as accuracy, sensitivity, specificity, and F1 score were reported less frequently.

In contrast, calibration assessment and clinical utility evaluation were less consistently reported. Among studies with detailed methodological coding, only 52.2% reported any form of calibration analysis, while decision curve analysis was performed in 22.2% of studies.

These findings highlight a substantial imbalance between discrimination-focused evaluation and clinically oriented performance assessment (Figure 8).

**Figure 8.**
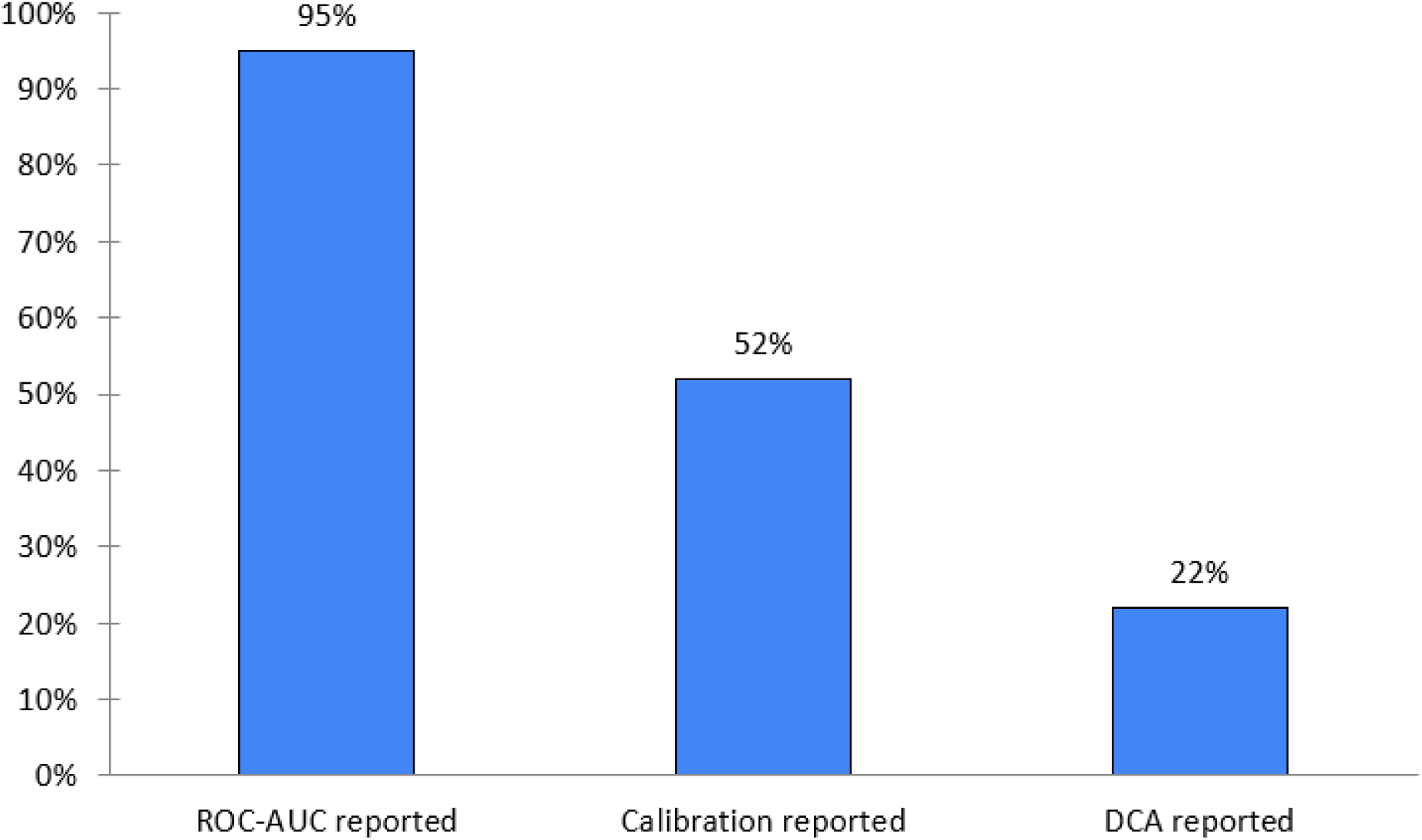
Disparity between discrimination-focused and clinically oriented evaluation metrics.

### 3.6 Model Interpretability and Explainability

Interpretability approaches were reported inconsistently across the reviewed studies (Table 6, Figure 9). Among formal explainability techniques, SHAP was the most frequently used method (26.7%), followed by LIME (10.8%).

**Figure 9.**
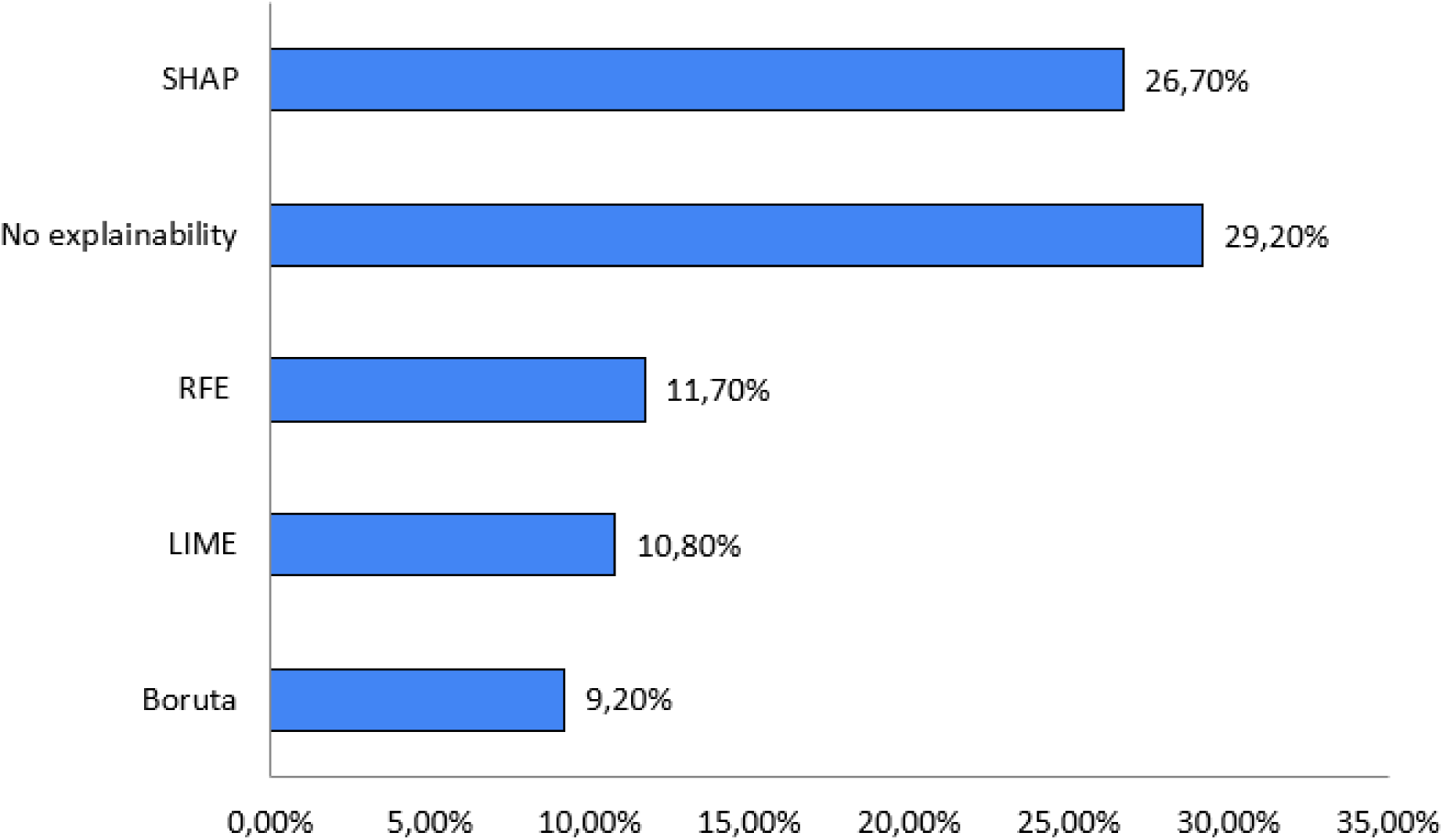
Reporting frequency of interpretability and explainability methods in AI studies of cardiac biomarkers.

**Table 6.**
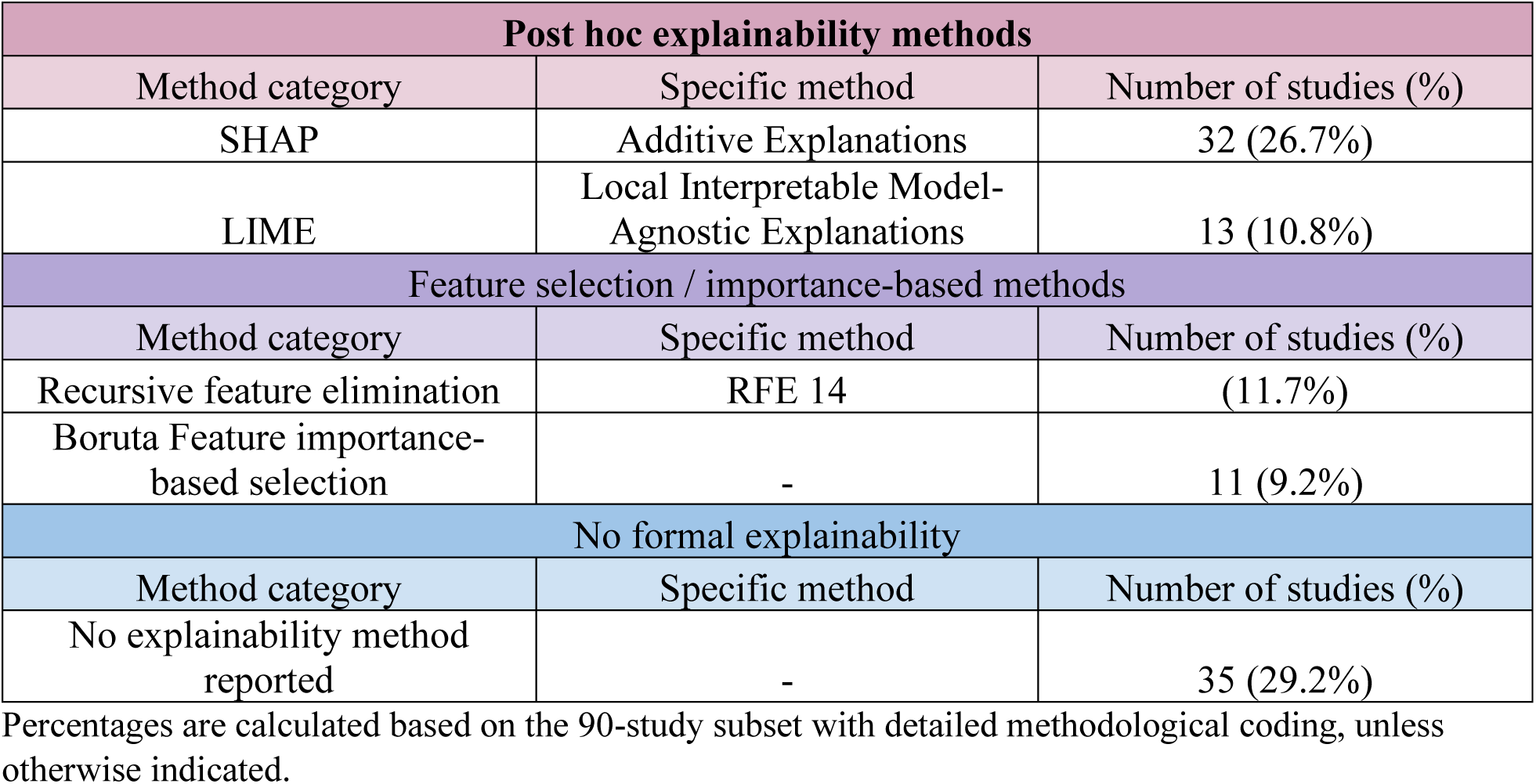
Interpretability and explainability methods reported in AI studies of cardiac biomarkers after MI.

Feature selection–based approaches, such as recursive feature elimination and Boruta, were applied in a smaller proportion of studies. Notably, 29.2% of studies explicitly reported no formal explainability method.

In many cases, interpretation was limited to variable importance rankings, regression coefficients, or qualitative discussion of biologically plausible predictors. Overall, the level of model interpretability varied considerably across the literature.

### 3.7 Reporting Quality and Methodological Transparency

The systematic review identified recurring limitations related to reporting transparency and reproducibility (Table 7, Figure 10). Among studies with extended methodological coding, only 13.3% provided publicly available code, while the majority did not make code publicly available.

**Figure 10.**
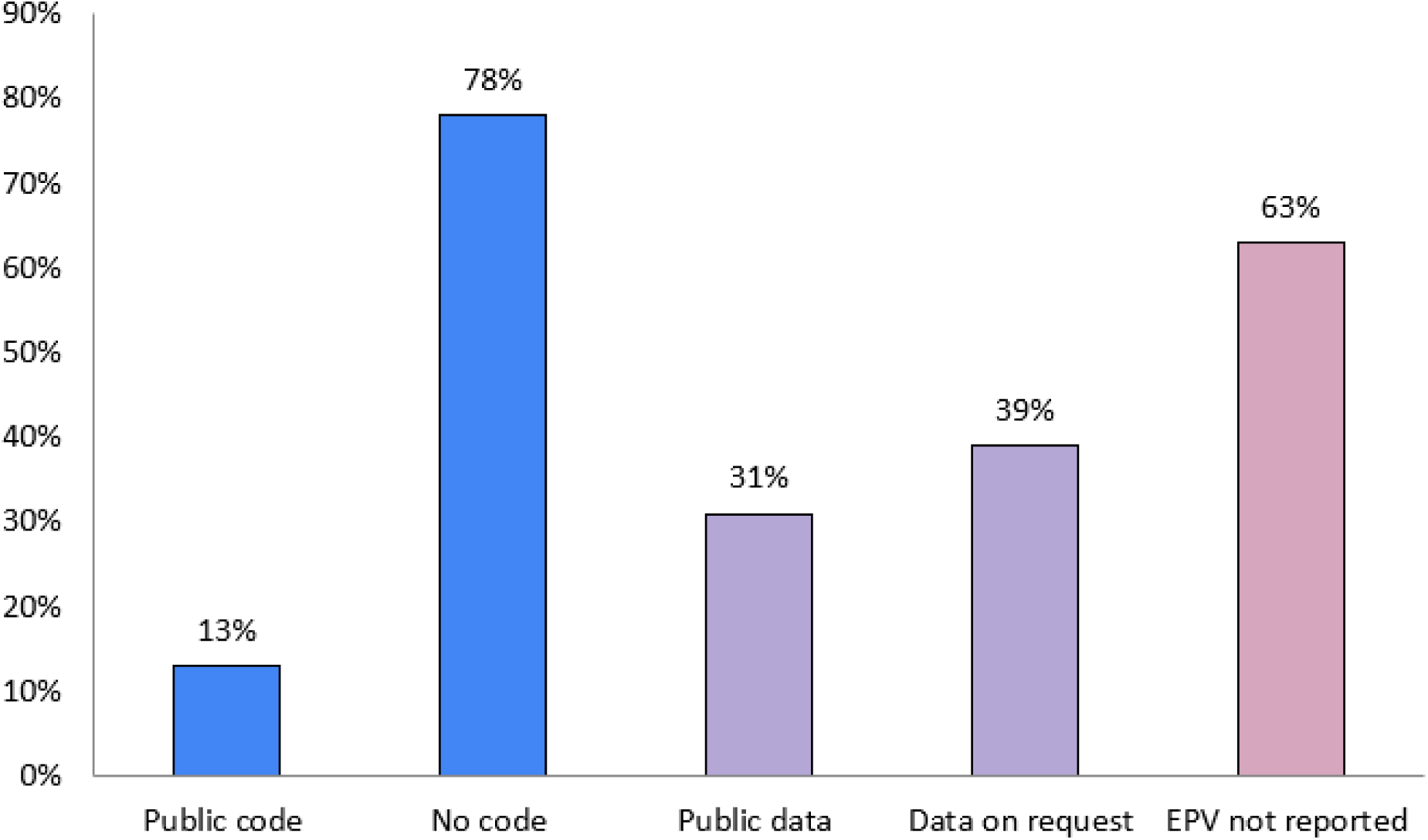
Reporting practices related to reproducibility and transparency in AI studies of cardiac biomarkers.

**Table 7.**
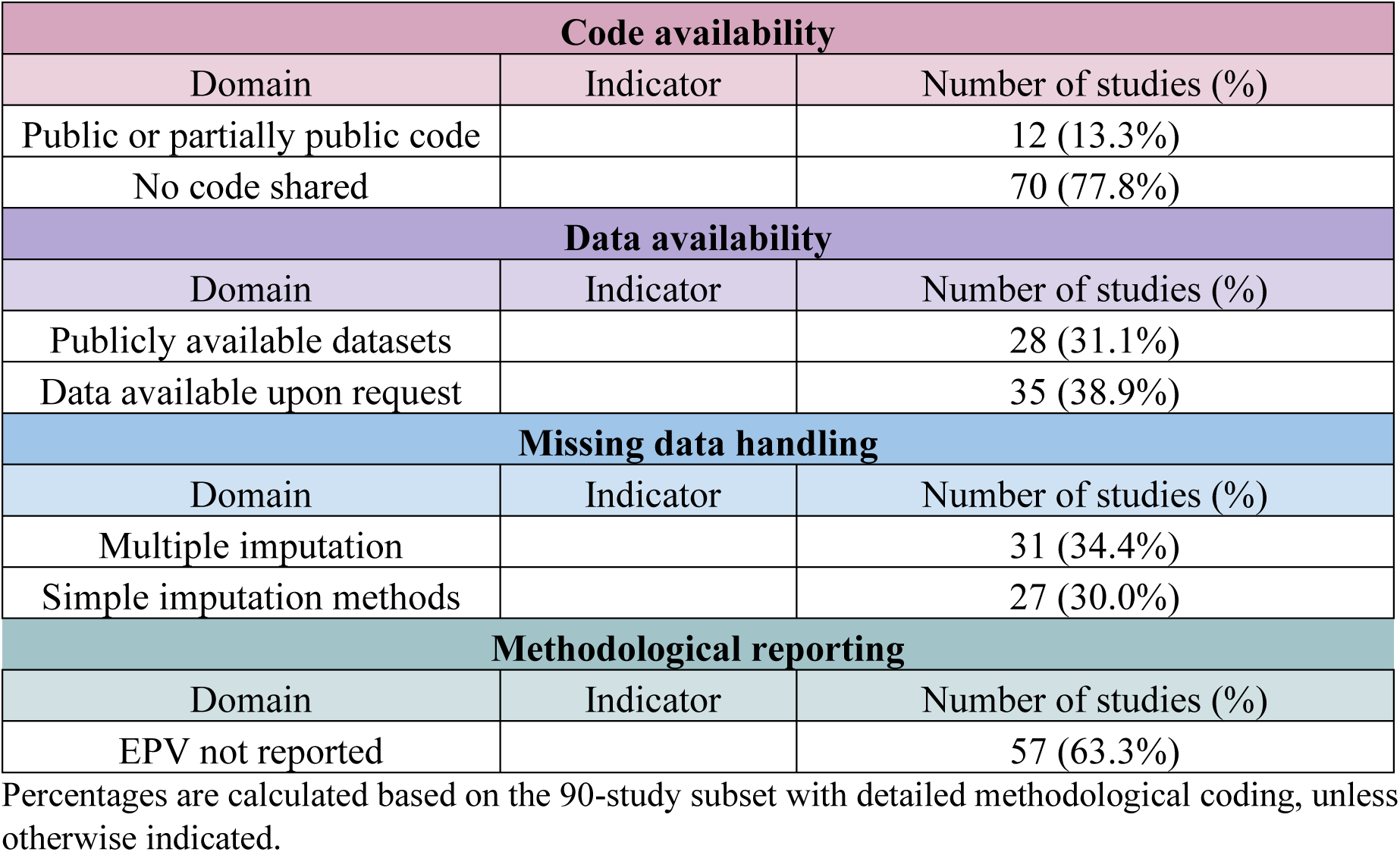
Reporting transparency and reproducibility practices in AI studies of cardiac biomarkers after MI.

Public datasets were reported in 31.1% of studies, whereas data were often available only upon request. Strategies for handling missing data varied, with both multiple imputation and simpler approaches commonly used.

Importantly, key methodological aspects such as events-per-variable considerations were frequently not reported, limiting assessment of potential overfitting.

Overall, these findings highlight persistent limitations in methodological transparency and reproducibility across the literature.

### 3.8 Summary of quantitative patterns in the reviewed literature

Several quantitative patterns emerged across the included studies (Figure 11). First, multimodal modeling was the dominant approach, observed in 90.8% of studies, reflecting the integration of biomarkers with broader clinical datasets. Second, the literature was strongly oriented toward prediction and prognostic modeling, whereas biomarker discovery studies were less common.

**Figure 11.**
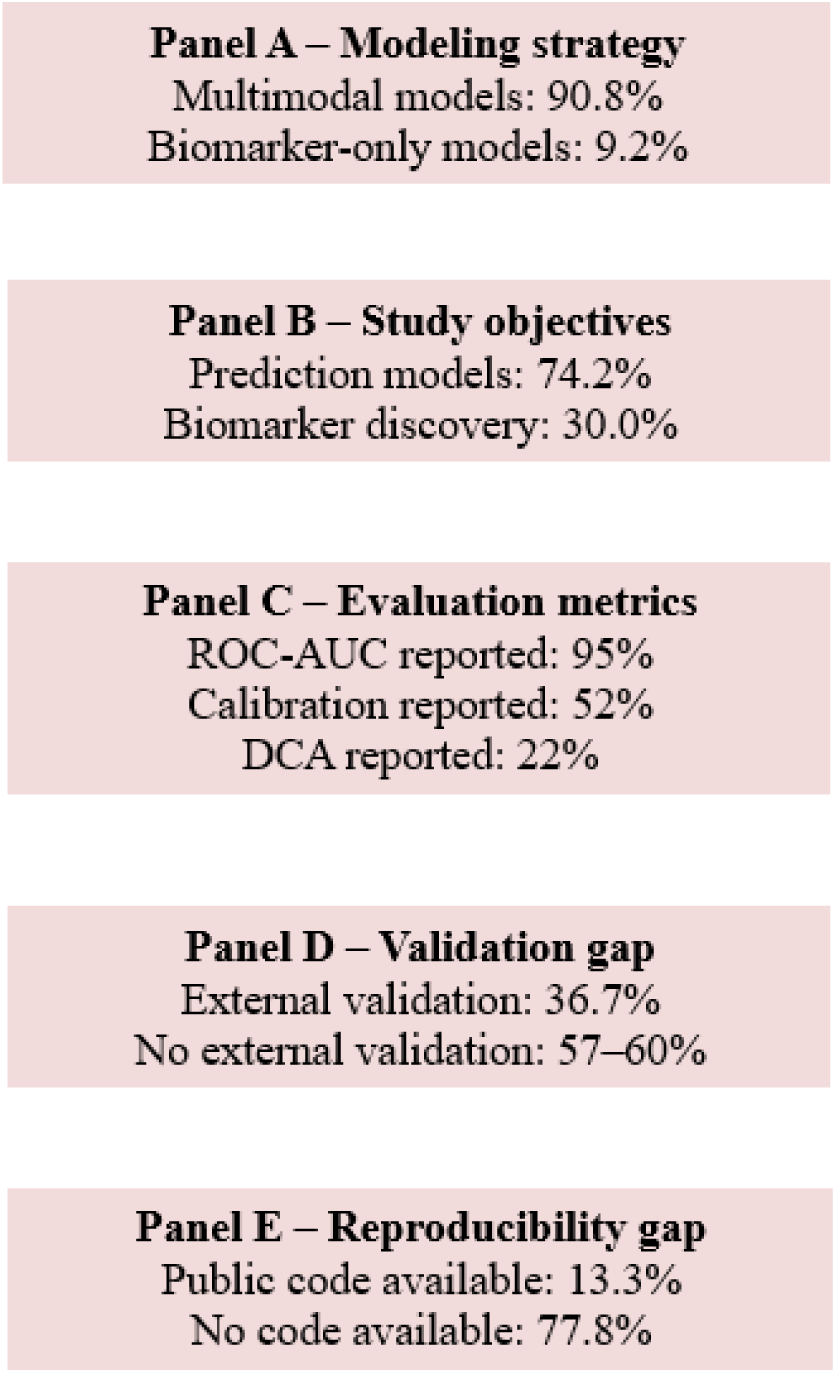
Key methodological patterns and limitations in AI studies of cardiac biomarkers after MI.

Third, model evaluation was heavily dominated by discrimination metrics, with ROC-AUC reported in 95% of studies, while calibration and decision-analytic assessment remained limited. Fourth, independent external validation was reported in only 36.7% of studies, indicating substantial gaps in generalizability assessment.

Finally, transparency and reproducibility were limited, with publicly available code reported in only 13.3% of studies.

Taken together, these findings highlight a consistent pattern of methodological imbalance, characterized by widespread use of multimodal models and discrimination-based evaluation, alongside limited external validation, insufficient clinical utility assessment, and restricted reproducibility.

## 4. Methodological framework: leakage-aware AI workflow for post–MI biomarker modeling

Following the systematic literature review, a proof-of-concept empirical case study was conducted to operationalize and critically evaluate a leakage-aware machine-learning workflow for post–MI biomarker modeling. The methodological framework described below pertains exclusively to this empirical case study and is conceptually distinct from the PRISMA-based review methodology (Figure S1).

The primary objective of this framework was not to develop or validate a clinically deployable prediction model, but rather to rigorously interrogate methodological assumptions commonly encountered in AI-driven biomarker research. In particular, the workflow was constructed to explicitly address prevention and detection of information leakage, strict separation of model selection from performance estimation, evaluation of probabilistic and decision-analytic properties beyond discrimination metrics alone, transparent and auditable feature governance, and reproducible generation of reporting-ready analytical artifacts.

Importantly, the analytical task used throughout this framework (binary classification of STEMI vs NSTEMI) represents a diagnostic subtyping problem rather than a clinically meaningful post-MI prognostic endpoint. This task was deliberately selected as a controlled methodological proxy, enabling systematic evaluation of model behavior, feature governance, and leakage susceptibility under realistic data conditions, while avoiding overinterpretation of results as clinically actionable prognosis.

The framework was implemented as a structured, multi-stage pipeline encompassing data governance, nested cross-validation benchmarking, out-of-fold performance consolidation, cross-fitted calibration, robustness and ablation analyses, locked-split diagnostic validation, and permutation-based falsification testing. All analytical steps were executed through explicit, version-controlled scripts, each generating intermediate outputs and audit logs to enable full traceability and replication of methodological decisions. Performance reporting throughout the workflow relied exclusively on out-of-fold or strictly held-out predictions, thereby minimizing optimistic bias and preventing contamination between model development and evaluation stages.

### 4.1. Stage 1 — Data preparation, quality control, and feature governance

Each row in the dataset corresponded to a unique patient, with no longitudinal structure or repeated measurements. All Stage 1 procedures were strictly outcome-agnostic and confined to data harmonization, quality control, and explicit feature governance, with the objective of producing an analysis-ready model-agnostic dataset.

At this stage, the downstream analytical task (STEMI vs NSTEMI classification) was not used in any form to guide preprocessing decisions, ensuring that all transformations remained independent of both diagnostic labels and any potential prognostic interpretation.

The original source database was imported and preserved as an immutable raw snapshot to ensure full auditability and reproducibility. Feature names originating from heterogeneous data sources were harmonized using an explicit name-mapping table, allowing bidirectional traceability between original and standardized variable identifiers. Data types were coerced using predefined, rule-based transformations. Binary textual encodings (e.g., YES/NO equivalents) were normalized prior to numeric coercion to prevent unintended feature collapse. All type conversions - including non-parsable values and coercion-induced missingness - were systematically logged to enable retrospective inspection of data integrity decisions (Table S6).

Patterns of missingness were quantitatively assessed at both the individual feature level and the broader feature-category level. No outcome-driven filtering, threshold-based exclusion, or missingness-informed feature removal was performed at this stage. The overall dataset structure - including the number of patients, outcome prevalence, feature composition by governance category, and missingness patterns -is summarized in Table S7.

In parallel, values falling outside physiologically plausible ranges were identified through a dedicated quality-control audit. These observations were neither automatically removed nor winsorized. Instead, they were retained and explicitly flagged to ensure that quality control remained descriptive rather than interventional during the governance phase.

A structured data dictionary was constructed to document feature semantics, measurement units, clinical provenance, and assigned governance role. As part of an initial leakage firewall, a first-pass screening was performed using keyword-based pattern matching to identify variables potentially entangled with post-baseline information, downstream clinical processes, procedural interventions, or treatment decisions. Variables deemed non-interpretable, identifier-like, temporally post-baseline by definition, or incompatible with cross-sectional modeling assumptions were excluded through explicit rule-based governance. All exclusions were documented to preserve traceability and ensure that removal decisions were transparent and reproducible.

Based on this governance process, three predefined feature-set variants were constructed: FULL, CLINICAL, and BIOMARKERS. The composition of each variant, including the number of governed input variables and their categorical breakdown, is summarized in Table S8. Feature-set definitions were explicitly frozen at the conclusion of Stage 1 to prevent any data-driven redefinition of feature availability assumptions in downstream modeling stages.

No outcome-based univariate screening, feature selection, imputation, scaling, model fitting, or threshold optimization was performed during Stage 1.

### 4.2. Stage 2 — Algorithm benchmarking using nested cross-validation

Algorithm benchmarking was conducted using a stratified 5×5 nested cross-validation framework to obtain unbiased estimates of predictive performance while structurally preventing information leakage during hyperparameter optimization. The prediction task was formulated as a binary classification problem distinguishing STEMI from NSTEMI, with STEMI treated as the positive class. Consistent with the overarching design of the case study, this task represents a diagnostic subtyping problem and was used as a methodological proxy endpoint, rather than as a clinically meaningful post-MI prognostic outcome. The outer cross-validation loop consisted of five stratified folds preserving outcome prevalence and was used exclusively for performance estimation. The inner loop, also comprising five folds, was used solely for model selection and hyperparameter tuning. No model was refitted on the full dataset for performance reporting.

All preprocessing steps - including imputation, scaling, and categorical encoding—were implemented within scikit-learn pipelines and fitted strictly within training folds to prevent cross-fold contamination. Multiple classification algorithms were benchmarked under identical validation conditions for each predefined feature-set variant. Feature-set definitions were inherited unchanged from Stage 1 and remained frozen throughout benchmarking.

Hyperparameter search spaces, random seeds, and software package versions were fixed a priori and recorded to ensure reproducibility. Out-of-fold (OOF) predictions aggregated across outer folds were treated as the single source of truth for all downstream analyses, including calibration, decision-analytic evaluation, and robustness diagnostics.

For each feature-set variant, a winner model was designated based on the highest mean outer-fold ROC-AUC. This designation serves exclusively as an analytical convenience and does not imply external validity or clinical readiness. Additional fold-level sanity checks were applied, including detection of fold-specific all-missing features, verification of feature availability, and inspection of potential outcome-proxy variables, particularly to ensure that no features implicitly encoded downstream clinical decision processes. Benchmarking results are summarized in Table S8. Fold-level variability in discrimination across outer folds is provided in Supplementary Figure S2.

### 4.3. Stage 3 — Consolidated OOF performance reporting and uncertainty quantification

All OOF predictions generated during Stage 2 were consolidated across outer folds and analyzed without refitting any model on the full dataset. This approach ensured that all reported performance estimates were based exclusively on predictions obtained from samples not used for model training within their respective folds.

To quantify performance uncertainty, stratified bootstrap resampling was conducted at the patient level, preserving class imbalance in each resample. Bootstrap distributions were constructed from OOF predictions, and percentile-based confidence intervals were derived for all primary performance metrics.

Discrimination and probabilistic accuracy were assessed using the ROC-AUC, area under the precision–recall curve (PR AUC) with explicit prevalence reference baselines, and the Brier score. All curve construction, metric estimation, and uncertainty quantification relied exclusively on OOF predictions to prevent optimistic bias and to preserve strict separation between model development and evaluation.

Given that the analytical task represents a diagnostic proxy classification problem (STEMI vs NSTEMI), these performance metrics are interpreted strictly as indicators of methodological behavior under controlled conditions, rather than as measures of clinically actionable prognostic performance.

Winner-model OOF performance estimates with corresponding 95% bootstrap confidence intervals are summarized in Table S9. Consolidated OOF ROC and precision–recall curves for the winner models are shown in Figure S4.

### 4.4. Probability calibration and decision-analytic evaluation

Probabilistic calibration and decision-analytic behavior were treated as integral components of the evaluation framework rather than secondary performance descriptors. All calibration analyses were based exclusively on OOF predictions generated during Stage 2, without refitting models on the full dataset.

Calibration quality was assessed using the Brier score, calibration slope and intercept, expected calibration error (ECE), maximum calibration error (MCE), and reliability curves constructed from aggregated OOF predictions. To prevent calibration-induced information leakage, probability recalibration methods were implemented using a cross-fitting strategy. Specifically, Platt scaling and isotonic regression models were trained on predictions from K−1 outer folds and applied exclusively to the held-out fold, ensuring strict separation between calibration fitting and evaluation.

Calibration behavior differed across feature-set variants. In the FULL variant, near-deterministic discrimination produced highly polarized predicted probabilities, resulting in sparsely populated reliability bins. Under such conditions, apparent alignment with the identity line must be interpreted cautiously, as calibration curves primarily reflect extreme probability separation rather than smooth risk stratification. In contrast, the CLINICAL and BIOMARKERS variants exhibited broader probability distributions, allowing more informative assessment of calibration properties. Cross-fitted isotonic regression provided the most stable recalibration behavior for these variants without introducing visible overfitting artifacts.

Decision thresholds were evaluated using the Youden index, F1 score, and Matthews correlation coefficient. Threshold optimization was performed within training folds and subsequently evaluated on held-out folds to preserve strict separation between selection and assessment. Decision-curve analysis (DCA) was conducted on aggregated OOF predictions to quantify net clinical benefit across a clinically plausible range of threshold probabilities.

Given that the underlying prediction task represents a diagnostic proxy classification problem rather than a prognostic endpoint, decision-analytic outputs (including DCA) are interpreted as methodological indicators of model behavior under varying threshold conditions, rather than as direct estimates of clinical utility in post-MI risk stratification.

Cross-fitted calibration metrics and threshold summaries are reported in Table S10. OOF reliability curves before and after cross-fitted recalibration are shown in Figure S4, and OOF-based decision-curve analysis is presented in Figure S6.

### 4.5. Performance–complexity trade-off and robustness checks

A structured evaluation of performance–complexity trade-offs was conducted by jointly considering discrimination, uncertainty, number of governed predictors, and decision-analytic behavior. This analysis aimed to contextualize predictive performance in relation to model dimensionality rather than to maximize a single performance metric. Summary metrics for the winner models across feature-set variants are provided in Table S12.

To assess structural robustness and model dependence on highly influential predictors, controlled ablation analyses were performed. Features were iteratively removed in descending order of outer-fold permutation importance, computed exclusively on held-out folds to prevent bias. At each ablation step, model performance was re-evaluated under fixed hyperparameters using the same nested cross-validation configuration. These analyses were conducted as diagnostic stress tests to evaluate performance degradation patterns and potential sensitivity to dominant predictors, and were not intended as feature selection procedures.

As a complementary robustness analysis, controlled model simplification was performed using regularization-based feature reduction within a fixed nested cross-validation framework. Feature reduction was applied within training folds only and without altering the predefined governance-based feature-set variants. Reduced-feature models were subsequently evaluated using the identical outer-fold, out-of-fold prediction protocol to determine whether performance–complexity relationships were preserved under constrained model capacity.

This dual robustness strategy - ablation-based stress testing and regularization-based simplification - was designed to probe methodological stability and structural reliance on predictor subsets rather than to optimize predictive performance.

Consistent with the proxy nature of the underlying classification task, these robustness analyses are interpreted as diagnostic assessments of model stability and feature dependence, rather than as evidence supporting clinically meaningful biomarker selection or prognostic relevance.

Performance–complexity summaries are reported in Table S13. Feature ablation trajectories across variants are shown in Figure S6. Results of regularization-based feature reduction are summarized in Table S14, with corresponding stability comparisons presented in Figure S7.

### 4.6. Locked-model diagnostic evaluation

To further interrogate the robustness of the methodological framework and to guard against implicit decision leakage, a locked-model diagnostic evaluation was conducted as a post-benchmarking stress test. This stage was designed to emulate a one-shot evaluation scenario in which all modeling decisions were finalized prior to exposure to evaluation data.

A single stratified train–test split was constructed and fixed a priori. The test partition remained fully isolated throughout this stage. Only samples assigned to the training partition were used for model fitting and hyperparameter optimization, while the held-out test set was accessed exactly once for final diagnostic evaluation.

For each predefined feature-set variant, the winner model architecture identified during nested cross-validation in Stage 2 was retained. Feature preprocessing pipelines, governance rules, and frozen feature-set definitions were inherited unchanged. Hyperparameter tuning was performed exclusively within the training partition, using the same modeling configuration and preprocessing structure as in Stage 2. No feature redefinition, recalibration, threshold re-optimization, or additional model selection steps were performed after exposure to test-set predictions.

The locked models were evaluated once on the held-out test set using the same primary performance metrics applied in earlier stages (ROC-AUC, PR AUC, and Brier score). This evaluation was explicitly framed as a diagnostic robustness check rather than as an estimate of deployable or externally valid performance. Accordingly, locked-split results complement - but do not replace - the out-of-fold estimates derived from nested cross-validation.

Consistent with the proxy nature of the underlying classification task, locked-model evaluation is interpreted as a workflow-level diagnostic of generalization behavior under strict isolation constraints, rather than as evidence of clinically meaningful predictive performance.

Consistency between nested cross-validation (OOF) performance patterns and locked-split behavior was assessed qualitatively across feature-set variants. Structural diagnostics of split integrity - including verification of feature availability, confirmation of unchanged governance-based feature lists, and documentation of any features dropped due to fold-specific constraints - are summarized in Table S15. Locked test-set performance metrics are reported in Table S16.

### 4.7. Diagnostic validation of the modeling workflow

To further interrogate the structural integrity of the modeling pipeline and detect potential methodological failure modes not captured by conventional performance metrics, two complementary workflow-level diagnostic procedures were implemented.

#### Permutation-based falsification testing

A permutation-based diagnostic evaluation was conducted using the identical nested 5×5 cross-validation configuration applied during algorithm benchmarking in Stage 2. This analysis was performed exclusively on the TRAIN-only subset defined in Stage 4.6, with the locked test partition excluded to prevent contamination of the diagnostic process. The governed feature lists, preprocessing pipelines, and hyperparameter search spaces were retained unchanged.

Outcome labels were randomly permuted 500 times while preserving the original feature matrix and fixed outer-fold splits. For each permutation, the full nested cross-validation procedure-including inner-loop hyperparameter optimization - was executed. Maintaining identical fold structure across true-label and permuted-label runs ensured direct comparability of performance distributions and eliminated partition-driven artifacts.

Permutation testing was implemented as a falsification procedure designed to determine whether the modeling pipeline could generate spurious discrimination in the absence of a genuine association between predictors and outcome labels.

#### Simplified-model capacity stress testing

Complementary workflow validation was performed using simplified model classes with reduced representational capacity. Linear and shallow baseline classifiers were evaluated under the same nested cross-validation protocol and governance-defined feature sets used in the primary benchmarking stage. Hyperparameter optimization remained restricted to inner folds, and feature definitions were not modified.

This diagnostic analysis aimed to assess whether trivial or low-capacity model architectures could achieve disproportionately high discrimination under the established pipeline. Instances of fold-level perfect discrimination, when present, were explicitly flagged and examined as potential diagnostic anomalies rather than being treated as confirmatory evidence.

Together, permutation-based falsification testing and simplified-model benchmarking were designed to evaluate the credibility and internal consistency of the modeling workflow itself rather than to optimize predictive performance or modify model-selection decisions.

Consistent with the proxy nature of the underlying classification task, these diagnostic procedures are interpreted as workflow-level validation of methodological integrity, and not as evidence of clinically meaningful predictive capability or prognostic validity.

Summary results of permutation-based null distributions and simplified-model benchmarking are reported in Table S17A, B, with detailed fold-level diagnostics provided in the Supplementary Materials.

## 5. Case study: methodological insights from the post–MI dataset

Application of the leakage-aware analytical workflow to the post–myocardial infarction dataset served as a methodological stress test of the proposed modeling framework rather than as an attempt to construct a clinically deployable prediction model. The comparative behavior of the FULL, CLINICAL, and BIOMARKERS feature-set variants provides insight into how feature governance, predictor dimensionality, and model capacity interact under strictly controlled validation conditions.

Across all variants, predictive performance estimates were derived exclusively from out-of-fold predictions obtained under the nested cross-validation protocol described in Section 5. Bootstrap resampling was used to estimate uncertainty intervals for discrimination metrics.

### 5.1 FULL variant: near-perfect discrimination as a leakage diagnostic signal

The FULL feature-set variant incorporated 69 governed predictors and achieved near-deterministic discrimination under nested cross-validation. Out-of-fold evaluation yielded ROC-AUC = 0.9988 (95% CI 0.9925–1.000), PR AUC = 0.9985, and Brier score = 0.0777.

Decision-curve analysis indicated high apparent net benefit across a broad range of threshold probabilities. However, subsequent robustness diagnostics contextualized this extremely high discrimination.

Ablation analyses revealed pronounced sensitivity of predictive performance to removal of top-ranked predictors, with rapid degradation of discrimination following iterative feature exclusion. Regularization-based simplification reduced model dimensionality while partially preserving discrimination, suggesting that predictive signal was concentrated within a restricted subset of variables.

Within the present leakage-aware framework, such near-perfect internal discrimination is interpreted as a diagnostic warning signal rather than evidence of clinical readiness. Extremely high separability under maximal feature inclusion may reflect proximity to downstream clinical decision processes or post-baseline information rather than true early predictive capability.

### 5.2 CLINICAL variant: conservative performance under realistic predictor constraints

The CLINICAL variant consisted of 58 governance-constrained clinical predictors. Under the same nested cross-validation framework, this configuration produced substantially lower discrimination than the FULL model.

Out-of-fold performance estimates were ROC-AUC = 0.6025 (95% CI 0.4463–0.7450), PR AUC = 0.5797, and Brier score = 0.2365.

Compared with the FULL configuration, predicted probabilities were more broadly distributed, enabling more stable estimation of calibration curves following cross-fitted recalibration. Robustness diagnostics indicated reduced sensitivity to removal of individual predictors, suggesting that predictive signal was not concentrated in a small number of dominant variables.

Within the methodological context of this analysis, the CLINICAL configuration represents a conservative baseline reflecting feature availability plausibly aligned with early clinical decision contexts. The modest discrimination observed under this configuration highlights the substantial gap between maximal feature inclusion and realistically available clinical predictors.

### 5.3 BIOMARKERS-only variant: translational signal under constrained dimensionality

The BIOMARKERS-only configuration contained 10 laboratory-derived predictors and achieved strong discrimination relative to its substantially reduced dimensionality.

Out-of-fold evaluation produced ROC-AUC = 0.9300 (95% CI 0.8537–0.9863), PR AUC = 0.8928, and Brier score = 0.1115.

Performance–complexity analysis indicated favorable trade-offs between discrimination and model dimensionality compared with the FULL configuration. Feature-ablation trajectories showed gradual rather than abrupt degradation of discrimination, suggesting distributed predictive contribution among multiple biomarkers rather than dependence on a single dominant variable.

Within the leakage-aware analytical framework, this configuration can therefore be interpreted as a hypothesis-generating translational signal, although external validation and temporal alignment analyses would be required before drawing any clinical conclusions.

### 5.4 Workflow diagnostics and methodological implications

Permutation-based falsification testing demonstrated that, under random label assignment, predictive discrimination collapsed to chance levels across all feature-set variants (mean permuted ROC-AUC ≈ 0.50). This result indicates that the observed predictive performance under true labels was not driven by technical leakage within the modeling pipeline or by deterministic preprocessing artifacts.

Complementary simplified-model benchmarking further confirmed that low-capacity model architectures did not systematically reproduce the high discrimination observed in the most expressive configurations.

Taken together, these diagnostic analyses support the internal validity of the modeling workflow while simultaneously illustrating the importance of feature governance and model complexity when interpreting discrimination metrics in biomarker-based machine-learning studies.

Overall, this proof-of-concept case study was designed not to define a clinically deployable model but to operationalize and empirically evaluate a leakage-aware machine-learning workflow under controlled conditions. By applying a fixed end-to-end validation, calibration, and reporting strategy across predefined feature-set variants, the analysis isolates the methodological consequences of feature composition while holding modeling strategy constant.

These findings highlight that discrimination metrics alone are insufficient to establish translational credibility. Instead, layered safeguards—including permutation diagnostics, simplified-model stress testing, and governance-based feature control—provide complementary protection against both data leakage and decision leakage in AI-driven biomarker research.

## 6. Toward a Methodological Framework for AI in Cardiac Biomarker Research

### 6.1 Lessons from the systematic literature review (PRISMA-derived gaps and needs)

The systematic literature review conducted in this study revealed a rapidly expanding body of research exploring AI methods applied to cardiac biomarkers in the context of myocardial infarction. However, despite the growing number of publications, the PRISMA-based analysis and thematic synthesis identified several recurring methodological limitations that may compromise the reliability, interpretability, and clinical applicability of many reported models.

These limitations were observed across multiple dimensions of the machine learning workflow, including feature selection, validation strategies, model evaluation, interpretability, and reporting transparency. Collectively, these challenges highlight the need for a more structured methodological framework guiding the development and evaluation of AI models in cardiac biomarker research.

The following subsections summarize the key methodological issues identified in the reviewed literature and outline the rationale for the proposed framework.

#### 6.1.1 Inadequate control of information leakage in biomarker studies

One of the most critical methodological issues identified in the reviewed studies is the insufficient control of information leakage during model development. Information leakage occurs when variables that are directly or indirectly related to the outcome are inadvertently incorporated into the predictive model in a way that artificially inflates its performance.

In biomarker-based studies, this issue often arises when biomarkers strongly associated with the clinical endpoint are included in the feature set without careful consideration of their temporal relationship to the outcome. For example, biomarkers measured after the onset of complications or near the time of outcome occurrence may contain information that would not be available in real-world prediction settings.

Furthermore, some studies combine heterogeneous groups of variables - such as routine clinical measurements, research biomarkers, and post-event laboratory markers - without explicitly defining feature governance or data partitioning rules. This practice increases the risk that the model may inadvertently exploit information that should not be available at the time of prediction.

The systematic review revealed that relatively few studies explicitly addressed the risk of information leakage or described strategies to mitigate it. Clear methodological safeguards - such as strict temporal separation of predictors and outcomes, feature governance frameworks, and transparent documentation of variable selection - were rarely reported.

Addressing this challenge is essential to ensure that AI models for cardiac biomarker analysis produce realistic performance estimates and remain applicable in prospective clinical settings.

#### 6.1.2 Overreliance on discrimination metrics without clinical utility assessment

Another common limitation identified in the reviewed literature is the strong emphasis on discrimination metrics, particularly the AUC-ROC, as the primary measure of model performance.

While discrimination metrics are useful for evaluating the ability of a model to distinguish between outcome classes, they do not provide sufficient information about the model’s clinical usefulness. A model may achieve high AUC values while still being poorly calibrated or offering limited benefit for clinical decision-making.

Only a minority of studies reported additional evaluation metrics such as calibration curves, Brier scores, or decision curve analysis. As a result, many models lack comprehensive evaluation of their potential impact on clinical workflows.

This finding reflects a broader trend in medical AI research, where predictive performance is often prioritized over clinical applicability. However, for models intended to support clinical decision-making, it is essential to assess not only whether predictions are accurate but also whether they lead to improved patient outcomes or better resource allocation.

Future studies should therefore incorporate a broader set of evaluation metrics, including calibration measures, clinical utility analysis, and decision curve analysis, to provide a more complete assessment of model performance.

#### 6.1.3 Limited interpretability and lack of stability analysis

Interpretability is a key requirement for the successful integration of AI models into clinical practice. Physicians must be able to understand the factors driving model predictions in order to trust and appropriately apply algorithmic recommendations.

Although several studies incorporated explainability techniques such as SHAP or feature importance analysis, interpretability was often treated as an optional add-on rather than a core methodological component. In many cases, the reported explanations were limited to global importance rankings without deeper analysis of model behavior across patient subgroups.

Another related issue is the lack of model stability analysis. Few studies evaluated the robustness of feature importance rankings or prediction outputs under different data perturbations, sampling strategies, or validation folds. Without such analyses, it is difficult to determine whether the identified predictors represent stable biological signals or artifacts of a specific dataset.

In the context of biomarker research, where biological variability and measurement noise are common, evaluating the stability of model predictions and feature importance is particularly important. Incorporating stability analysis and robust interpretability methods should therefore become a standard component of AI biomarker studies.

#### 6.1.4 Insufficient transparency, data governance, and reproducibility

The systematic review also revealed significant challenges related to transparency and reproducibility. Many studies provided limited methodological detail regarding data preprocessing, feature engineering, missing data handling, and model tuning procedures.

In addition, relatively few studies made their datasets or code publicly available, which restricts the ability of other researchers to replicate findings or validate proposed models. Lack of transparency can also obscure potential sources of bias or methodological weaknesses.

Another related issue is the absence of explicit data governance frameworks. In biomarker-based research, variables may originate from diverse sources, including clinical laboratory systems, research assays, and derived computational features. Without clear governance rules defining which variables belong to specific feature categories and how they are incorporated into models, the risk of methodological inconsistencies increases.

Adopting standardized reporting guidelines - such as TRIPOD-AI - and promoting open science practices could significantly improve the reproducibility and credibility of AI studies in cardiovascular biomarker research.

### 6.2. Lessons from the proof-of-concept case study (workflow-driven insights)

The proof-of-concept case study provides a set of workflow-driven methodological insights that extend beyond the specific dataset or prediction task examined. Rather than offering conclusions about the clinical utility of any individual model, the case study elucidates how methodological design choices fundamentally shape the interpretation, credibility, and translational plausibility of AI-based biomarker analyses after myocardial infarction.

#### 6.2.1. Leakage-aware validation fundamentally alters performance interpretation

The analyses demonstrate that validation strategies explicitly designed to detect and prevent information leakage materially influence how apparent model performance is interpreted. In the present dataset, near-deterministic discrimination emerged under maximal feature inclusion. However, when evaluated within a leakage-aware framework - including nested cross-validation, governance-constrained feature definitions, ablation stress testing, regularization-based simplification, locked-model diagnostic evaluation, and permutation-based falsification - exceptionally high discrimination functioned primarily as a structural warning signal rather than as direct evidence of deployable predictive accuracy.

Importantly, permutation testing showed convergence toward chance-level discrimination under label randomization, indicating that the observed performance was not driven by technical leakage within the validation procedure itself. Similarly, simplified low-capacity models did not systematically reproduce near-perfect discrimination. These results suggest that high internal separability can coexist with methodological integrity, while still warranting careful scrutiny of feature provenance and temporal alignment. Leakage-aware validation therefore reframes performance interpretation from a pursuit of maximal discrimination toward a structured assessment of credibility, robustness, and contextual plausibility.

#### 6.2.2. Calibration and decision-curve analysis reveal clinically meaningful trade-offs

The case study highlights that discrimination metrics alone provide an incomplete basis for translational interpretation. Calibration analyses demonstrated variant-dependent probability behavior: models with similar discrimination exhibited differing reliability characteristics, particularly under cross-fitted recalibration. In the FULL configuration, probability concentration limited the interpretability of reliability curves despite high discrimination, whereas CLINICAL and BIOMARKERS variants exhibited more informative calibration profiles.

Decision-curve analysis further showed that discrimination gains do not uniformly translate into consistent net benefit across plausible decision thresholds. Joint evaluation of calibration behavior and decision-analytic performance exposed trade-offs between dimensional richness, probability stability, and threshold-dependent utility that are not visible when performance is summarized solely by ROC-AUC. These findings reinforce the necessity of integrating calibration and decision-analytic evaluation as core components of AI-based biomarker research.

#### 6.2.3. Interpretability as stability and consistency, not explanation

The proof-of-concept analyses support a reframing of interpretability away from mechanistic explanation toward stability- and consistency-based criteria. Ablation trajectories revealed that, in the FULL configuration, predictive performance was highly sensitive to removal of top-ranked features, indicating structural concentration of signal. In contrast, the BIOMARKERS variant demonstrated more gradual degradation under feature removal, consistent with distributed contribution rather than single-variable dominance.

Regularization-based simplification further illustrated that interpretability claims are most credible when performance remains relatively stable under dimensional constraint. Within a leakage-aware workflow, interpretability is therefore best conceptualized as evidence of reproducible, governance-consistent signal across folds, perturbation strategies, and modeling constraints, rather than as post hoc explanation of isolated feature importance rankings.

#### 6.2.4. Feature-set variants as an explicit design dimension

By evaluating predefined feature-set variants under identical validation, calibration, and reporting conditions, the case study demonstrates that feature availability assumptions constitute a critical design dimension in AI-based biomarker research. Observed differences in discrimination, calibration stability, robustness under perturbation, and performance–complexity trade-offs were attributable to feature composition rather than to variation in modeling strategy.

Treating feature-set definition as an explicit, auditable modeling decision - frozen prior to model benchmarking - enabled transparent comparison of structural behavior across configurations. This approach prevents overinterpretation of performance derived from unrealistically expansive feature spaces and promotes a more contextually grounded interpretation of AI-based biomarker models.

### 6.3. Toward a unified methodological framework

Building on these insights, the proof-of-concept case study motivates a unified methodological framework for AI-based biomarker research that prioritizes credibility, transparency, and translational plausibility over isolated performance optimization.

#### 6.3.1. Core principles for AI-based biomarker research

Several core principles emerge from the workflow-driven analyses. First, validation must be explicitly leakage-aware, incorporating governance-constrained feature definitions, nested cross-validation with strict separation of model selection and evaluation, and structured diagnostic procedures such as ablation, simplification, locked-split testing, and permutation-based falsification. Second, evaluation should extend beyond discrimination to include calibration behavior, decision-analytic performance, and structural robustness under perturbation. Third, interpretability should be grounded in reproducibility and stability across analytical conditions rather than in post hoc explanatory narratives. Finally, analytical workflows should be version-controlled, auditable, and fully reproducible.

#### 6.3.2. A minimal reporting checklist for future studies

To operationalize these principles, future AI-based biomarker studies would benefit from a minimal reporting checklist including: explicit definition and justification of feature availability assumptions; description of leakage-prevention strategies; separation of model selection and final evaluation; reporting of calibration and decision-curve analyses; documentation of robustness and falsification diagnostics; and provision of reproducible code and intermediate artifacts. Such reporting practices facilitate critical appraisal, replication, and meaningful comparison across studies.

#### 6.3.3. Alignment with FAIR data and reproducibility standards

The proposed framework aligns naturally with FAIR data principles and contemporary reproducibility standards. Explicit feature governance, structured metadata, version-controlled code, and auditable intermediate outputs enhance transparency and reusability of both data and analytical workflows. Embedding AI-based biomarker research within such standards strengthens methodological rigor and supports cumulative scientific progress.

### 6.4. Positioning within cardiovascular AI research

Within the expanding landscape of cardiovascular AI research, the present work contributes a methodological perspective that complements algorithmic innovation. Rather than proposing new modeling techniques or clinical prediction tools, this study emphasizes the conditions under which performance claims can be interpreted as credible and methodologically defensible.

By demonstrating how apparent success may reflect structural characteristics of feature inclusion - and how layered validation, calibration, robustness, and falsification strategies can contextualize these signals—the study addresses recurring concerns in cardiovascular AI regarding overfitting, leakage, and translational fragility. The proposed workflow is therefore not a prescriptive modeling recipe, but a structured methodological scaffold adaptable to diverse cardiovascular biomarker applications while maintaining rigor, transparency, and interpretative restraint.

## 7. Discussion

The present study addresses a central and persistent challenge in cardiovascular AI research: the tension between impressive internal model performance and credible, clinically interpretable inference. By combining a systematic literature review with a rigorously implemented proof-of-concept case study, this work demonstrates that methodological design choices - particularly those related to validation strategy, feature governance, calibration, and robustness diagnostics - substantially shape how model performance should be interpreted.

Importantly, the proof-of-concept case study was intentionally designed as a methodological demonstration rather than a clinically oriented prognostic modeling exercise. The analytical task - binary classification of STEMI vs NSTEMI - represents a diagnostic subtyping problem and was deliberately used as a proxy endpoint to enable controlled evaluation of the modeling workflow under realistic data conditions. This design choice allows isolation of methodological effects, including leakage susceptibility and feature-dependence structure, while avoiding overinterpretation of results as clinically actionable post-MI risk prediction.

The proof-of-concept analyses illustrate that near-perfect discrimination can emerge under internally valid, nested cross-validation settings when expansive feature configurations are used. In the FULL variant, discrimination approached deterministic separation under out-of-fold evaluation. However, complementary diagnostics - including feature ablation, regularization-based simplification, locked-split evaluation, and permutation-based falsification - contextualized this performance pattern. Permutation testing led to collapse of discrimination toward chance levels, indicating absence of technical leakage within the validation pipeline itself. Simplified low-capacity models did not systematically reproduce extreme performance, and locked-split evaluation qualitatively preserved relative performance ordering across feature-set variants. Together, these findings suggest that exceptionally high discrimination may reflect structural feature composition rather than procedural validation flaws, and therefore requires scrutiny of feature provenance and temporal plausibility rather than automatic acceptance.

A central contribution of this work lies in demonstrating how calibration and decision-analytic evaluation materially enrich performance interpretation. Although discrimination differed across feature-set variants, calibration behavior and net benefit profiles exhibited additional structural differences. In the FULL configuration, high discrimination coincided with probability concentration and limited calibration granularity, whereas CLINICAL and BIOMARKERS variants displayed broader probability distributions and more interpretable reliability curves under cross-fitted recalibration. Decision-curve analysis further revealed that discrimination gains did not translate uniformly into stable net benefit across clinically plausible threshold ranges. Within the context of a proxy classification task, these findings highlight methodological properties of model behavior rather than direct estimates of clinical utility, underscoring that discrimination alone is insufficient for assessing translational relevance.

The study also advances a stability-centered perspective on interpretability. Rather than relying on isolated feature importance rankings, the analyses examined how performance behaved under structured perturbations. In the FULL variant, performance degradation following removal of top-ranked predictors indicated concentration of predictive signal within a narrow subset of variables.

In contrast, the BIOMARKERS configuration demonstrated more gradual degradation under ablation, consistent with distributed contribution. Regularization-based feature reduction partially preserved performance while reducing dimensionality, further illustrating that interpretability is most credible when predictive behavior remains structurally consistent under constraint. Within a leakage-aware framework, interpretability is therefore best conceptualized as stability across folds, perturbation strategies, and modeling constraints rather than as post hoc explanation. Importantly, these findings should not be interpreted as evidence for clinically validated biomarker importance but rather as indicators of structural model behavior.

Importantly, the study reframes feature availability as an explicit methodological dimension. By freezing and comparing predefined feature-set variants under identical validation, calibration, and reporting conditions, the analyses isolated the structural consequences of feature composition. Observed performance differences across variants reflected not only predictive signal but also implicit assumptions about timing, accessibility, and workflow integration of included variables. Making these assumptions explicit prevents overinterpretation of models derived from richly instrumented feature spaces that may not correspond to realistic deployment scenarios.

A key limitation of the present study is that the case study does not evaluate clinically meaningful post-MI prognostic outcomes, such as mortality, heart failure, or rehospitalization. Instead, the use of a diagnostic proxy endpoint was intended to support controlled methodological investigation. While this design strengthens internal validity of workflow evaluation, it limits direct clinical interpretability and generalizability of the empirical results. Future studies should therefore extend the proposed framework to true prognostic endpoints in externally validated and temporally structured datasets.

Within the broader cardiovascular AI landscape, this study contributes a methodological complement to algorithm-driven innovation. Rather than proposing a new predictive instrument, it offers a structured workflow for interrogating the credibility of internally derived performance claims. As AI applications move closer to clinical translation, such structured, leakage-aware, and stability-centered validation frameworks are essential for distinguishing robust signal from structural artifact and for supporting reproducible, trustworthy cardiovascular AI research.

## 8. Conclusions and Future Directions

This study demonstrates that the credibility and interpretability of AI-based cardiac biomarker models depend as much on methodological discipline as on modeling technique. Through a leakage-aware proof-of-concept case study, we show that exceptionally strong internal discrimination can arise under formally valid nested cross-validation, yet still require structural scrutiny with respect to feature composition, calibration behavior, and robustness under perturbation. High discrimination, in isolation, does not establish clinical readiness; it must be interpreted within a broader framework that evaluates calibration stability, decision-analytic behavior, feature dependence, and falsification diagnostics.

The proposed workflow integrates explicit feature governance, nested leakage-aware validation, cross-fitted calibration, structured robustness testing (including ablation and regularization-based simplification), locked-split diagnostic evaluation, and permutation-based falsification into a coherent and reproducible analytical pipeline. The empirical results demonstrate that combining these layers enables differentiation between structurally concentrated signal, distributed predictive contribution, and purely artifactual performance patterns. Importantly, permutation diagnostics confirmed collapse of performance under label randomization, and locked-split evaluation qualitatively preserved relative variant ordering, supporting procedural integrity of the validation framework.

The primary contribution of this work therefore lies not in maximizing predictive performance, but in advancing methodological standards for cardiovascular AI research. By treating feature availability, temporal plausibility, calibration reliability, and robustness under constraint as core evaluation dimensions, the study shifts emphasis from isolated performance metrics toward defensible and transparent inference.

Future research should extend these principles to external, prospective, and multi-center datasets, where independent validation can further clarify the relationship between internal structural robustness and real-world performance. Incorporating temporally explicit modeling frameworks, standardized biomarker ontologies, and harmonized clinical metadata may enhance interpretability while preserving governance constraints. Importantly, future investigations should continue to treat feature availability, timing, and clinical workflow integration as explicit design dimensions rather than implicit assumptions.

More broadly, adoption of leakage-aware, transparency-centered validation workflows may help bridge the gap between promising algorithmic performance and clinically meaningful translation. By embedding methodological rigor, auditability, and structured diagnostic safeguards into AI-based biomarker research, the field can progress toward models that are not only performant, but also reproducible, trustworthy, and contextually appropriate for cardiovascular decision-making.

## Data Availability

The code used for data preprocessing, feature engineering, model development, and evaluation in the proof-of-concept study is publicly available in a GitHub repository at: https://github.com/npiorkowska-science/post-mi-biomarkers-ml-review-case-study
The repository contains the scripts required to reproduce the machine learning pipeline, including data preprocessing procedures, model training routines, and evaluation workflows.
Due to privacy regulations and institutional data protection policies, the original clinical dataset cannot be publicly shared. However, the repository includes detailed documentation describing the data structure and variables required to reproduce the analysis using compatible datasets.

## Acknowledgments

The authors acknowledge the Wroclaw Medical University for providing access to the clinical data used in the proof-of-concept analysis. The authors also thank collaborators who contributed to the development of the methodological framework and provided valuable feedback during the preparation of this study.

## Author Contributions

Natalia Piórkowska (NP): Conceptualization, Methodology, Software, Formal analysis, Data curation, Investigation, Writing – original draft, Writing – review & editing.

Lech Madeyski (LM): Conceptualization, Methodology, Software, Validation, Supervision, Writing – original draft, Writing – review & editing.

Aleksandra Żyłka (AŻ): Visualization, Writing – review & editing. Aleksandra Musz (AMu): Visualization, Writing – review & editing.

Agnieszka Olejnik (AO): Methodology (clinical dataset design and biomarker panel selection), Data curation, Investigation, Validation, Writing – original draft, Writing – review & editing.

Wiktor Kuliczkowski (WK): The acquisition and curation of patient clinical and biomarker data. Andrzej Mysiak (AMy): The acquisition and curation of patient clinical and biomarker data.

Iwona Bil-Lula (IBL): Methodology (clinical dataset design and biomarker panel selection), Data curation, Investigation, Writing – original draft, Writing – review & editing.

All authors approved the final version of the manuscript and agreed to be accountable for all aspects of the work.

## Ethics Statement

The study was conducted in accordance with the Declaration of Helsinki. The use of clinical data for the proof-of-concept analysis was approved by the Ethics Committee of the Medical University of Wroclaw (approval numbers: KB-54/2019, KB-514/2019, KB-387/2021). All data used in the analysis were anonymized prior to processing. The systematic review part of the study was conducted using previously published studies and did not require additional ethical approval. The requirement for informed consent was waived due to the retrospective nature of the study and the use of anonymized data.

## Data and Code Availability

The code used for data preprocessing, feature engineering, model development, and evaluation in the proof-of-concept study is publicly available in a GitHub repository at: https://github.com/npiorkowska-science/post-mi-biomarkers-ml-review-case-study

The repository contains the scripts required to reproduce the machine learning pipeline, including data preprocessing procedures, model training routines, and evaluation workflows.

Due to privacy regulations and institutional data protection policies, the original clinical dataset cannot be publicly shared. However, the repository includes detailed documentation describing the data structure and variables required to reproduce the analysis using compatible datasets.

## Supplementary Materials

### Supplementary Tables

**Table S1.** Eligibility criteria applied in study selection

**Table S2.** Detailed search strategies used in each database

**Table S3.** Full list of all studies included in the systematic review (n = 120), including bibliographic details, country/centre, and primary study objective.

**Table S4.** Summary of risk of bias assessment using PROBAST

**Table S5.** Certainty of evidence assessment using a modified GRADE framework

**Table S6.** Data harmonization and type-coercion audit log

**Table S7.** Dataset structure and missingness profile after governance preprocessing

**Table S8.** Feature-set variant composition and governance summary **Table S9.** Nested cross-validation performance summary (OOF-based) **Table S10.** Winner model selection by feature-set variant

**Table S11.** Winner-model OOF performance with bootstrap confidence intervals

**Table S12.** Cross-fitted calibration and threshold-selection summary

**Table S13.** Performance–complexity summary for winner models (variant with net_benefit)

**Table S14.** Regularization-based feature reduction summary

**Table S15.** Locked-model split diagnostics and feature integrity checks

**Table S16.** Locked-model diagnostic test performance

**Table S17A.** Workflow-level diagnostic checks using permutation testing and simple models - Permutation diagnostic

**Table S17B.** Workflow-level diagnostic checks using permutation testing and simple models - Simplified models

### Supplementary Figures

**Figure S1.** Overview of the leakage-aware modeling workflow.

**Figure S2.** Distribution of outer-fold ROC-AUC across feature-set variants

**Figure S3.** OOF ROC and precision–recall curves for winner models

**Figure S4.** OOF reliability curves after cross-fitted calibration

**Figure S5.** OOF-based decision-curve analysis

**Figure S6.** Performance–complexity trade-off across feature-set variants.

**Figure S7.** Feature ablation curves across variants

