## Supplementary figures and images for "Artificial Intelligence for Cardiac Biomarkers After Myocardial Infarction: A Systematic Review and a Leakage-Aware Modeling Framework"

### Figure S1

**Figure S1.** Overview of the leakage-aware modeling workflow.
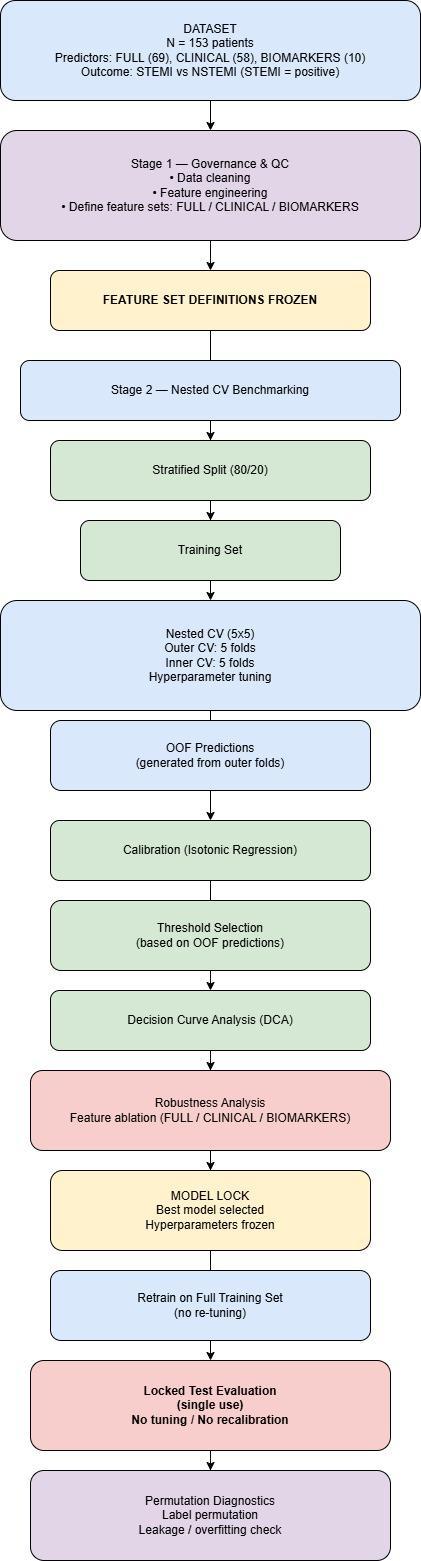

### Figure S2

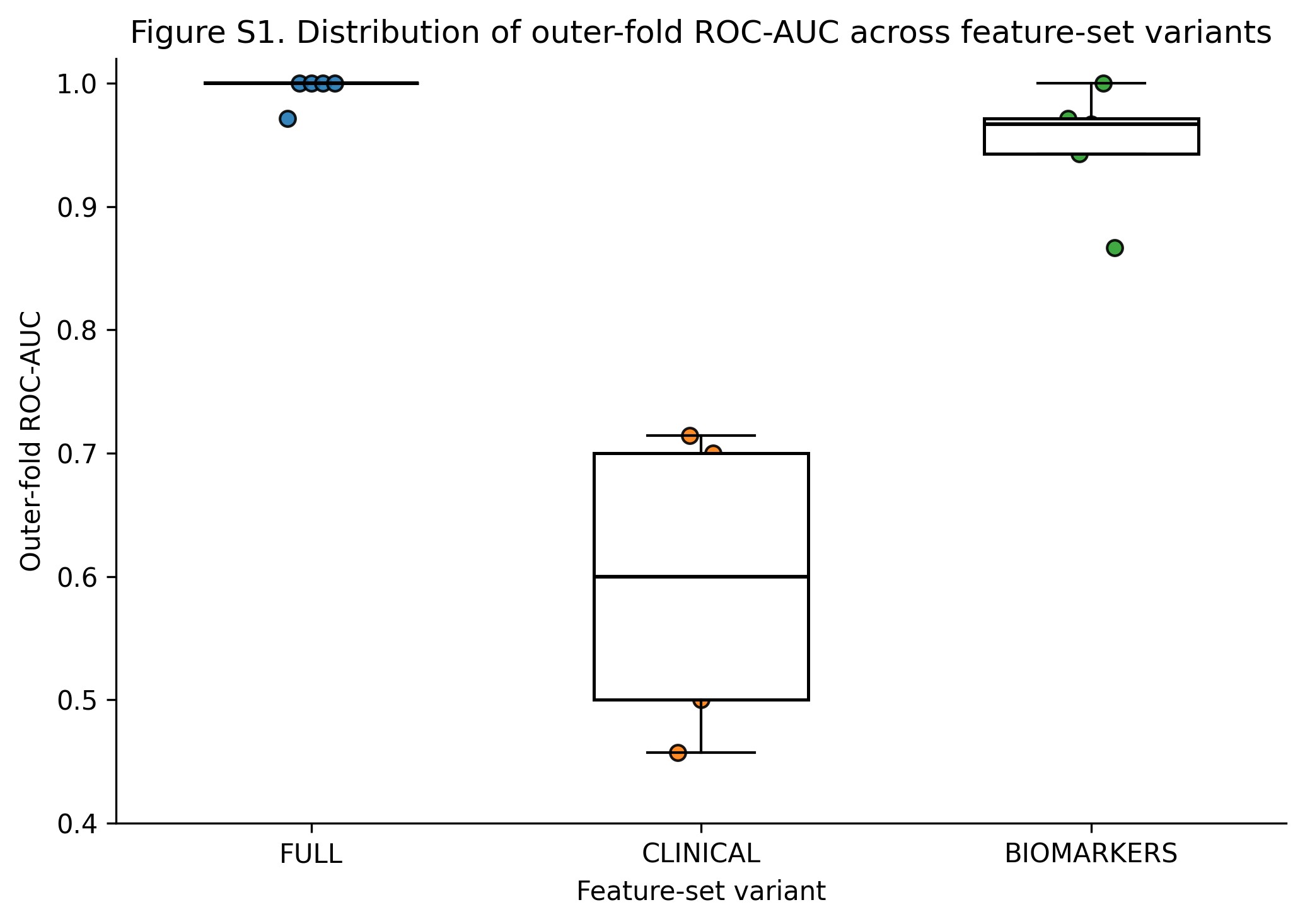

### Figure S3

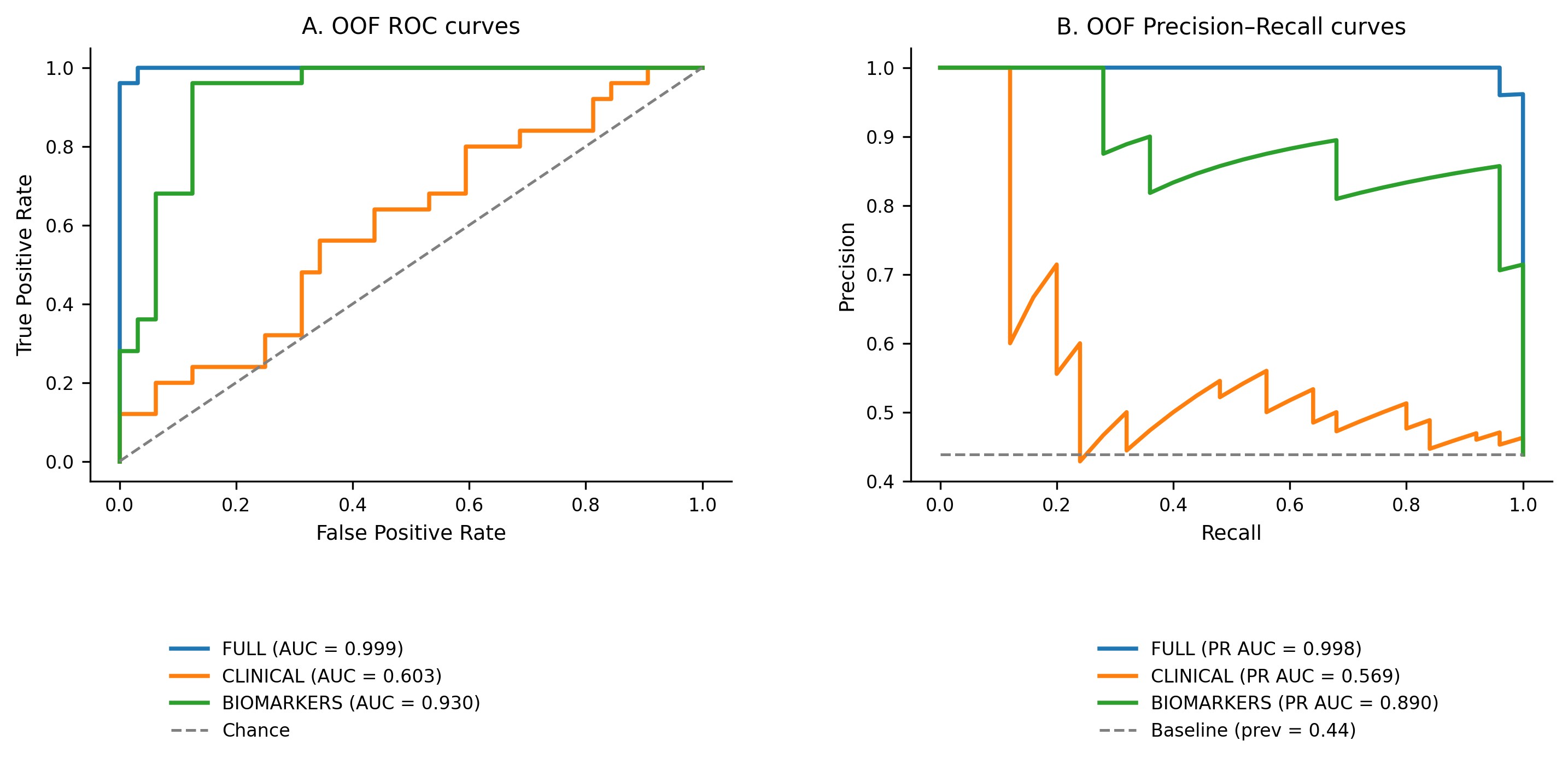

### Figure S4

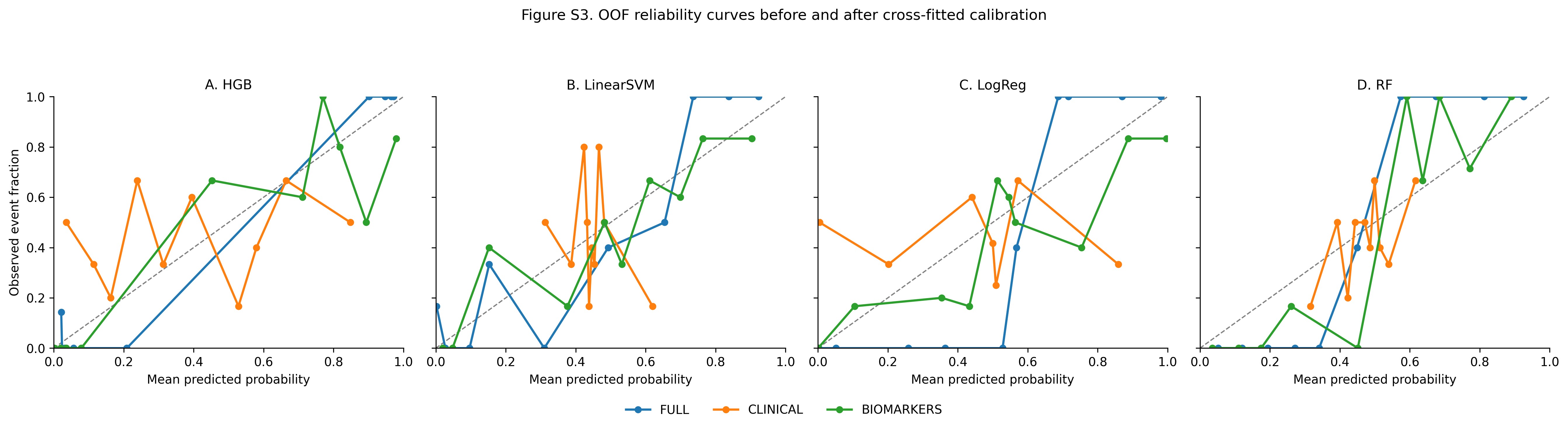

### Figure S5

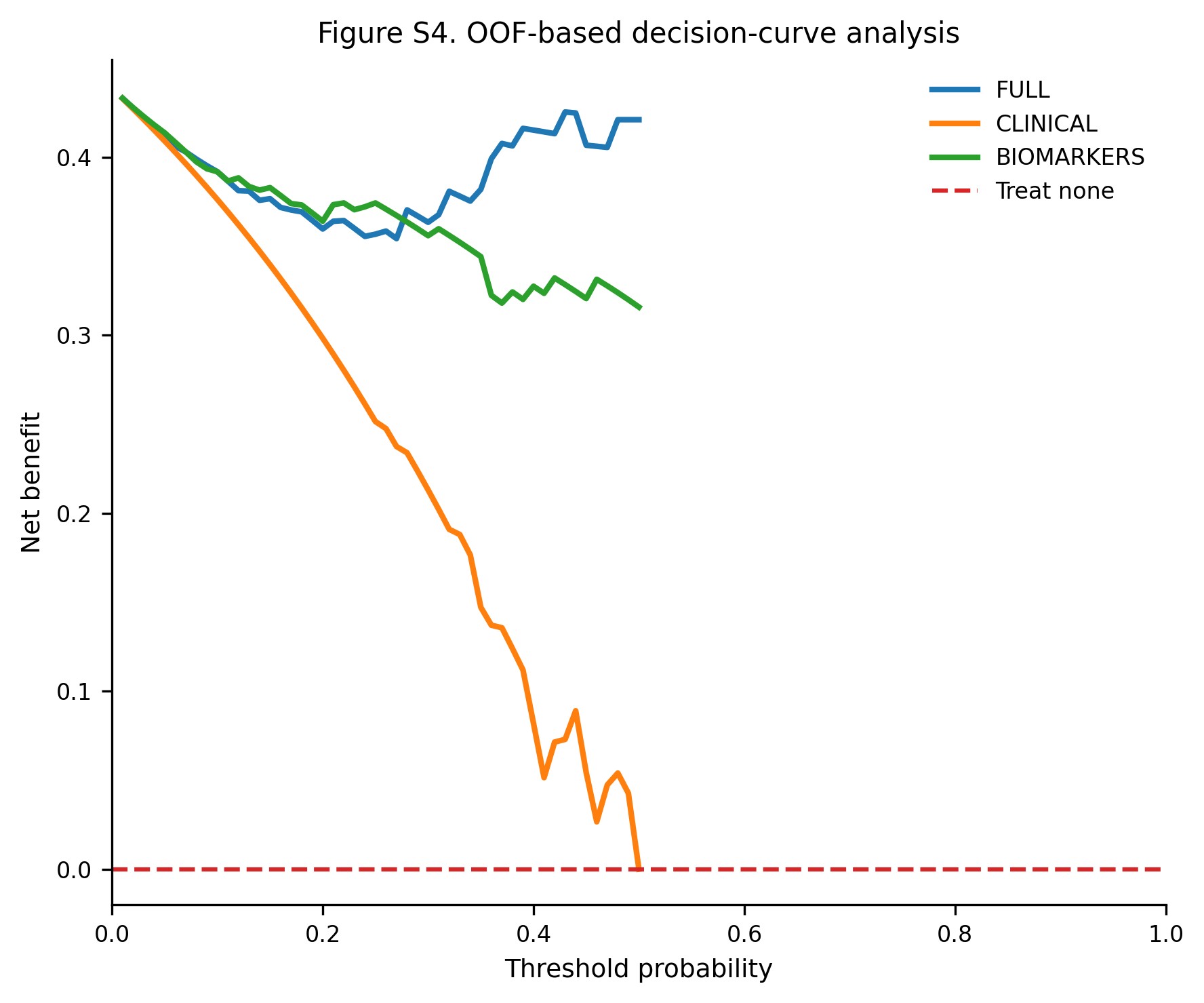

### Figure S6

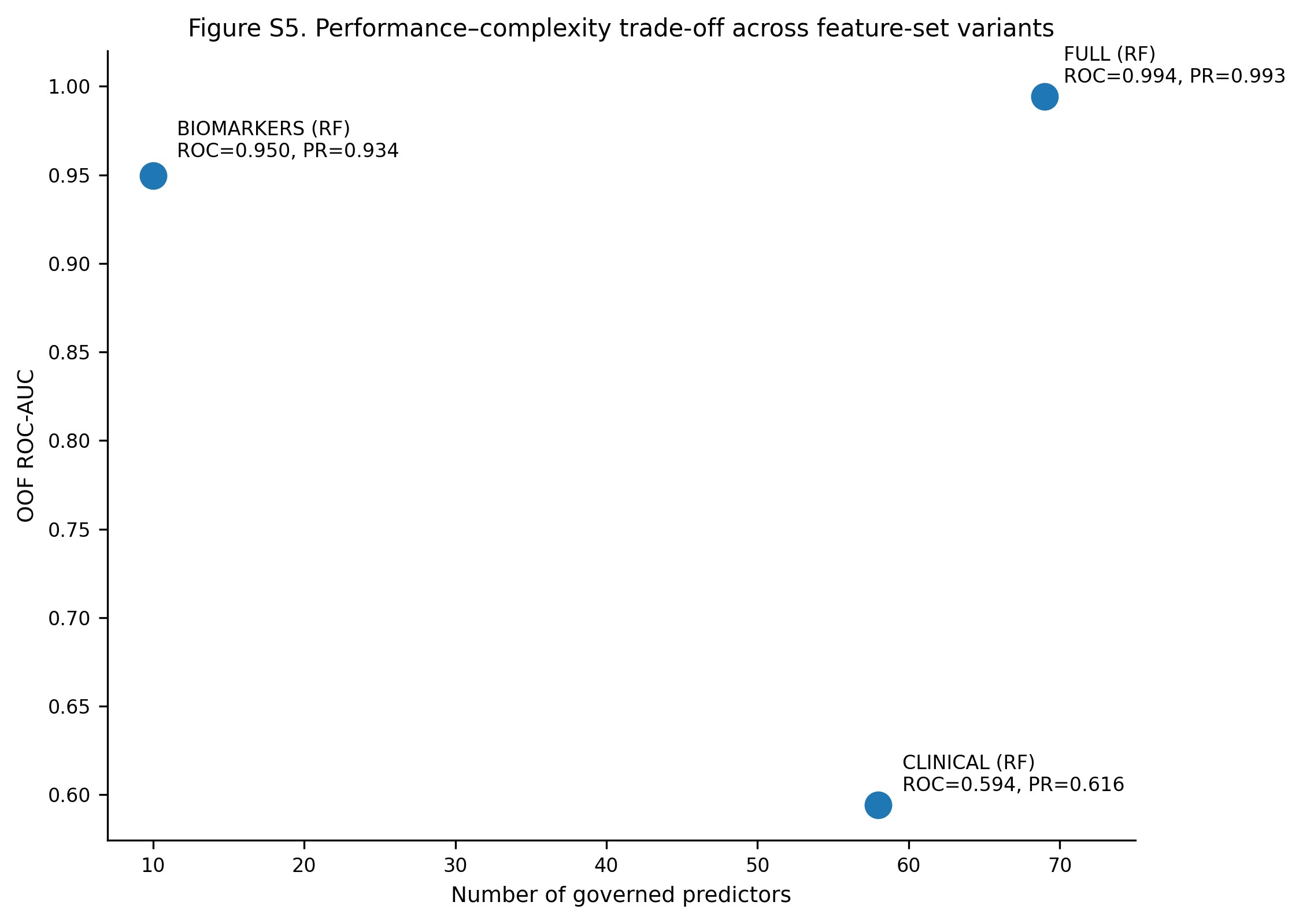

### Figure S7

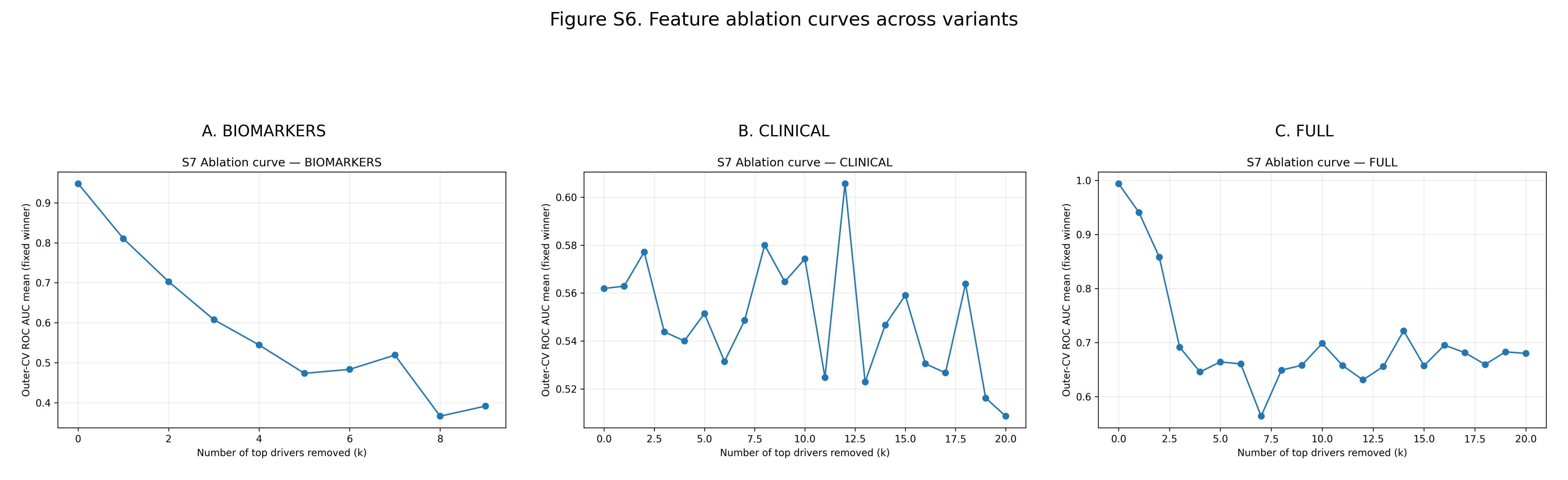
