## Supplementary material for "Artificial Intelligence for Cardiac Biomarkers After Myocardial Infarction: A Systematic Review and a Leakage-Aware Modeling Framework": Table S1

**Supplementary Table S1.** Eligibility criteria applied in study selection

| **Category** | **Criteria** |
| --- | --- |
| **Inclusion criteria** | |
| Population | Adult patients with a history of myocardial infarction (MI) |
| Intervention / Exposure | Application of artificial intelligence (AI) or machine learning (ML) algorithms |
| Data type | Analysis of cardiac biomarkers (biochemical, functional, or clinical) |
| Outcomes | Reporting of model performance and validation metrics |
| Study design | Original empirical studies (retrospective, prospective, or registry-based) |
| **Exclusion criteria** | |
| Study focus | Studies focused exclusively on acute MI diagnosis without predictive modeling |
| Methodology | Lack of identifiable AI/ML analytical component |
| Publication type | Reviews, editorials, commentaries, or conference abstracts without full methodological data |
| Model type | Studies involving animal or in vitro models |
| Reporting quality | Insufficient methodological detail upon full-text assessment |
| **Additional criteria** | |
| Language | English only |
| Publication type | Full-text articles |
| Time frame | 1 January 2015 – 30 September 2025 |
| Preprints | Included if complete methodological reporting was available; analyzed separately in sensitivity analyses |
