## Supplementary material for "Artificial Intelligence for Cardiac Biomarkers After Myocardial Infarction: A Systematic Review and a Leakage-Aware Modeling Framework": Table S2

**Supplementary Table S2.** Detailed search strategies used in each database

| **Database** | **Search strategy** |
| --- | --- |
| **PubMed/MEDLINE** | (("myocardial infarction"[MeSH Terms] OR "acute coronary syndrome"[MeSH Terms] OR "heart attack"[Title/Abstract] OR "post-MI"[Title/Abstract]) AND ("biomarkers"[MeSH Terms] OR biomarker*[Title/Abstract] OR "troponin"[Title/Abstract] OR "NT-proBNP"[Title/Abstract] OR "CRP"[Title/Abstract] OR "LVEF"[Title/Abstract]) AND ("artificial intelligence"[MeSH Terms] OR "machine learning"[MeSH Terms] OR "deep learning"[Title/Abstract] OR "neural networks"[Title/Abstract] OR "data mining"[Title/Abstract])) AND ("2010/01/01"[Date - Publication] : "2025/03/31"[Date - Publication]) AND English[lang] |
| **Scopus** | (TITLE-ABS-KEY("myocardial infarction" OR "acute coronary syndrome" OR "heart attack" OR "post-MI") AND TITLE-ABS-KEY("biomarker*" OR "troponin" OR "NT-proBNP" OR "CRP" OR "LVEF") AND TITLE-ABS-KEY("artificial intelligence" OR "machine learning" OR "deep learning" OR "neural network*" OR "data mining")) AND PUBYEAR > 2009 AND PUBYEAR < 2026 AND (LIMIT-TO(LANGUAGE, "English")) |
| **Web of Science Core Collection** | TS=("myocardial infarction" OR "heart attack" OR "acute coronary syndrome" OR STEMI OR NSTEMI) AND TS=("biomarker*" OR "troponin*" OR BNP OR "NT-proBNP" OR "CK-MB" OR "cardiac marker*" OR "inflammatory marker*" OR "metabolomic*" OR "proteomic*" OR "echocardiograph*" OR "cardiac MRI" OR strain OR "ejection fraction") AND TS=("artificial intelligence" OR "machine learning" OR "deep learning" OR "neural network*" OR "support vector machine*" OR "random forest*" OR "logistic regression" OR "gradient boosting" OR XGBoost OR AutoML) AND TS=("prediction model*" OR "risk prediction" OR "prognostic model*" OR validation OR "cross-validation" OR "external validation" OR "model calibration" OR "model performance") AND TS=("explainable AI" OR interpretability OR SHAP OR LIME OR "feature importance" OR "sensitivity analysis" OR ROC OR AUC OR "calibration curve" OR "decision curve analysis") AND PY=(2015–2025) |
| **IEEE Xplore** | ("myocardial infarction" OR "acute coronary syndrome" OR "heart attack") AND (biomarker* OR troponin OR NT-proBNP OR CRP OR LVEF) AND ("artificial intelligence" OR "machine learning" OR "deep learning" OR "neural network" OR "data mining") |
| **ACM Digital Library** | Abstract:("myocardial infarction" OR "acute coronary syndrome" OR "heart attack" OR "post-MI") AND Abstract:("biomarker*" OR "troponin" OR "NT-proBNP" OR "CRP" OR "LVEF") AND Abstract:("artificial intelligence" OR "machine learning" OR "deep learning" OR "neural network*" OR "data mining") AND Publication Date: (2010 TO 2025) AND Language: English |
| **Google Scholar (screening)** | (("myocardial infarction" OR "acute coronary syndrome" OR "heart attack" OR "post-MI") AND ("biomarker*" OR "troponin" OR "NT-proBNP" OR "CRP" OR "LVEF") AND ("artificial intelligence" OR "machine learning" OR "deep learning" OR "neural network*" OR "data mining")); first 200 results screened; publication years 2010–2025; English language |
