## Supplementary material for "Artificial Intelligence for Cardiac Biomarkers After Myocardial Infarction: A Systematic Review and a Leakage-Aware Modeling Framework": Table S3

**Supplementary Table S3.** Full list of all studies included in the systematic review (n = 120), including bibliographic details, country/centre, and primary study objective.

| **No.** | **Authors / Study** | **Year** | **Country / Centre** | **Primary study objective** |
| --- | --- | --- | --- | --- |
| 1 | Sladojević M, Čanković M, Čemerlić S, Mihajlović B, Ađić F, Jaraković M. Data mining approach for in-hospital treatment outcome in patients with acute coronary syndrome | 2015 | Serbia – Institute of Cardiovascular Diseases of Vojvodina, Sremska Kamenica; in collaboration with the University of Novi Sad, Faculty of Medicine | To develop a predictive model of in-hospital mortality in patients with ACS undergoing PCI using data-mining / machine-learning methods, including algorithm selection and validation. |
| 2 | Baars T, Neumann U, Jinawy M, Hendricks S, Sowa J-P, Kälfsch J, et al. In Acute Myocardial Infarction Liver Parameters Are Associated with Stenosis Diameter | 2016 | Germany – University Hospital Essen (West German Heart and Vascular Centre; Department of Gastroenterology and Hepatology); University of Applied Sciences Weihenstephan-Triesdorf | To assess whether non-invasive parameters, especially liver enzymes, can predict stenosis diameter in AMI patients referred for coronary angiography / PCI and to develop a predictive model based on the most informative features. |
| 3 | Sun C, Jiang H, Sun Z, Gui Y, Xia H. Identification of long non-coding RNAs biomarkers for early diagnosis of myocardial infarction from the dysregulated coding-non-coding co-expression network | 2016 | China – The Fourth Affiliated Hospital of Harbin Medical University; The Affiliated Hongqi Hospital of Mudanjiang Medical University | To identify an lncRNA panel for early AMI diagnosis, construct an lncRNA–mRNA co-expression network, and evaluate classification performance using SVM in discovery and independent validation cohorts. |
| 4 | Vignoli A, Tenori L, Giusti B, Takis PG, Valente S, Carrabba N, et al. NMR-based metabolomics identifies patients at high risk of death within two years after acute myocardial infarction in the AMI-Florence II cohort | 2019 | Italy – University of Florence, Careggi Hospital, CIRMMP, CERM, Giotto Biotech, ASL 10 Florence | To develop a metabolomics-based model for identifying patients at high risk of death within 2 years after AMI and to assess the prognostic value of NMR metabolomics after PCI. |
| 5 | Trainor PJ, Yampolskiy RV, DeFilippis AP. Wisdom of artificial crowds feature selection in untargeted metabolomics: An application to the development of a blood-based diagnostic test for thrombotic myocardial infarction | 2018 | USA – University of Louisville, Department of Medicine and Department of Computer Science and Engineering | To develop the Wisdom of Artificial Crowds feature-selection approach for untargeted metabolomics and apply it to a blood-based diagnostic test differentiating thrombotic MI, non-thrombotic MI, and stable CAD. |
| 6 | Steinhoff G, Nesteruk J, Wolfien M, Kundt G, Börgermann J, David R, et al. Cardiac Function Improvement and Bone Marrow Response – Outcome Analysis of the Randomized PERFECT Phase III Clinical Trial of Intramyocardial CD133+ Application After Myocardial Infarction | 2017 | Germany – multicentre: University Medicine Rostock, Hannover Medical School, University of Leipzig, University of Lübeck, University of Hamburg, Charité Berlin, University of Göttingen | To assess the safety and efficacy of intramyocardial autologous CD133+ cell therapy after MI and to identify molecular and clinical factors associated with improvement in LVEF and left ventricular remodelling. |
| 7 | Li L, Cong Y, Gao X, Wang Y, Lin P. Differential expression profiles of long non-coding RNAs as potential biomarkers for the early diagnosis of acute myocardial infarction | 2017 | China – Harbin Medical University and Second Affiliated Hospital; collaboration with the University of Calgary, Canada | To identify lncRNAs as biomarkers for early AMI diagnosis and build an SVM classifier based on 11 lncRNAs with internal and external validation. |
| 8 | Kwon J-M, Kim K-H, Jeon K-H, Park J. Deep learning for predicting in-hospital mortality among heart disease patients based on echocardiography | 2019 | South Korea – Mediplex Sejong Hospital and Sejong General Hospital, Incheon | To develop and validate a deep-learning model predicting in-hospital mortality in heart disease patients from echocardiography reports alone, without laboratory data. |
| 9 | Htun NM, Magliano DJ, Zhang Z-Y, Lyons J, Petit T, Nkuipou-Kenfack E, et al. Prediction of acute coronary syndromes by urinary proteome analysis | 2017 | Australia, Belgium, Germany, USA, France, United Kingdom, Netherlands – multicentre collaboration | To identify urinary peptides predictive of future ACS, build the ACSP75 classifier, and assess whether a composite score improves prediction beyond Framingham risk estimates. |
| 10 | Hernesniemi JA, Mahdiani S, Tynkkynen JA, Lyytikäinen L-P, Mishra PP, Lehtimäki T, et al. Extensive phenotype data and machine learning in prediction of mortality in acute coronary syndrome – the MADDEC study | 2019 | Finland – Tays Heart Hospital / Tampere University Hospital; Tampere University; VTT Technical Research Centre of Finland | To determine whether extended phenotyping from EHR data combined with machine learning improves prediction of 6-month mortality after ACS compared with the GRACE score. |
| 11 | Neumann JT, Sörensen NA, Zeller T, Magaret C, Barnes G, Rhyne RF, et al. Application of a machine learning-driven, multibiomarker panel for prediction of incident cardiovascular events in patients with suspected myocardial infarction | 2020 | Germany – University Heart & Vascular Center Hamburg / DZHK; USA – Prevencio Inc. and Massachusetts General Hospital / Baim Institute | To externally validate a machine-learning multibiomarker panel for predicting 1-year MACE in patients with suspected MI and compare it with hs-cTnI. |
| 12 | Segar MW, Patel KV, Ayers C, Basit M, Tang WHW, Willett D, et al. Phenomapping of patients with heart failure with preserved ejection fraction using machine learning-based unsupervised cluster analysis | 2019 | USA – University of Texas Southwestern Medical Center and Cleveland Clinic | To identify phenotypically distinct HFpEF subgroups using unsupervised machine learning to improve risk stratification and understanding of heterogeneity. |
| 13 | Zhang K-P, Guo Q-C, Mu N, Liu C-H. Establishment and validation of nomogram model for predicting major adverse cardiac events in patients with acute ST-segment elevation myocardial infarction based on glycosylated hemoglobin A1c to apolipoprotein A1 ratio | 2024 | China – Huaibei Miners’ General Hospital; First People’s Hospital of Hefei | To assess the utility of the HbA1c/ApoA1 ratio for predicting in-hospital MACE in STEMI patients after PCI and to develop and validate a nomogram. |
| 14 | Li CKH, Xu Z, Ho J, Lakhani I, Bazoukis G, Liu T, et al. Association of NPAC score with survival after acute myocardial infarction | 2020 | Hong Kong – Chinese University of Hong Kong; Prince of Wales Hospital; City University of Hong Kong; external validation centre in Hong Kong | To develop and validate a simple four-component NPAC score for 90-day mortality after first AMI and examine whether deep learning improves classification. |
| 15 | Kusunose K, Abe T, Haga A, Fukuda D, Yamada H, Harada M, et al. A Deep Learning Approach for Assessment of Regional Wall Motion Abnormality From Echocardiographic Images | 2019 | Japan – Tokushima University Hospital; external validation at Hoetsu Hospital | To determine whether a deep convolutional neural network can automatically detect regional wall-motion abnormalities and localize infarct territory on standard 2D echocardiographic images. |
| 16 | Hoogeveen RM, Belo Pereira JP, Nurmohamed NS, et al. Improved cardiovascular risk prediction using targeted plasma proteomics in primary prevention | 2020 | Netherlands, United Kingdom, Italy, Germany | To develop and validate a proteomics-based cardiovascular risk prediction model in primary prevention. |
| 17 | Chan MY, Motakis E, Tan SH, Pickering JW, Troughton R, Pemberton C, et al. Prioritizing Candidates of Post-Myocardial Infarction Heart Failure Using Plasma Proteomics and Single-Cell Transcriptomics | 2020 | Singapore, New Zealand, Malaysia | To prioritize candidate biomarkers for post-MI heart failure by integrating plasma proteomics with single-cell transcriptomics and validating findings in an independent cohort. |
| 18 | Zhou P, Wan J, Ran F, Gao F, Yang D, Dai X, et al. Development and validation of a prognostic prediction model for antithrombotic-related chronic subdural hematoma in patients with recent acute myocardial infarction | 2020 | China – multicentre: Chengdu Medical College, Western Theater Command General Hospital, Anhui Medical University | To develop and validate a nomogram predicting adverse 6-month outcomes in patients with recent AMI complicated by antithrombotic-related chronic subdural hematoma. |
| 19 | Burrello J, Bolis S, Balbi C, Burrello A, Provasi E, Caporali E, et al. An extracellular vesicle epitope profile is associated with acute myocardial infarction | 2020 | Switzerland – Cardiocentro Ticino Institute; Italy – University of Torino and collaborating centres | To investigate whether extracellular-vesicle surface epitope profiles can serve as diagnostic biomarkers distinguishing AMI patients from healthy controls and chronic CAD. |
| 20 | Al-Zaiti SS, Besomi L, Bouzid Z, Faramand Z, Frisch S, Martin-Gill C, et al. Machine learning-based prediction of acute coronary syndrome using only the pre-hospital 12-lead electrocardiogram | 2020 | USA – University of Pittsburgh, UPMC hospitals; collaboration with Philips Healthcare | To develop and validate a machine-learning algorithm using prehospital 12-lead ECG alone for early detection of ACS, including NSTE-ACS, and compare it with expert interpretation and commercial ECG software. |
| 21 | Lin A, Kolossváry M, Yuvaraj J, Cadet S, McElhinney PA, Jiang C, et al. Myocardial Infarction Associates With a Distinct Pericoronary Adipose Tissue Radiomic Phenotype | 2020 | USA, Australia, Hungary | To determine whether CCTA-based pericoronary adipose tissue radiomics can distinguish acute MI from stable CAD and no CAD, and to build an ML model integrating clinical and radiomic features. |
| 22 | Wolfien M, Klatt D, Salybekov AA, Ii M, Komatsu-Horii M, Gaebel R, et al. The novel cystatin C, lactate, interleukin-6, and N-terminal pro-B-type natriuretic peptide (CLIP)-based mortality risk score in cardiogenic shock after acute myocardial infarction | 2020 | Germany and Japan – multicentre collaboration | To identify and validate a biomarker-based signature associated with mortality and cardiac recovery after MI-related cardiogenic shock and regenerative therapy contexts. |
| 23 | Yan P, Liu T, Zhang K, Cao J, Dang H, Song Y, et al. Development and Validation of a Novel Nomogram for Preoperative Prediction of In-Hospital Mortality After Coronary Artery Bypass Grafting Surgery in Heart Failure With Reduced Ejection Fraction | 2021 | China – Beijing Anzhen Hospital, Capital Medical University | To develop and validate a nomogram predicting in-hospital mortality after CABG in patients with HFrEF and compare its performance with EuroSCORE II. |
| 24 | Lin W-C, Hsiung M-C, Yin W-H, Tsao T-P, Lai W-T, Huang K-C. Electrocardiography Score for Left Ventricular Systolic Dysfunction in Non-ST Segment Elevation Acute Coronary Syndrome | 2022 | Taiwan – Cheng Hsin General Hospital; National Taiwan University | To develop and validate an ECG-based score predicting left ventricular systolic dysfunction in NSTE-ACS and assess its prognostic value for 24-month mortality. |
| 25 | Chen H, Wang Z, Qin M, Zhang B, Lin L, Ma Q, et al. Comprehensive Metabolomics Identified the Prominent Role of Glycerophospholipid Metabolism in Coronary Artery Disease Progression | 2021 | China – Guangdong Provincial People’s Hospital; Xiangya Hospital; Sun Yat-sen University | To identify metabolomic and lipidomic markers of CAD progression and develop diagnostic models differentiating CAD subtypes. |
| 26 | Rojas-Mendizabal V, Castillo-Olea C, Gómez-Siono A, Zuñiga C. Assessment of Thoracic Pain Using Machine Learning: A Case Study from Baja California, Mexico | 2021 | Mexico – CETYS Universidad, Autonomous University of Baja California, General Hospital of Tijuana | To identify factors most strongly associated with cardiac chest pain / AMI and propose secondary triage parameters in the emergency setting. |
| 27 | Stewart J, Lu J, Goudie A, Bennamoun M, Sprivulis P, Sanfilippo F, Dwivedi G. Applications of machine learning to undifferentiated chest pain in the emergency department: A systematic review | 2021 | Australia – University of Western Australia and collaborating institutions | To systematically review machine-learning applications in adults with undifferentiated chest pain in the emergency department and assess whether ML outperforms clinicians or existing tools. |
| 28 | Liu W-C, Lin C, Lin C-S, Tsai M-C, Chen S-J, Tsai S-H, et al. An Artificial Intelligence-Based Alarm Strategy Facilitates Management of Acute Myocardial Infarction | 2021 | Taiwan – Tri-Service General Hospital, National Defense Medical Center | To develop and implement an AI-based alarm strategy for early AMI recognition in the emergency department and assess its effect on treatment times. |
| 29 | Jung S, Ahn E, Koh SB, Lee S-H, Hwang G-S. Purine metabolite-based machine learning models for risk prediction, prognosis, and diagnosis of coronary artery disease | 2021 | South Korea – Korea Basic Science Institute, Yonsei University, Ewha Womans University | To evaluate whether serum purine metabolites can support machine-learning models for long-term risk prediction, prognosis, and diagnosis in coronary artery disease. |
| 30 | Marashly Q, Taleb I, Kyriakopoulos CP, et al. Predicting mortality in cardiogenic shock secondary to ACS requiring short-term mechanical circulatory support: The ACS-MCS score | 2021 | USA – University of Utah Health | To identify predictors and develop a risk score for 30-day mortality in ACS-related cardiogenic shock requiring short-term mechanical circulatory support. |
| 31 | Mehran R, Owen R, Chiarito M, Baber U, Sartori S, Cao D, et al. A contemporary simple risk score for prediction of contrast-associated acute kidney injury after percutaneous coronary intervention | 2021 | USA – Mount Sinai Hospital; collaboration with LSHTM | To derive and validate a contemporary simple risk score (Mehran 2) for CA-AKI after PCI and assess its association with 1-year mortality. |
| 32 | Serrano AB, Gomez-Rojo M, Ureta E, Nuñez M, Fernández Félix B, Velasco E, et al. Preoperative clinical model to predict myocardial injury after non-cardiac surgery | 2021 | Spain – Ramón y Cajal University Hospital, Madrid | To identify preoperative factors associated with myocardial injury after non-cardiac surgery and develop a nomogram for risk prediction. |
| 33 | Panchavati S, Lam C, Zelin NS, Pellegrini E, Barnes G, Hoffman J, et al. Retrospective validation of a machine learning clinical decision support tool for myocardial infarction risk stratification | 2021 | USA – large academic centre using EHR data | To retrospectively validate a machine-learning algorithm for MI prediction based on the first 3 hours of emergency department EHR data and compare it with TIMI and GRACE. |
| 34 | Wei W, Zhang L, Zhang Y, Tang R, Zhao M, Huang Z, et al. Predictive value of creatine kinase MB for contrast-induced acute kidney injury among myocardial infarction patients | 2021 | China – REICIN multicentre registry coordinated by Guangdong Provincial People’s Hospital | To assess whether peak preprocedural CK-MB predicts contrast-induced AKI in MI patients and whether it improves ACEF and Mehran models. |
| 35 | Yeung W, Sia C-H, Pollard T, Leow AST, Tan BYQ, Kaur R, et al. Predicting mortality, thrombus recurrence and persistence in patients with post-acute myocardial infarction left ventricular thrombus | 2021 | Singapore – National University Hospital / National University Heart Centre / MIT-HST collaboration | To develop a model predicting mortality and thrombus persistence / recurrence in patients with post-AMI left ventricular thrombus using data available at diagnosis. |
| 36 | Helgestad OKL, Povlsen AL, Josiassen J, Møller S, Hassager C, Jensen LO, et al. Data-driven point-of-care risk model in patients with acute myocardial infarction and cardiogenic shock | 2021 | Denmark – Odense University Hospital and Copenhagen University Hospital (Rigshospitalet) | To develop a point-of-care risk model for 30-day mortality in AMI-related cardiogenic shock before PCI and compare it with IABP-SHOCK II and CardShock. |
| 37 | Yifan C, Shi J, Pu J. Development and Validation of a Random Forest Diagnostic Model of Acute Myocardial Infarction Based on Ferroptosis-Related Genes in Circulating Endothelial Cells | 2021 | China – Renji Hospital, Shanghai Jiao Tong University | To create a diagnostic random-forest model based on ferroptosis-related genes in circulating endothelial cells for early AMI detection. |
| 38 | Chen S, Pan X, Mo J, Wang B. Establishment and validation of a prediction nomogram for heart failure risk in patients with acute myocardial infarction during hospitalization | 2023 | China – Wenzhou Medical University Affiliated Dongyang Hospital | To develop and validate a nomogram predicting the risk of heart failure during hospitalization in patients with AMI. |
| 39 | Vázquez B, Fuentes-Pineda G, García F, Borrayo G, Prohías J. Risk markers by sex for in-hospital mortality in patients with acute coronary syndrome: A machine learning approach | 2021 | Mexico and Cuba – UNAM, IMSS, Hermanos Ameijeiras | To identify sex-specific risk markers for in-hospital mortality in ACS, including STEMI and NSTEMI subgroups, using EHR-based machine learning. |
| 40 | Li X, Yan F, Liu X, Li M, Li J, Chen Y, Li C. Acute coronary syndrome screening in patients presenting with arteriosclerosis in health check-ups: a case-control study | 2022 | China – Qilu Hospital of Shandong University | To develop a simple automated machine-learning model for early ACS screening in patients with atherosclerosis identified during health check-ups. |
| 41 | Chen Z, Shi J, Pommier T, Cottin Y, Salomon M, Decourselle T, et al. Prediction of Myocardial Infarction From Patient Features With Machine Learning | 2022 | France – University Hospital of Dijon, FEMTO-ST Institute, CASIS Company | To develop and validate machine-learning models for automatic assessment of MI severity from clinical, paraclinical, and physiological features to support emergency department decision-making. |
| 42 | Rong F, Xiang H, Qian L, Xue Y, Ji K, Yin R. Machine Learning for Prediction of Outcomes in Cardiogenic Shock | 2022 | China – Wenzhou Medical University; data from MIMIC-III, USA | To develop a machine-learning prognostic model for 30-day mortality in older patients with cardiogenic shock using CoxBoost. |
| 43 | Cai Y-L, Hao B-C, Chen J-Q, Li Y-R, Liu H-B. Correlation Between Plasma Proteomics and Adverse Outcomes Among Older Men With Chronic Coronary Syndrome | 2022 | China – PLA General Hospital, Beijing | To identify proteomic biomarkers predictive of MACE in older men with chronic coronary syndrome using proteomics and machine learning. |
| 44 | Cai D, Xiao T, Zou A, Mao L, Chi B, Wang Y, et al. Predicting acute kidney injury risk in acute myocardial infarction patients: An artificial intelligence model using MIMIC databases | 2022 | China – Affiliated Changzhou No. 2 People’s Hospital of Nanjing Medical University; data from MIMIC-III and IV | To develop and validate a machine-learning model for predicting AKI risk in AMI patients using critical-care databases. |
| 45 | Chen H, Jiang R, Huang W, Chen K, Zeng R, Wu H, et al. Identification of energy metabolism-related biomarkers for risk prediction of heart failure patients using random forest algorithm | 2022 | China and Hong Kong – Guangdong Provincial People’s Hospital, Southern Medical University, Queen Mary Hospital | To identify energy metabolism-related biomarkers predictive of heart-failure risk in post-MI patients using random forest and transcriptomic analysis. |
| 46 | Wang Y, Li C, Yuan M, Ren B, Liu C, Zheng J, et al. Development of a complete blood count with differential-based prediction model for in-hospital mortality among patients with acute myocardial infarction in the coronary care unit | 2022 | China – Xi’an Jiaotong University and collaborating institutions; data from MIMIC-IV | To develop a prediction model for in-hospital mortality in AMI patients in the coronary care unit based on CBC with differential using neural networks. |
| 47 | Yang G, Zhou S, He H, Shen Z, Liu Y, Hu J, Wang J. Exploring the gene–protein–metabolite network of coronary heart disease with phlegm and blood stasis syndrome by integrated multi-omics strategy | 2022 | China – Guang’anmen Hospital, China Academy of Chinese Medical Sciences | To identify the biological basis of phlegm and blood stasis syndrome in CHD through integrated multi-omics analysis and validate diagnostic biomarkers. |
| 48 | Liu M, Wang M, Peng T, Ma W, Wang Q, Niu X, et al. Gut-microbiome-based predictive model for ST-elevation myocardial infarction in young male patients | 2022 | China – Tangdu Hospital and Xijing Hospital, Fourth Military Medical University | To develop an early predictive model of STEMI risk in young men based on gut microbiome data and clinical parameters. |
| 49 | Liu J, Huang L, Shi X, Gu C, Xu H, Liu S. Clinical Parameters and Metabolomic Biomarkers That Predict Inhospital Outcomes in Patients With ST-Segment Elevated Myocardial Infarctions | 2022 | China – Third Central Hospital of Tianjin | To develop an early prognostic model for in-hospital outcomes in STEMI after PCI by integrating metabolomic biomarkers with clinical parameters. |
| 50 | Yang J, Ouyang X, Yang M, Xie G, Cao Q. Identification of key programmed cell death-related genes and immune infiltration in extracorporeal membrane oxygenation treatment for acute myocardial infarction | 2022 | China – Jiangxi Provincial People’s Hospital / Nanchang Medical College | To identify programmed cell death-related genes and immune-infiltration patterns in AMI patients with cardiogenic shock treated with ECMO in search of prognostic biomarkers. |
| 51 | Xu C, Li W, Li T, Yuan J, Pang X, Liu T, et al. Iron metabolism-related genes reveal predictive value of acute coronary syndrome | 2022 | China – Shenzhen People’s Hospital; Xiangya Hospital | To identify iron metabolism-related genes and develop a molecular diagnostic signature for ACS. |
| 52 | Nishi M, Uchino E, Okuno Y, Matoba S. Robust prognostic prediction model developed with integrated biological markers for acute myocardial infarction | 2022 | Japan – Kyoto Prefectural University of Medicine and Kyoto University | To develop a prognostic machine-learning model for STEMI using integrated biological and clinical markers and support clinical application through a dedicated tool. |
| 53 | Zheng J, Zhang J, Zheng M, Zhang Y, Zhao X, Wang K. Artificial Intelligence-Enabled Electrocardiography Predicts Left Ventricular Dysfunction and Future Cardiovascular Outcomes | 2022 | China – Beijing Chaoyang Hospital, Capital Medical University | To identify risk factors and support prediction of left ventricular dysfunction and subsequent cardiovascular outcomes using AI-enabled ECG. |
| 54 | Burrello J, Gallone G, Burrello A, Jahier Pagliari D, Ploumen EH, Iannaccone M, et al. Prediction of All-Cause Mortality Following Percutaneous Coronary Intervention in Bifurcation Lesions Using Machine Learning Algorithms | 2022 | Italy with international collaboration – RAIN registry, DUTCH-PEERS, BIO-RESORT | To develop a machine-learning model for 2-year all-cause mortality after PCI in bifurcation lesions using clinical, anatomical, and procedural variables. |
| 55 | Shetty MK, Kunal S, Girish MP, Qamar A, Arora S, Bhatt DL, Gupta A, et al. Machine learning based model for risk prediction after ST-Elevation myocardial infarction: Insights from the NORIN-STEMI registry | 2022 | India – MAMC, GB Pant Hospital, AIIMS, IIIT Delhi; collaboration with USA | To develop and validate a machine-learning model for 30-day mortality after STEMI using registry data and explainability methods such as SHAP and LIME. |
| 56 | Emakhu J, Monplaisir L, Aguwa C, Arslanturk S, Masoud S, Nassereddine H, Hamam MS, Miller JB. Acute Coronary Syndrome Prediction in Emergency Care: A Machine Learning Approach | 2022 | USA – Wayne State University and Henry Ford Hospital | To develop a machine-learning diagnostic framework for classifying patients with suspected ACS in the emergency department and distinguishing NSTEMI / unstable angina from non-cardiac chest pain. |
| 57 | Liu W, Zhang L, Shi X, Shen G, Feng J. Cross-comparative metabolomics reveal sex-age specific metabolic fingerprints and metabolic interactions in acute myocardial infarction | 2022 | China – Zhongshan Hospital Affiliated to Xiamen University; First Affiliated Hospital of Xiamen University | To characterize sex- and age-specific metabolic disturbances in AMI and identify single and composite biomarkers for differentiating healthy controls, AMI, and AMI with cardiovascular disease. |
| 58 | Rudnicka AR, Welikala R, Barman S, Foster PJ, Luben R, Hayat S, et al. Artificial intelligence-enabled retinal vasculometry for prediction of circulatory mortality, myocardial infarction and stroke | 2022 | United Kingdom – St George’s University of London, Kingston University, UCL, University of Cambridge | To evaluate whether AI-enabled retinal vasculometry improves prediction of circulatory mortality, stroke, and MI beyond standard risk models. |
| 59 | Zhu X, Li K, Chen M. Nomogram for Risk Prediction of Mortality for Patients with Critical Cardiovascular Disease Treated by Continuous Renal Replacement Therapy in Coronary Care Unit | 2022 | China – Beijing Chaoyang Hospital, Capital Medical University | To develop and validate a simple nomogram predicting in-hospital mortality in critically ill cardiovascular patients treated with CRRT in the coronary care unit. |
| 60 | Simon S, Mandair D, Albakri A, Fohner A, Simon N, Lange L, et al. The Impact of Time Horizon on Classification Accuracy: Application of Machine Learning to Prediction of Incident Coronary Heart Disease | 2022 | USA – University of Colorado, University of Washington, Beth Israel Deaconess | To determine the optimal time horizon for machine-learning classification of incident MI / CHD risk and compare performance with Framingham-based prediction. |
| 61 | Jentzer JC, Soussi S, Lawler PR, Kennedy JN, Kashani KB. Validation of cardiogenic shock phenotypes in a mixed cardiac intensive care unit population | 2022 | USA – Mayo Clinic; international collaboration | To validate previously described cardiogenic shock phenotypes in a mixed CICU population using unsupervised clustering and assess their clinical relevance. |
| 62 | Liu S, Yang S, Xing A, Zheng L, Shen L, Tu B, Yao Y. Machine learning-based long-term outcome prediction in patients undergoing percutaneous coronary intervention | 2021 | China – Fuwai Hospital, National Center for Cardiovascular Diseases, Beijing | To develop and validate machine-learning models for predicting 5-year all-cause mortality after PCI and identify the most important risk factors. |
| 63 | Juan-Salvadores P, Veiga C, Jiménez Díaz VA, Guitián González A, Iglesia Carreño C, Martínez Reglero C, et al. Using Machine Learning Techniques to Predict MACE in Very Young Acute Coronary Syndrome Patients | 2022 | Spain – Hospital Álvaro Cunqueiro, Vigo; IIS Galicia Sur; University of Santiago de Compostela | To predict MACE in very young ACS patients after coronary angiography and compare machine-learning algorithms with logistic regression. |
| 64 | Vezzoli M, Inciardi RM, Oriecuia C, Paris S, et al. Machine learning for prediction of in-hospital mortality in coronavirus disease 2019 patients | 2021 | Italy – 13 cardiology departments, multicentre study | To develop an in-hospital mortality risk score in COVID-19 patients using a limited number of admission variables. |
| 65 | Jin BT, Palleti R, Shi S, Ng AY, Quinn JV, Rajpurkar P, Kim D. Transfer learning enables prediction of myocardial injury from continuous single-lead electrocardiography | 2022 | USA – Stanford University and Harvard University | To develop a predictive model of myocardial injury from continuous single-lead ECG using transfer learning from large labelled 12-lead ECG datasets. |
| 66 | Doudesis D, Lee KK, Yang J, Wereski R, Shah ASV, Tsanas A, Anand A, Pickering JW, Than MP, Mills NL. Validation of the myocardial-ischaemic-injury-index machine learning algorithm to guide the diagnosis of myocardial infarction in a heterogeneous population | 2022 | Multicentre / international (exact centre details not fully specified in the source list) | To validate a machine-learning myocardial ischaemic injury index for guiding MI diagnosis in a heterogeneous patient population. |
| 67 | Backhaus SJ, Aldehayat H, Kowallick JT, Evertz R, Lange T, Kutty S, et al. Artificial intelligence fully automated myocardial strain quantification for risk stratification following acute myocardial infarction | 2022 | Germany – multicentre; collaboration with USA | To assess reproducibility of fully automated AI-based LV strain quantification from cine CMR and evaluate its prognostic value for 12-month MACE after AMI. |
| 68 | Nishi M, Uchino E, Okuno Y, Matoba S. Robust prognostic prediction model developed with integrated biological markers for acute myocardial infarction | 2022 | Japan – Kyoto Prefectural University of Medicine; AMI-Kyoto multicentre registry | To develop a machine-learning model and online tool for in-hospital mortality prediction in STEMI and compare performance with GRACE and TIMI. |
| 69 | Liu J, Huang L, Shi X, Gu C, Xu H, Liu S. Clinical Parameters and Metabolomic Biomarkers That Predict Inhospital Outcomes in Patients With ST-Segment Elevated Myocardial Infarctions | 2022 | China – Third Central Hospital of Tianjin | To identify metabolic fingerprints associated with different in-hospital outcomes in STEMI after PCI and develop a prognostic model integrating metabolomics and clinical variables. |
| 70 | Wang Y, Li C, Yuan M, Ren B, Liu C, Zheng J, Lin Z, Ren F, Gao D. Development of a complete blood count with differential-based prediction model for in-hospital mortality among patients with acute myocardial infarction in the coronary care unit | 2022 | China and USA – Xi’an Jiaotong University and collaborators; MIMIC-IV | To develop a prediction model for in-hospital mortality in AMI patients in the CCU using CBC with differential and compare it with multiple ML models and CBC-derived indicators. |
| 71 | Corral Acero J, Schuster A, Zacur E, Lange T, Stiermaier T, Backhaus SJ, et al. Understanding and Improving Risk Assessment After Myocardial Infarction Using Automated Left Ventricular Shape Analysis | 2022 | United Kingdom and Germany – University of Oxford and German clinical centres | To develop an automated AI pipeline for 3D analysis of LV shape and function from CMR after AMI and identify novel prognostic markers for 12-month MACE. |
| 72 | Shetty MK, Kunal S, Girish MP, Qamar A, Arora S, Hendrickson M, et al. Machine learning based model for risk prediction after ST-Elevation myocardial infarction: Insights from the NORIN-STEMI registry | 2022 | India and USA | To develop and validate an interpretable machine-learning model for 30-day mortality after STEMI in a low-resource setting using the NORIN-STEMI registry. |
| 73 | Liu W, Zhang L, Shi X, Shen G, Feng J. Cross-comparative metabolomics reveal sex-age specific metabolic fingerprints and metabolic interactions in acute myocardial infarction | 2022 | China – Xiamen University and affiliated hospitals | To identify metabolomic disturbances and sex- / age-specific fingerprints in AMI and derive multi-biomarker sets for differentiating stages and outcomes of AMI. |
| 74 | Li X, Shang C, Xu C, Wang Y, Xu J, Zhou Q. Development and comparison of machine learning-based models for predicting heart failure after acute myocardial infarction | 2023 | China – First Hospital of Jilin University | To compare seven machine-learning algorithms and develop an early warning model for heart failure after AMI based on routine laboratory data. |
| 75 | Zhou Q, Boeckel J-N, Yao J, Zhao J, et al. Diagnosis of acute myocardial infarction using a combination of circulating circular RNA cZNF292 and clinical information based on machine learning | 2023 | China and Germany – Tongji Hospital, Shanghai Tenth People’s Hospital, Goethe University, Leipzig collaborators | To determine whether combining circulating circRNA cZNF292 with clinical information and machine learning improves AMI diagnosis. |
| 76 | Fang C, Li J, Wang W, Wang Y, Chen Z, Zhang J. Establishment and validation of a clinical nomogram model based on serum YKL-40 to predict major adverse cardiovascular events during hospitalization in patients with acute ST-segment elevation myocardial infarction | 2023 | China – Second People’s Hospital of Hefei / Anhui Medical University | To evaluate the prognostic value of YKL-40 and develop a nomogram for predicting in-hospital MACE in STEMI patients after PCI. |
| 77 | Li M, Zeng D, Zhou Y, Chen J, Cao S, Song H, et al. A novel risk stratification model for STEMI after primary PCI: global longitudinal strain and deep neural network assisted myocardial contrast echocardiography quantitative analysis | 2023 | China – Renmin Hospital of Wuhan University; collaboration with USA | To develop and validate a risk-stratification model for MACE after primary PCI in STEMI using DNN-assisted myocardial contrast echocardiography and global longitudinal strain. |
| 78 | Chen S, Pan X, Mo J, Wang B. Establishment and validation of a prediction nomogram for heart failure risk in patients with acute myocardial infarction during hospitalization | 2023 | China – Dongyang People’s Hospital / Wenzhou Medical University | To develop and validate a nomogram for prediction of heart failure during hospitalization in AMI. |
| 79 | Zhang S, Zhu Z, Luo M, Chen L, He C, You Z, et al. The optimal definition and prediction nomogram for left ventricular remodelling after acute myocardial infarction | 2023 | China – Fujian Provincial Hospital / Fujian Medical University | To identify the optimal definition of left ventricular remodelling after AMI and develop and validate a nomogram for its prediction. |
| 80 | Tindale A, Cretu I, Meng H, Panoulas V. Complete revascularization is associated with higher mortality in patients with ST-elevation myocardial infarction, multi-vessel disease and shock defined by hyperlactataemia | 2023 | United Kingdom – Harefield Hospital, Imperial College London | To evaluate the impact of complete versus culprit-only revascularization on mortality in STEMI patients with multivessel disease and shock defined by hyperlactataemia. |
| 81 | Liu W, Zhang L, Bao L, Shen G, Feng J. Accurate Classification and Prediction of Acute Myocardial Infarction through an ARMD Procedure | 2023 | China – Zhongshan Hospital and First Affiliated Hospital of Xiamen University | To develop an ARMD procedure for AMI classification and outcome prediction and identify characteristic metabolites for different clinical trajectories. |
| 82 | Zhao J, Zhao P, Li C, Hou Y. Optimized Machine Learning Models to Predict In-Hospital Mortality for Patients with ST-Segment Elevation Myocardial Infarction | 2021 | China – Tianjin Medical University / Tianjin Chest Hospital / Tianjin University | To optimize machine-learning models for predicting in-hospital mortality in STEMI and compare full versus simplified pre-reperfusion feature sets. |
| 83 | Shakhgeldyan KI, Kuksin NS, Domzhalov IG, Geltser BI. Performance of the Models Predicting In-Hospital Mortality in Patients with ST-Segment Elevation Myocardial Infarction with Predictors in Categorical and Continuous Forms | 2024 | Russia – Vladivostok State University; Far Eastern Federal University; Regional Vascular Center, Vladivostok | To evaluate models for predicting in-hospital mortality after PCI in STEMI using predictors in continuous versus categorized form and compare cut-off selection approaches. |
| 84 | Song H, Qin W-B, Yang F-F, Tang W-Z, He G-X. Risk analysis and risk prediction of in-hospital heart failure in patients with acute myocardial infarction after emergency intervention surgery | 2024 | China – First Affiliated Hospital of Guangxi University of Chinese Medicine | To identify risk factors for heart failure during hospitalization after emergency PCI in AMI and develop a personalized predictive model / nomogram. |
| 85 | Hou S, Zhang L, Ji H, Zhao T, Hu M, Jiang Y, et al. Development and evaluation of the model for acute kidney injury in patients with cardiac arrest after successful resuscitation | 2024 | China – First Affiliated Hospital of USTC; Wannan Medical College | To develop and evaluate a clinical model predicting AKI after successful resuscitation from cardiac arrest and create a visual nomogram. |
| 86 | Schweiger V, Hiller P, Utters R, Fenice A, Cammann VL, Di Vece D, et al. A novel score to predict in-hospital mortality for patients with acute coronary syndrome and out-of-hospital cardiac arrest: the FACTOR study | 2024 | Switzerland – University Hospital Zurich / University of Zurich and EMS Zurich | To develop and validate a simple admission score for in-hospital mortality prediction in patients with OHCA caused by ACS undergoing coronary angiography. |
| 87 | Li P, Yao W, Wu J, Gao Y, Zhang X, Hu W. Development and Validation of a Nomogram Model for Predicting in-Hospital Mortality in non-Diabetic Patients with non-ST-Segment Elevation Acute Myocardial Infarction | 2024 | China – Xiaogan Hospital affiliated with Wuhan University of Science and Technology; Jilin University | To develop and validate a nomogram for predicting in-hospital mortality in non-diabetic patients with NSTEMI for rapid risk stratification. |
| 88 | Zhang P, Wu L, Zou T-T, Zou Z, Tu J, Gong R, Kuang J. Machine Learning for Early Prediction of Major Adverse Cardiovascular Events After First Percutaneous Coronary Intervention in Patients With Acute Myocardial Infarction | 2024 | China – Second Affiliated Hospital of Nanchang University | To develop and compare machine-learning models for early prediction of MACE after first PCI in newly diagnosed AMI and identify the most important predictors. |
| 89 | Kirdeev A, Burkin K, Vorobev A, Zbirovskaya E, Lifshits G, Nikolaev K, et al. Machine learning models for predicting risks of MACEs for myocardial infarction patients with different VEGFR2 genotypes | 2024 | Russia – Surgut District Center for Diagnostics and Cardiovascular Surgery; Surgut State University; HSE University | To build machine-learning models for long-term MACE prediction after MI using clinical, imaging, laboratory, and VEGFR2 genotype data and assess the incremental value of genetics. |
| 90 | Zheng W, Guo Q, Guo R, et al. Predicting left ventricular remodeling post-MI through coronary physiological measurements based on computational fluid dynamics | 2024 | China – Beijing Anzhen Hospital, Capital Medical University | To develop and assess machine-learning models for predicting left ventricular remodelling after STEMI and test whether coronary physiological measures plus echocardiography can substitute for CMR. |
| 91 | Jing J, Wan L, Wang M. A Practical Nomogram for Predicting the Bleeding Risk in Patients with a History of Myocardial Infarction Treating with Aspirin | 2024 | China – Affiliated Dongyang Hospital, Wenzhou Medical University | To create a practical nomogram predicting bleeding risk in patients with prior MI treated with aspirin. |
| 92 | Roudini B, Khajehpiri B, Moghaddam HA, Forouzanfar M. Machine learning predicts long-term mortality after acute myocardial infarction using systolic time intervals and routinely collected clinical data | 2024 | Taiwan data source with analysis in Canada / Iran | To evaluate machine-learning models for 14-year all-cause mortality after AMI and assess the added predictive role of systolic time interval biomarkers. |
| 93 | Gupta A, Gupta A, Shetty M, Goyal D, Girish MP, Gupta M. CardioGraph: GCN-Inspired Short-Term Mortality Predictor in First-Time STEMI Patients | 2024 | India – G.B. Pant Hospital, New Delhi; IIIT-Delhi | To develop a graph-convolution-inspired model (CardioGraph) for short-term mortality prediction after first STEMI in the Indian population. |
| 94 | Xia Q, Deng C, Yang S, Gu N, Shen Y, Shi B, Zhao R. Machine Learning Constructed Based on Patient Plaque and Clinical Features for Predicting Stent Malapposition | 2024 | China – Affiliated Hospital of Zunyi Medical University | To develop machine-learning models for predicting stent malapposition after PCI in AMI by integrating OCT plaque features and clinical variables. |
| 95 | Zheng H, Sherazi SWA, Lee JY. A cost-sensitive deep neural network-based prediction model for the mortality in acute myocardial infarction patients with hypertension on imbalanced data | 2024 | South Korea – KAMIR-NIH registry; collaboration with China | To predict 24-month mortality in patients with AMI and hypertension using cost-sensitive deep neural networks and threshold moving on imbalanced data. |
| 96 | Jeong JH, Lee K-S, Park S-M, Kim SR, Kim M-N, Chae SC, et al. Prediction of longitudinal clinical outcomes after acute myocardial infarction using a dynamic machine learning algorithm | 2024 | South Korea – KAMIR-NIH registry, 20 tertiary centres | To develop machine-learning models for dynamic time-dependent prediction of clinical outcomes after AMI, including 1-year mortality and MACE at 1 and 3 years. |
| 97 | Järvensivu-Koivunen M, Kallonen A, van Gils M, Lyytikäinen L-P, Tynkkynen J, Hernesniemi J. Predicting long-term risk of sudden cardiac death with automatic computer-interpretations of electrocardiogram | 2024 | Finland – Tampere University / Tays Heart Hospital (MADDEC) | To assess whether automatic 12-lead ECG interpretations can predict 5-year risk of sudden cardiac death after ACS as a proof of concept. |
| 98 | Hathaway QA, Jamthikar AD, Rajiv N, Chaitman BR, Carson JL, Yanamala N, Sengupta PP. Cardiac ultrasomics for acute myocardial infarction risk stratification and prediction of all-cause mortality: a feasibility study | 2024 | USA – Rutgers RWJMS, University of Pennsylvania, Saint Louis University; external DTU-STEMI pilot cohort | To stratify AMI risk and predict 1-year all-cause mortality using echocardiographic ultrasomics / radiomics and assess incremental value beyond GRACE 2.0. |
| 99 | Kirdeev A, Burkin K, Vorobev A, Zbirovskaya E, Lifshits G, Nikolaev K, et al. Machine learning models for predicting risks of MACEs for myocardial infarction patients with different VEGFR2 genotypes | 2024 | Russia – Surgut District Center and collaborating centres | To develop machine-learning models for long-term MACE prediction after MI and evaluate whether VEGFR2 genotype improves prognostic performance. |
| 100 | Li D, Liu T, Li Z. Sema4D combined with electrocardiographic parameters for predicting STEMI prognosis: Development and validation of a nomogram model | 2025 | China – Zhongshan People’s Hospital, Guangdong Province; Zhejiang Hospital of Integrated Traditional Chinese and Western Medicine | To evaluate the influence of clinical and electrocardiographic parameters on STEMI prognosis and develop a personalized nomogram for adverse outcome prediction. |
| 101 | Zhou Z, Chen Q, Zhang Z, Wang T, Zhao Y, Chen W, et al. Early prediction of microvascular obstruction prior to percutaneous coronary intervention | 2025 | China – Affiliated Hospital of Xuzhou Medical University and collaborating centres | To develop a model for early prediction of microvascular obstruction before PCI in AMI using preprocedural clinical data and validate it prospectively. |
| 102 | Shahri B, Tajik A, Moohebati M, Mahdavizadeh V. Incorporating the STOP-BANG questionnaire improves prediction of cardiovascular events during hospitalization after myocardial infarction | 2025 | Iran – Ghaem Hospital, Mashhad University of Medical Sciences | To assess whether including the STOP-BANG questionnaire improves GRACE-based prediction of in-hospital cardiovascular events after MI and compare conventional and ML approaches. |
| 103 | Wei P, Hou J, Yu R, Na K, He J, Zhao X, et al. Development and validation of an LDH-focused nomogram for the early prediction of heart failure in patients with acute STEMI after PCI | 2025 | China – General Hospital of the Northern Theater Command; China Medical University | To develop and validate an LDH-based nomogram for early prediction of heart failure within 1 year after PCI in STEMI and assess the incremental value of LDH. |
| 104 | Yang Y, Tang J, Ma L, Wu F, Guan X. A systematic comparison of short-term and long-term mortality prediction in acute myocardial infarction using machine learning models | 2025 | China – Yueyang Hospital and Changhai Hospital, Shanghai | To develop machine-learning models for mortality prediction after AMI at 1, 5, and 10 years and systematically compare short- and long-term prognostic performance. |
| 105 | Deng L, Zhao X, Su X, Zhou M, Huang D, Zeng X. Machine learning to predict no reflow and in-hospital mortality in patients with STEMI that underwent primary PCI | 2022 | China – four-centre National Chest Pain Center Alliance, Guangxi | To develop and compare machine-learning models for predicting no-reflow and in-hospital mortality in STEMI patients undergoing primary PCI using clinical, laboratory, echocardiographic, and angiographic data. |
| 106 | Wang Y, Lu Y, Yang Y, Han S, Chi Q, Zhou Y, Duan Y. A predictive nomogram for intramyocardial hemorrhage in patients with STEMI undergoing primary PCI based on coronary angiography-derived IMR | 2025 | China – Affiliated Hospital of Xuzhou Medical University; collaborating institutions | To develop and validate a nomogram combining caIMR with clinical features to predict intramyocardial hemorrhage after primary PCI in STEMI and assess prognostic implications. |
| 107 | Guo C, Gao B, Han X, Zhang T, Tao T, Xia J, Liu H. Interpretable artificial intelligence model for predicting heart failure severity after acute myocardial infarction | 2025 | China – Xuanwu Hospital, Capital Medical University; collaborating centres | To develop an interpretable AI model predicting heart-failure severity after AMI using multidimensional clinical data and Killip class as the endpoint. |
| 108 | Yan M, Miao Y, Sheng S, Gan X, He B, Shen L. Ensemble Learning-Based Mortality Prediction After Acute Myocardial Infarction | 2025 | China – Shanghai Jiao Tong University / Shanghai Chest Hospital; data from seven hospitals in Shanghai | To develop a model for 2-year mortality prediction after AMI in a small and highly imbalanced cohort using ensemble learning and derive a simplified clinical model. |
| 109 | Mohd Faizal AS, Hon WY, Thevarajah TM, Khor SM, Chang S-W. A biomarker discovery of acute myocardial infarction using feature selection and machine learning | 2023 | Malaysia – Universiti Malaya / University of Malaya Medical Centre | To develop an integrated feature-selection and machine-learning framework for biomarker discovery and prognostic modelling in AMI with in-hospital mortality as the endpoint. |
| 110 | Prasad SB, Scanlon L, Krishnan A, Chan NI, Mallouhi M, Vollbon W, et al. Machine learning integration of echocardiographic and clinical data to improve prediction of survival following myocardial infarction | 2025 | Australia – Royal Brisbane and Women’s Hospital; University of Queensland; collaborators | To develop machine-learning models integrating echocardiographic and clinical data to predict all-cause mortality after MI and compare them with conventional logistic regression. |
| 111 | Du Z, Lu Y, Ma Y, Yang Y, Luo W, Liu S, et al. The prognostic and therapeutic significance of polyunsaturated fatty acid-derived oxylipins in STEMI | 2025 | China – Beijing Anzhen Hospital and Peking University Third Hospital | To evaluate the significance of PUFA-derived oxylipins in STEMI and develop an oxylipin-based risk model for recurrent MACE after STEMI. |
| 112 | Li W, Yan D, Hu W, Su X, Zhang Z. Enhancing one-year mortality prediction in STEMI patients post-PCI: an interpretable machine learning model with risk stratification | 2025 | China – The First Hospital of Lanzhou University; Qinghai Provincial People’s Hospital | To develop an interpretable machine-learning model for 1-year mortality after PCI in STEMI while reducing the number of predictors and improving clinical usability through risk stratification. |
| 113 | Li M, Song B, Zeng X, Wang X, Ma A, Meng Z, et al. Nomogram for predicting the 28-day mortality risk of patients with subendocardial infarction | 2025 | China; source data from MIMIC-III, USA | To develop and evaluate a nomogram for 28-day mortality prediction in patients with subendocardial myocardial infarction. |
| 114 | Lin Q, Zhao W, Zhang H, Chen W, Lian S, Ruan Q, et al. Predicting the risk of heart failure after acute myocardial infarction using an interpretable machine learning model | 2025 | China – First Affiliated Hospital of Fujian Medical University; Fuzhou University | To develop and validate an interpretable machine-learning model for early prediction of heart-failure risk after AMI. |
| 115 | Genc O, Yildirim A, Erdogan A, Ibisoglu E, Guler Y, Capar G, et al. Modification, validation and comparison of Naples prognostic score to determine in-hospital mortality in STEMI | 2025 | Turkey – multicentre, including Basaksehir Cam & Sakura City Hospital | To modify, validate, and compare a modified Naples Prognostic Score against other inflammatory indices for predicting in-hospital mortality in STEMI treated with primary PCI. |
| 116 | Zhang Y, Duan J, Huang F, Huang K, Wang R, Liu W, et al. Modified stress hyperglycemia ratio identifies the risk of early cardiovascular complications in patients with acute myocardial infarction | 2025 | USA and China – MIMIC-IV discovery cohort; external validation in China | To assess whether the modified stress hyperglycemia ratio predicts early cardiovascular complications in AMI and whether it outperforms related glycaemic indicators for risk stratification. |
| 117 | Soleimani A, et al. Predicting In-Hospital Mortality in Patients With Acute Myocardial Infarction: A Comparison of Machine Learning Approaches | 2025 | Iran – hospital-based clinical datasets | To develop and compare machine-learning models for in-hospital mortality prediction in patients with AMI. |
| 118 | Al Mamun MR, Rafi M, Chakraborty H, Anjum F, Gani R, Rashid MRA, et al. Cardiovascular Health Analysis: Machine Learning and Explainable AI to Predict Heart Attacks | 2025 | Bangladesh – East West University; secondary data from Zheen Hospital, Erbil, Iraq | To predict heart attacks and identify the most important cardiovascular indicators using machine learning and explainable AI. |
| 119 | Wang J-X, Liang M-M, Lu P-J, Cui Z, Liang Y, Wang Y-H, et al. Using Machine Learning to Predict MACEs Risk in Patients with Premature Myocardial Infarction | 2025 | China – Tianjin Chest Hospital / Tianjin Medical University | To develop an interpretable machine-learning model for long-term MACE risk assessment and stratification in premature MI and identify key prognostic factors. |
| 120 | Plasma Ceramides as Prognostic Biomarkers and Their Arterial and Myocardial Tissue Correlates in Acute Myocardial Infarction | 2018 | Singapore – two tertiary centres; New Zealand – Christchurch Hospital validation cohort | To evaluate the prognostic value of plasma ceramides in AMI for 12-month MACCE prediction and investigate their possible arterial and myocardial tissue correlates. |
