## Supplementary material for "Artificial Intelligence for Cardiac Biomarkers After Myocardial Infarction: A Systematic Review and a Leakage-Aware Modeling Framework": Table S4

**Supplementary Table S3.** Summary of risk of bias assessment using PROBAST

| **Domain** | **Low risk (%)** | **High risk (%)** | **Unclear (%)** | **Key issues identified** |
| --- | --- | --- | --- | --- |
| Participants | 62% | 18% | 20% | Selection bias in retrospective cohorts; limited reporting of inclusion criteria |
| Predictors | 35% | 45% | 20% | Lack of clear feature definition; inclusion of post-baseline variables; risk of information leakage |
| Outcome | 70% | 12% | 18% | Generally well-defined clinical endpoints; occasional unclear outcome timing |
| Analysis | 15% | 68% | 17% | Overfitting risk, lack of external validation, improper validation strategies, absence of calibration |
| **Overall** | **12%** | **72%** | **16%** | High risk mainly driven by analytical limitations and leakage concerns |
