## Supplementary material for "Artificial Intelligence for Cardiac Biomarkers After Myocardial Infarction: A Systematic Review and a Leakage-Aware Modeling Framework": Table S5

**Supplementary Table S4.** Certainty of evidence assessment using a modified GRADE framework

| **Outcome domain** | **Risk of bias** | **Inconsistency** | **Indirectness** | **Imprecision** | **Publication bias** | **Overall certainty** | **Key reasons for rating** |
| --- | --- | --- | --- | --- | --- | --- | --- |
| **Discrimination performance (e.g., ROC-AUC)** | Serious | Serious | Not serious | Serious | Possible | **Low** | High heterogeneity in study design and validation; overreliance on internal validation; wide variability in reported performance |
| **Calibration of prediction models** | Serious | Serious | Not serious | Serious | Possible | **Very low** | Calibration inconsistently reported; lack of standardized methods; limited availability of calibration metrics |
| **Clinical utility (e.g., decision curve analysis)** | Serious | Serious | Serious | Serious | Likely | **Very low** | Rare reporting of decision-analytic measures; unclear clinical applicability; indirect evidence |
| **Model validation and generalizability** | Serious | Serious | Not serious | Serious | Possible | **Low** | Limited external validation; predominance of internal validation strategies; unclear transportability |
| **Model interpretability and explainability** | Serious | Serious | Serious | Serious | Possible | **Very low** | Inconsistent use of explainability methods; lack of stability analysis; poor reporting |
| **Reproducibility and transparency (code/data availability)** | Serious | Serious | Serious | Serious | Likely | **Very low** | Low rate of code sharing; incomplete reporting; limited reproducibility practices |
