## Supplementary material for "Artificial Intelligence for Cardiac Biomarkers After Myocardial Infarction: A Systematic Review and a Leakage-Aware Modeling Framework": Table S6

**Table S5.** Data harmonization and type-coercion audit log

| **Variable - original name** | **Variable - harmonized name** | **Variable - English name** | **Raw type** | **Converted type** | **Non-parsable values** | **NA by coercion** |
| --- | --- | --- | --- | --- | --- | --- |
| 72 kDa pro-MMP-2 | 72_kda_pro_mmp_2 | 72 kDa pro-matrix metalloproteinase 2 | float64 | float64 | No | Yes |
| 92 kDa pro-MMP-9 | 92_kda_pro_mmp_9 | 92 kDa pro-matrix metalloproteinase 9 | float64 | float64 | No | Yes |
| ACE-I | ace_i | Angiotensin-converting-enzyme inhibitor | object | object | No | Yes |
| AlbumIny [g/L] | albuminy_g_l | Albumin [g/L] | float64 | float64 | No | Yes |
| ARB | arb | Angiotensin receptor blocker | object | object | No | Yes |
| ASA | asa | Acetylsalicylic acid | object | object | No | Yes |
| BB | bb | Beta-blocker | object | object | No | Yes |
| BMI | bmi | Body mass index | float64 | float64 | No | Yes |
| BMS/DES | bms_des | Bare-metal/drug-eluting stents | object | object | No | Yes |
| Brilique | brilique | Ticagrelor (Brilique) | object | object | No | Yes |
| (CAD) | cad | Coronary artery disease | object | object | No | Yes |
| CAD - ile lat? | cad_ile_lat | CAD duration (years) | object | object | No | Yes |
| CCS | ccs | Canadian Cardiovascular Society angina class | object | object | No | Yes |
| Cholesterol całkowity (TC) | cholesterol_calkowity_tc | Total cholesterol | float64 | float64 | No | Yes |
| CRP>5 zapalenie | crp_5_zapalenie | C-reactive protein >5 inflammation | object | object | No | Yes |
| CRT-D device | crt_d_device | cardiac resynchronization therapy defibrillator device | object | object | No | Yes |
| Cukrzyca (DM) | cukrzyca_dm | Diabetes mellitus | object | object | No | Yes |
| Czy pali? | czy_pali | Smoking status | object | object | No | No |
| Data CABG | data_cabg | Coronary artery bypass grafting date | object | object | No | Yes |
| Data obecnego PCI | data_obecnego_pci | Current percutaneous coronary intervention (PCI) date | object | object | No | Yes |
| Data obecnego zawału | data_obecnego_zawalu | Current MI date | object | object | No | Yes |
| Data pobrania | data_pobrania | Sample collection date | datetime64[ns] | datetime64[ns] | No | Yes |
| Data przeszłych zawałów | data_przeszlych_zawalow | Past MI dates | object | object | No | Yes |
| Data urodzenia | data_urodzenia | Date of birth | datetime64[ns] | datetime64[ns] | No | Yes |
| Data zapaenia żył | data_zapaenia_zyl | Vein inflammation date | object | object | No | Yes |
| Data zaplenie serca | data_zaplenie_serca | Myocarditis date | object | object | No | Yes |
| Data zatorowości płuc | data_zatorowosci_pluc | Pulmonary embolism date | object | object | No | Yes |
| Daty przeszłych PCI | daty_przeszlych_pci | Past PCI dates | object | object | No | Yes |
| DM - ile lat? | dm_ile_lat | Diabetes duration (years) | object | object | No | Yes |
| DM -leczenie | dm_leczenie | Diabetes treatment | object | object | No | Yes |
| EMMPRIN | emmprin | Extracellular matrix metalloproteinase inducer | float64 | float64 | No | Yes |
| Frakcja wyrzutowa lewej komory serca (LVEF) | frakcja_wyrzutowa_lewej_komory_serca_lvef | Left ventricular ejection fraction | float64 | float64 | No | Yes |
| GFR MIn. | gfr_min | Minimum glomerular filtration rate | float64 | float64 | No | Yes |
| Glukoza max. Mg/dl | glukoza_max_mg_dl | Maximum glucose (mg/dL) | float64 | float64 | No | Yes |
| Hb przyjęcie | hb_przyjecie | Hemoglobin on admission | float64 | float64 | No | Yes |
| Hb wypis | hb_wypis | Hemoglobin at discharge | float64 | float64 | No | Yes |
| HbA1C | hba1c | Hemoglobin A1c | float64 | float64 | No | Yes |
| HDL | hdl | High-density lipoprotein cholesterol | float64 | float64 | No | Yes |
| hsCRP max. | hscrp_max | Maximum high-sensitivity C-reactive protein | float64 | float64 | No | Yes |
| Ht przyjęcie | ht_przyjecie | Hematocrit on admission | float64 | float64 | No | Yes |
| Ht wypis | ht_wypis | Hematocrit at discharge | float64 | float64 | No | Yes |
| ICD implantable cardioverter defibrillator | icd_implantable_cardioverter_defibrillator | Implantable cardioverter defibrillator | object | object | No | Yes |
| Il-6 [pg/ml] | il_6_pg_ml | Interleukin-6 (pg/mL) | float64 | float64 | No | Yes |
| Ile lat? | ile_lat | Age (years) | object | object | No | Yes |
| Ile lat?.1 | ile_lat_1 | Age (years) 1 | float64 | float64 | No | Yes |
| Ile lat?.2 | ile_lat_2 | Age (years) 2 | object | object | No | Yes |
| Ile lat? CRT-D | ile_lat_crt_d | Cardiac resynchronization therapy defibrillator duration (years) | object | object | No | Yes |
| Ile lat? Miażdżyca t. szyjnych | ile_lat_miazdzyca_t_szyjnych | Carotid atherosclerosis duration | object | object | No | Yes |
| Ile lat? Miżdżyca kończyn | ile_lat_mizdzyca_konczyn | Peripheral atherosclerosis duration | float64 | float64 | No | Yes |
| Ile lat nie pali? | ile_lat_nie_pali | Years since quitting smoking | object | object | No | Yes |
| Ile lat pali? | ile_lat_pali | Smoking duration (years) | float64 | float64 | No | Yes |
| Ilość paczek dziennie | ilosc_paczek_dziennie | Packs per day | object | object | No | Yes |
| Internal ID | internal_id | Internal ID | object | object | No | Yes |
| Jaka? | jaka | Type | object | object | No | Yes |
| Jaka?.1 | jaka_1 | Type 1 | object | object | No | Yes |
| Kidnej Failure<15 ml/MIn/m2 | kidnej_failure_15_ml_min_m2 | Kidney failure <15 ml/min/m² | object | object | No | Yes |
| Kidnej insuficiency <60 ml/MIn/m2 | kidnej_insuficiency_60_ml_min_m2 | Kidney insufficiency <60 ml/min/m² | object | object | No | Yes |
| klop | klop | Clopidogrel | object | object | No | Yes |
| Krwawienie | krwawienie | Bleeding | object | object | No | Yes |
| Kwas moczowy | kwas_moczowy | Uric acid | float64 | float64 | No | Yes |
| LDL | ldl | Low-density lipoprotein cholesterol | float64 | float64 | No | Yes |
| Leukocytoza przyjęcie >10000 | leukocytoza_przyjecie_10000 | Leukocytosis on admission >10000 | object | object | No | Yes |
| MIażdżyca kończyn dolnych | miazdzyca_konczyn_dolnych | Peripheral artery disease | object | object | No | Yes |
| MIażdżyca tętnic szyjnych | miazdzyca_tetnic_szyjnych | Carotid artery disease | object | object | No | Yes |
| MIlurit/diuretyk | milurit_diuretyk | Milurit/diuretic | object | object | No | Yes |
| MPV przyjęcie | mpv_przyjecie | Mean platelet volume on admission | float64 | float64 | No | Yes |
| MPV wypis | mpv_wypis | Mean platelet volume at discharge | float64 | float64 | No | Yes |
| Nabyta wada serca | nabyta_wada_serca | Acquired heart disease | object | object | No | Yes |
| Nadciśnienie tętnicze (NT) | nadcisnienie_tetnicze_nt | Hypertension | object | object | No | Yes |
| NoAC | noac | Non-vitamin K antagonist oral anticoagulant | object | object | No | Yes |
| NoN-HDL(całkowity -hdl | non_hdl_calkowity_hdl | Non-HDL cholesterol | float64 | float64 | No | Yes |
| Nr na próbce | nr_na_probce | Sample number | object | object | No | Yes |
| Nr pacjenta | nr_pacjenta | Patient number | int64 | int64 | No | Yes |
| NYHA | nyha | New York Heart Association classification of heart failure | object | object | No | Yes |
| Obwód bioder | obwod_bioder | Hip circumference | float64 | float64 | No | Yes |
| Otyłość BMI >30 | otylosc_bmi_30 | Obesity BMI >30 | object | object | No | No |
| P-LCR przyjęcie | p_lcr_przyjecie | Platelet-large cell ratio on admission | float64 | float64 | No | Yes |
| P-LCR wypis | p_lcr_wypis | Platelet-large cell ratio at discharge | float64 | float64 | No | Yes |
| Patient ID | patient_id | Patient ID | object | object | No | Yes |
| PCI naczynie obecne | pci_naczynie_obecne | Current PCI vessel | object | object | No | Yes |
| PCT przyjęcie | pct_przyjecie | Plateletcrit on admission | float64 | float64 | No | Yes |
| PCT wypis | pct_wypis | Plateletcrit at discharge | float64 | float64 | No | Yes |
| PDW przyjęcie | pdw_przyjecie | Platelet distribution width on admission | float64 | float64 | No | Yes |
| PDW wypis | pdw_wypis | Platelet distribution width at discharge | float64 | float64 | No | Yes |
| Pętlowy | petlowy | Loop diuretic | object | object | No | Yes |
| Płeć | plec | Sex | object | object | No | Yes |
| PLT przyjęcie | plt_przyjecie | Platelets on admission | float64 | float64 | No | Yes |
| PLT wypis | plt_wypis | Platelets at discharge | float64 | float64 | No | Yes |
| Pomostowanie aortalNo-wieńcowe (CABG) | pomostowanie_aortalno_wiencowe_cabg | Coronary artery bypass grafting | object | object | No | Yes |
| Pomosty | pomosty | Grafts | object | object | No | Yes |
| Przewlekła niewydolNość serca (CHF) | przewlekla_niewydolnosc_serca_chf | Chronic heart failure | object | object | No | Yes |
| Przezskórna interwencja wieńcowa (PCI) | przezskorna_interwencja_wiencowa_pci | Percutaneous coronary intervention | object | object | No | Yes |
| RBC przyjęcie | rbc_przyjecie | Red blood cells on admission | float64 | float64 | No | Yes |
| RBC wypis | rbc_wypis | Red blood cells at discharge | float64 | float64 | No | Yes |
| Rodzaj zaburzeń | rodzaj_zaburzen | Type of disorder | object | object | No | Yes |
| Rodzeństwo | rodzenstwo | Siblings | object | object | No | Yes |
| Rodzice | rodzice | Parents | object | object | No | Yes |
| Statyna | statyna | Statin | object | object | No | Yes |
| Stymulator | stymulator | Pacemaker | object | object | No | Yes |
| TNFalfa [pg/ml] | tnfalfa_pg_ml | Tumour necrosis factor-alpha (pg/mL) | float64 | float64 | No | Yes |
| Trijglicerydy (TG) | trijglicerydy_tg | Triglycerides | float64 | float64 | No | Yes |
| Troponiny max. | troponiny_max | Maximum troponin | float64 | float64 | No | Yes |
| Typ zawału | typ_zawalu | MI type | object | object | No | Yes |
| Waga (kg) | waga_kg | Weight (kg) | float64 | float64 | No | Yes |
| WBC przyjęcie | wbc_przyjecie | White blood cells on admission | float64 | float64 | No | Yes |
| WBC wypis | wbc_wypis | White blood cells at discharge | float64 | float64 | No | Yes |
| Wiek w momencie zbiórki | wiek_w_momencie_zbiorki | Age at collection | int64 | int64 | No | Yes |
| Wrodzona wada serca | wrodzona_wada_serca | Congenital heart defect | object | object | No | Yes |
| Wyniki FGF23 pg/ml | wyniki_fgf23_pg_ml | Fibroblast growth factor 23 (pg/mL) | float64 | float64 | No | Yes |
| Wyniki Klotho pg/ml | wyniki_klotho_pg_ml | Klotho (pg/mL) | float64 | float64 | No | Yes |
| Wzrost (cm) | wzrost_cm | Height (cm) | float64 | float64 | No | Yes |
| Zaburzenia homeostazy glukozy >6 | zaburzenia_homeostazy_glukozy_6 | Glucose homeostasis disorder >6 | object | object | No | Yes |
| Zaburzenia rytmu | zaburzenia_rytmu | Arrhythmia | object | object | No | Yes |
| Zapalenie MIęśnia serca | zapalenie_miesnia_serca | Myocarditis | object | object | No | Yes |
| Zapalenie żył kończyn dolnych/ Zakrzepica | zapalenie_zyl_konczyn_dolnych_zakrzepica | Deep vein thrombosis | object | object | No | Yes |
| Zatorowość płucna | zatorowosc_plucna | Pulmonary embolism | object | object | No | Yes |
| Zawał: 0 nigdy, 1 teraz, 2 kiedyś, 3 kiedyś i teraz, a- steMI, b-nsteMI. c- ua | zawal_0_nigdy_1_teraz_2_kiedys_3_kiedys_i_teraz_a_stemi_b_nstemi_c_ua | MI classification | object | object | No | Yes |
