## Supplementary material for "Artificial Intelligence for Cardiac Biomarkers After Myocardial Infarction: A Systematic Review and a Leakage-Aware Modeling Framework": Table S7

**.** Dataset structure and missingness profile after governance preprocessing

| **Parameter** | **Before governance, N = 57** | **After governance, N = 57** |
| --- | --- | --- |
| **Typ zawału (Type of heart attack)** |  |  |
| NSTEMI | 32 (56.1%) | 32 (56.1%) |
| STEMI | 25 (43.9%) | 25 (43.9%) |
| **Nr na próbce**  **(No. on the sample)** |  |  |
| Data completed | 57 (100.0%) | 57 (100.0%) |
| **Typ zawału**  **(Type of heart attack)** |  |  |
| Data completed | 57 (100.0%) | 57 (100.0%) |
| **Data pobrania**  **(Date of download)** |  |  |
| Data completed | 57 (100.0%) | 57 (100.0%) |
| **Płeć (Sex)** |  |  |
| Data completed | 57 (100.0%) | 57 (100.0%) |
| **Data urodzenia**  **(Date of birth)** |  |  |
| Data completed | 57 (100.0%) | 57 (100.0%) |
| **EMMPRIN** |  |  |
| Data completed | 57 (100.0%) | 57 (100.0%) |
| **92 kDa pro-MMP-9** |  |  |
| Data completed | 55 (96.5%) | 55 (96.5%) |
| Missing data | 2 (3.5%) | 2 (3.5%) |
| **72 kDa pro-MMP-2** |  |  |
| Data completed | 48 (84.2%) | 48 (84.2%) |
| Missing data | 9 (15.8%) | 9 (15.8%) |
| **Wyniki Klotho pg/ml**  **(Klotho results pg/ml)** |  |  |
| Data completed | 57 (100.0%) | 57 (100.0%) |
| **Wyniki FGF23 pg/ml**  **(FGF23 pg/ml results)** |  |  |
| Data completed | 55 (96.5%) | 55 (96.5%) |
| Missing data | 2 (3.5%) | 2 (3.5%) |
| **TNFalfa [pg/ml]** |  |  |
| Data completed | 56 (98.2%) | 56 (98.2%) |
| Missing data | 1 (1.8%) | 1 (1.8%) |
| **Il-6 [pg/ml]** |  |  |
| Data completed | 56 (98.2%) | 56 (98.2%) |
| Missing data | 1 (1.8%) | 1 (1.8%) |
| **Wiek w momencie zbiórki**  **(Age at collection)** |  |  |
| Data completed | 57 (100.0%) | 57 (100.0%) |
| **Waga (kg)**  **(Weight (kg))** |  |  |
| Data completed | 57 (100.0%) | 57 (100.0%) |
| **Wzrost (cm)**  **(Height (cm))** |  |  |
| Data completed | 57 (100.0%) | 57 (100.0%) |
| **BMI** |  |  |
| Data completed | 57 (100.0%) | 57 (100.0%) |
| **Obwód bioder**  **(Hip circumference)** |  |  |
| Data completed | 54 (94.7%) | 54 (94.7%) |
| Missing data | 3 (5.3%) | 3 (5.3%) |
| **Otyłość BMI >30**  **(Obesity BMI >30)** |  |  |
| Data completed | 57 (100.0%) | 57 (100.0%) |
| **Czy pali?**  **(Does it smoke?)** |  |  |
| Data completed | 56 (98.2%) | 56 (98.2%) |
| Missing data | 1 (1.8%) | 1 (1.8%) |
| **Ile lat pali?**  **(How many years has he been smoking?)** |  |  |
| Data completed | 18 (31.6%) | 18 (31.6%) |
| Missing data | 39 (68.4%) | 39 (68.4%) |
| **Ilość paczek dziennie**  **(Number of packages per day)** |  |  |
| Data completed | 17 (29.8%) | 17 (29.8%) |
| Missing data | 40 (70.2%) | 40 (70.2%) |
| **Ile lat nie pali?**  **(How many years has he not smoked?)** |  |  |
| Data completed | 18 (31.6%) | 18 (31.6%) |
| Missing data | 39 (68.4%) | 39 (68.4%) |
| **Rodzice**  **(Parents)** |  |  |
| Data completed | 56 (98.2%) | 56 (98.2%) |
| Missing data | 1 (1.8%) | 1 (1.8%) |
| **Rodzeństwo**  **(Siblings)** |  |  |
| Data completed | 55 (96.5%) | 55 (96.5%) |
| Missing data | 2 (3.5%) | 2 (3.5%) |
| **Cukrzyca (DM)**  **(Diabetes mellitus (DM))** |  |  |
| Data completed | 57 (100.0%) | 57 (100.0%) |
| **DM - ile lat?**  **(DM - how many years does it last?)** |  |  |
| Data completed | 13 (22.8%) | 13 (22.8%) |
| Missing data | 44 (77.2%) | 44 (77.2%) |
| **DM -leczenie**  **(DM - treatment)** |  |  |
| Data completed | 19 (33.3%) | 19 (33.3%) |
| Missing data | 38 (66.7%) | 38 (66.7%) |
| **(CAD)** |  |  |
| Data completed | 0 (0.0%) | 57 (100.0%) |
| **CAD - ile lat?**  **(CAD - how many years does it last?)** |  |  |
| Data completed | 28 (49.1%) | 28 (49.1%) |
| Missing data | 29 (50.9%) | 29 (50.9%) |
| **CCS** |  |  |
| Data completed | 21 (36.8%) | 21 (36.8%) |
| Missing data | 36 (63.2%) | 36 (63.2%) |
| **Zawał: 0 nigdy, 1 teraz, 2 kiedyś, 3 kiedyś i teraz, a- steMI, b-nsteMI. c- ua**  **(Heart attack: 0 never, 1 now, 2 sometime, 3 sometime and now, a- steMI, b-nsteMI. c- ua)** |  |  |
| Data completed | 57 (100.0%) | 57 (100.0%) |
| **Data obecnego zawału**  **(Date of current heart attack)** |  |  |
| Data completed | 48 (84.2%) | 48 (84.2%) |
| Missing data | 9 (15.8%) | 9 (15.8%) |
| **Data przeszłych zawałów**  **(Date of past heart attacks)** |  |  |
| Data completed | 22 (38.6%) | 22 (38.6%) |
| Missing data | 35 (61.4%) | 35 (61.4%) |
| **Przezskórna interwencja wieńcowa (PCI)**  **(Percutaneous coronary intervention (PCI))** |  |  |
| Data completed | 57 (100.0%) | 57 (100.0%) |
| **Data obecnego PCI**  **(Current PCI Date)** |  |  |
| Data completed | 36 (63.2%) | 36 (63.2%) |
| Missing data | 21 (36.8%) | 21 (36.8%) |
| **Daty przeszłych PCI**  **(Past PCI Dates)** |  |  |
| Data completed | 14 (24.6%) | 14 (24.6%) |
| Missing data | 43 (75.4%) | 43 (75.4%) |
| **PCI naczynie obecne**  **(PCI vessel present)** |  |  |
| Data completed | 39 (68.4%) | 39 (68.4%) |
| Missing data | 18 (31.6%) | 18 (31.6%) |
| **BMS/DES** |  |  |
| Data completed | 39 (68.4%) | 39 (68.4%) |
| Missing data | 18 (31.6%) | 18 (31.6%) |
| **Pomostowanie aortalNo-wieńcowe (CABG)**  **(Coronary artery bypass grafting (CABG))** |  |  |
| Data completed | 57 (100.0%) | 57 (100.0%) |
| **Data CABG**  **(CABG Date)** |  |  |
| Data completed | 5 (8.8%) | 5 (8.8%) |
| Missing data | 52 (91.2%) | 52 (91.2%) |
| **Pomosty**  **(Bypass)** |  |  |
| Data completed | 3 (5.3%) | 3 (5.3%) |
| Missing data | 54 (94.7%) | 54 (94.7%) |
| **Wrodzona wada serca**  **(Congenital heart defect)** |  |  |
| Data completed | 57 (100.0%) | 57 (100.0%) |
| **Jaka?**  **(How?)** |  |  |
| Data completed | 1 (1.8%) | 1 (1.8%) |
| Missing data | 56 (98.2%) | 56 (98.2%) |
| **Nabyta wada serca**  **(Acquired heart defect)** |  |  |
| Data completed | 57 (100.0%) | 57 (100.0%) |
| **Jaka?**  **(How?)** |  |  |
| Data completed | 41 (71.9%) | 41 (71.9%) |
| Missing data | 16 (28.1%) | 16 (28.1%) |
| **Zapalenie MIęśnia serca**  **(Inflammation of the heart muscle)** |  |  |
| Data completed | 57 (100.0%) | 57 (100.0%) |
| **Data zaplenie serca**  **(Date of myocarditis)** |  |  |
| Data completed | 1 (1.8%) | 1 (1.8%) |
| Missing data | 56 (98.2%) | 56 (98.2%) |
| **Nadciśnienie tętnicze (NT)**  **(Arterial hypertension (HT))** |  |  |
| Data completed | 57 (100.0%) | 57 (100.0%) |
| **Ile lat?**  **(How many years?)** |  |  |
| Data completed | 34 (59.6%) | 34 (59.6%) |
| Missing data | 23 (40.4%) | 23 (40.4%) |
| **Przewlekła niewydolNość serca (CHF)**  **(Chronic heart failure (CHF))** |  |  |
| Data completed | 56 (98.2%) | 56 (98.2%) |
| Missing data | 1 (1.8%) | 1 (1.8%) |
| **Ile lat?**  **(How many years?)** |  |  |
| Data completed | 10 (17.5%) | 10 (17.5%) |
| Missing data | 47 (82.5%) | 47 (82.5%) |
| **NYHA** |  |  |
| Data completed | 28 (49.1%) | 28 (49.1%) |
| Missing data | 29 (50.9%) | 29 (50.9%) |
| **Frakcja wyrzutowa lewej komory serca (LVEF)**  **(Left ventricular ejection fraction (LVEF))** |  |  |
| Data completed | 56 (98.2%) | 56 (98.2%) |
| Missing data | 1 (1.8%) | 1 (1.8%) |
| **Zaburzenia rytmu**  **(Arrhythmias)** |  |  |
| Data completed | 57 (100.0%) | 57 (100.0%) |
| **Rodzaj zaburzeń**  **(Type of disorder)** |  |  |
| Data completed | 20 (35.1%) | 20 (35.1%) |
| Missing data | 37 (64.9%) | 37 (64.9%) |
| **Ile lat?**  **(How many years?)** |  |  |
| Data completed | 8 (14.0%) | 8 (14.0%) |
| Missing data | 49 (86.0%) | 49 (86.0%) |
| **MIażdżyca kończyn dolnych**  **(Atherosclerosis of the lower limbs)** |  |  |
| Data completed | 56 (98.2%) | 56 (98.2%) |
| Missing data | 1 (1.8%) | 1 (1.8%) |
| **Ile lat? Miażdżyca kończyn**  **(How old? Atherosclerosis of the limbs)** |  |  |
| Data completed | 4 (7.0%) | 4 (7.0%) |
| Missing data | 53 (93.0%) | 53 (93.0%) |
| **MIażdżyca tętnic szyjnych**  **(Carotid atherosclerosis)** |  |  |
| Data completed | 55 (96.5%) | 55 (96.5%) |
| Missing data | 2 (3.5%) | 2 (3.5%) |
| **Ile lat? Miażdżyca t. Szyjnych**  **(How old are you? Cervical atherosclerosis)** |  |  |
| Data completed | 3 (5.3%) | 3 (5.3%) |
| Missing data | 54 (94.7%) | 54 (94.7%) |
| **Zapalenie żył kończyn dolnych/ Zakrzepica**  **(Lower limb phlebitis/Thrombosis)** |  |  |
| Data completed | 56 (98.2%) | 56 (98.2%) |
| Missing data | 1 (1.8%) | 1 (1.8%) |
| **Data zapaenia żył**  **(Date of phlebitis)** |  |  |
| Data completed | 2 (3.5%) | 2 (3.5%) |
| Missing data | 55 (96.5%) | 55 (96.5%) |
| **Zatorowość płucna**  **(Pulmonary embolism)** |  |  |
| Data completed | 55 (96.5%) | 55 (96.5%) |
| Missing data | 2 (3.5%) | 2 (3.5%) |
| **Data zatorowości płuc**  **(Date of pulmonary embolism)** |  |  |
| Data completed | 2 (3.5%) | 2 (3.5%) |
| Missing data | 55 (96.5%) | 55 (96.5%) |
| **Stymulator**  **(Stimulator)** |  |  |
| Data completed | 57 (100.0%) | 57 (100.0%) |
| **ICD implantable cardioverter defibrillator** |  |  |
| Data completed | 57 (100.0%) | 57 (100.0%) |
| **CRT-D device** |  |  |
| Data completed | 57 (100.0%) | 57 (100.0%) |
| **Ile lat? CRT-D**  **(How old is CRT-D?)** |  |  |
| Data completed | 8 (14.0%) | 8 (14.0%) |
| Missing data | 49 (86.0%) | 49 (86.0%) |
| **GFR MIn.** |  |  |
| Data completed | 57 (100.0%) | 57 (100.0%) |
| **Kidnej Failure<15 ml/MIn/m2** |  |  |
| Missing data | 57 (100.0%) | 57 (100.0%) |
| **Kidnej insuficiency <60 ml/MIn/m2** |  |  |
| Missing data | 57 (100.0%) | 57 (100.0%) |
| **Troponiny max.** |  |  |
| Data completed | 57 (100.0%) | 57 (100.0%) |
| **Cholesterol całkowity (TC)**  **(Total cholesterol (TC))** |  |  |
| Data completed | 57 (100.0%) | 57 (100.0%) |
| **LDL** |  |  |
| Data completed | 57 (100.0%) | 57 (100.0%) |
| **HDL** |  |  |
| Data completed | 57 (100.0%) | 57 (100.0%) |
| **Trijglicerydy (TG)** |  |  |
| Data completed | 57 (100.0%) | 57 (100.0%) |
| **NoN-HDL(całkowity -hdl** |  |  |
| Data completed | 57 (100.0%) | 57 (100.0%) |
| **Kwas moczowy**  **(Uric acid)** |  |  |
| Data completed | 35 (61.4%) | 35 (61.4%) |
| Missing data | 22 (38.6%) | 22 (38.6%) |
| **HbA1C** |  |  |
| Data completed | 57 (100.0%) | 57 (100.0%) |
| **Zaburzenia homeostazy glukozy >6**  **(Glucose homeostasis disorders >6)** |  |  |
| Data completed | 57 (100.0%) | 57 (100.0%) |
| **Glukoza max. Mg/dl**  **(Glucose max Mg/dl)** |  |  |
| Data completed | 57 (100.0%) | 57 (100.0%) |
| **hsCRP max.** |  |  |
| Data completed | 57 (100.0%) | 57 (100.0%) |
| **CRP>5 zapalenie**  **(CRP>5 inflammation)** |  |  |
| Data completed | 57 (100.0%) | 57 (100.0%) |
| **AlbumIny [g/L]** |  |  |
| Missing data | 57 (100.0%) | 57 (100.0%) |
| **RBC przyjęcie**  **(RBC admission to the ward)** |  |  |
| Data completed | 57 (100.0%) | 57 (100.0%) |
| **Ht przyjęcie**  **(Ht admission to the ward)** |  |  |
| Data completed | 57 (100.0%) | 57 (100.0%) |
| **Hb przyjęcie**  **(Hb admission to the ward)** |  |  |
| Data completed | 57 (100.0%) | 57 (100.0%) |
| **WBC przyjęcie**  **(WBC admission to the ward)** |  |  |
| Data completed | 57 (100.0%) | 57 (100.0%) |
| **Leukocytoza przyjęcie >10000**  **(Leukocytosis admission >10000)** |  |  |
| Data completed | 57 (100.0%) | 57 (100.0%) |
| **MPV przyjęcie**  **(MPV admission to the ward)** |  |  |
| Data completed | 56 (98.2%) | 56 (98.2%) |
| Missing data | 1 (1.8%) | 1 (1.8%) |
| **PCT przyjęcie**  **(PCT admission to the ward)** |  |  |
| Data completed | 56 (98.2%) | 56 (98.2%) |
| Missing data | 1 (1.8%) | 1 (1.8%) |
| **PDW przyjęcie**  **(PDW admission to the ward)** |  |  |
| Data completed | 56 (98.2%) | 56 (98.2%) |
| Missing data | 1 (1.8%) | 1 (1.8%) |
| **P-LCR przyjęcie**  **(P-LCR admission to the ward)** |  |  |
| Data completed | 56 (98.2%) | 56 (98.2%) |
| Missing data | 1 (1.8%) | 1 (1.8%) |
| **PLT przyjęcie**  **(PLT admission to the ward)** |  |  |
| Data completed | 57 (100.0%) | 57 (100.0%) |
| **RBC wypis**  **(RBC extract)** |  |  |
| Data completed | 57 (100.0%) | 57 (100.0%) |
| **Ht wypis**  **(Ht extract)** |  |  |
| Data completed | 57 (100.0%) | 57 (100.0%) |
| **Hb wypis**  **(Hb extract)** |  |  |
| Data completed | 57 (100.0%) | 57 (100.0%) |
| **WBC wypis**  **(WBC extract)** |  |  |
| Data completed | 57 (100.0%) | 57 (100.0%) |
| **MPV wypis**  **(MPV extract)** |  |  |
| Data completed | 56 (98.2%) | 56 (98.2%) |
| Missing data | 1 (1.8%) | 1 (1.8%) |
| **PCT wypis**  **(PCT extract)** |  |  |
| Data completed | 56 (98.2%) | 56 (98.2%) |
| Missing data | 1 (1.8%) | 1 (1.8%) |
| **PDW wypis**  **(PDW extract)** |  |  |
| Data completed | 56 (98.2%) | 56 (98.2%) |
| Missing data | 1 (1.8%) | 1 (1.8%) |
| **P-LCR wypis**  **(P-LCR extract)** |  |  |
| Data completed | 56 (98.2%) | 56 (98.2%) |
| Missing data | 1 (1.8%) | 1 (1.8%) |
| **PLT wypis**  **(PLT extract)** |  |  |
| Data completed | 56 (98.2%) | 56 (98.2%) |
| Missing data | 1 (1.8%) | 1 (1.8%) |
| **ASA** |  |  |
| Data completed | 57 (100.0%) | 57 (100.0%) |
| **klop** |  |  |
| Data completed | 57 (100.0%) | 57 (100.0%) |
| **NoAC** |  |  |
| Data completed | 57 (100.0%) | 57 (100.0%) |
| **ACE-I** |  |  |
| Data completed | 57 (100.0%) | 57 (100.0%) |
| **ARB** |  |  |
| Data completed | 57 (100.0%) | 57 (100.0%) |
| **BB** |  |  |
| Data completed | 57 (100.0%) | 57 (100.0%) |
| **Statyna**  **(Statin)** |  |  |
| Data completed | 57 (100.0%) | 57 (100.0%) |
| **MIlurit/diuretyk** |  |  |
| Data completed | 56 (98.2%) | 56 (98.2%) |
| Missing data | 1 (1.8%) | 1 (1.8%) |
| **Pętlowy**  **(Loop)** |  |  |
| Data completed | 57 (100.0%) | 57 (100.0%) |
| **Krwawienie**  **(Bleeding)** |  |  |
| Data completed | 57 (100.0%) | 57 (100.0%) |
| **Brilique** |  |  |
| Data completed | 57 (100.0%) | 57 (100.0%) |
| ** (CAD)** |  |  |
| Data completed | 57 (100.0%) | 0 (0.0%) |
| Median shortage rate |  |  |
| Average shortage rate | 20.4 | 20.4 |
| Median shortage rate | 0.9 | 0.9 |
| **Number of variables with >50% missing values** | 25 | 25 |
