## Supplementary material for "Artificial Intelligence for Cardiac Biomarkers After Myocardial Infarction: A Systematic Review and a Leakage-Aware Modeling Framework": Table S8

**Table S7.** Feature-set variant composition and governance summary

| **Parameter** | **FULL** | **CLINICAL** | **BIOMARKERS** |
| --- | --- | --- | --- |
| **Features count** | 73 | 62 | 11 |
| **Features** | 72_kda_pro_mmp_2 | ace_i | 72_kda_pro_mmp_2 |
|  | 92_kda_pro_mmp_9 | albuminy_g_l | 92_kda_pro_mmp_9 |
|  | ace_i | arb | cholesterol_calkowity_tc |
|  | albuminy_g_l | bb | emmprin |
|  | arb | bmi | hdl |
|  | bb | bms_des | il_6_pg_ml |
|  | bmi | cad | ldl |
|  | bms_des | ccs | non_hdl_calkowity_hdl |
|  | cad | crp_5_zapalenie | tnfalfa_pg_ml |
|  | ccs | crt_d_device | wyniki_fgf23_pg_ml |
|  | cholesterol_calkowity_tc | cukrzyca_dm | wyniki_klotho_pg_ml |
|  | crp_5_zapalenie | czy_pali |  |
|  | crt_d_device | dm_leczenie |  |
|  | cukrzyca_dm | gfr_min |  |
|  | czy_pali | hb_przyjecie |  |
|  | dm_leczenie | hba1c |  |
|  | emmprin | ht_przyjecie |  |
|  | gfr_min | ile_lat_1 |  |
|  | hb_przyjecie | ile_lat_miazdzyca_t_szyjnych |  |
|  | hba1c | ile_lat_mizdzyca_konczyn |  |
|  | hdl | ile_lat_pali |  |
|  | ht_przyjecie | jaka |  |
|  | il_6_pg_ml | jaka_1 |  |
|  | ile_lat_1 | kidnej_failure_15_ml_min_m2 |  |
|  | ile_lat_miazdzyca_t_szyjnych | kidnej_insuficiency_60_ml_min_m2 |  |
|  | ile_lat_mizdzyca_konczyn | klop |  |
|  | ile_lat_pali | krwawienie |  |
|  | jaka | kwas_moczowy |  |
|  | jaka_1 | leukocytoza_przyjecie_10000 |  |
|  | kidnej_failure_15_ml_min_m2 | miazdzyca_konczyn_dolnych |  |
|  | kidnej_insuficiency_60_ml_min_m2 | miazdzyca_tetnic_szyjnych |  |
|  | klop | mpv_przyjecie |  |
|  | krwawienie | nabyta_wada_serca |  |
|  | kwas_moczowy | nadcisnienie_tetnicze_nt |  |
|  | ldl | noac |  |
|  | leukocytoza_przyjecie_10000 | nyha |  |
|  | miazdzyca_konczyn_dolnych | obwod_bioder |  |
|  | miazdzyca_tetnic_szyjnych | otylosc_bmi_30 |  |
|  | mpv_przyjecie | p_lcr_przyjecie |  |
|  | nabyta_wada_serca | pct_przyjecie |  |
|  | nadcisnienie_tetnicze_nt | pdw_przyjecie |  |
|  | noac | petlowy |  |
|  | non_hdl_calkowity_hdl | plec |  |
|  | nyha | plt_przyjecie |  |
|  | obwod_bioder | pomosty |  |
|  | otylosc_bmi_30 | przewlekla_niewydolnosc_serca_chf |  |
|  | p_lcr_przyjecie | rbc_przyjecie |  |
|  | pct_przyjecie | rodzaj_zaburzen |  |
|  | pdw_przyjecie | rodzenstwo |  |
|  | petlowy | rodzice |  |
|  | plec | stymulator |  |
|  | plt_przyjecie | trijglicerydy_tg |  |
|  | pomosty | waga_kg |  |
|  | przewlekla_niewydolnosc_serca_chf | wbc_przyjecie |  |
|  | rbc_przyjecie | wiek_w_momencie_zbiorki |  |
|  | rodzaj_zaburzen | wrodzona_wada_serca |  |
|  | rodzenstwo | wzrost_cm |  |
|  | rodzice | zaburzenia_homeostazy_glukozy_6 |  |
|  | stymulator | zaburzenia_rytmu |  |
|  | tnfalfa_pg_ml | zapalenie_miesnia_serca |  |
|  | trijglicerydy_tg | zapalenie_zyl_konczyn_dolnych_zakrzepica |  |
|  | waga_kg | zatorowosc_plucna |  |
|  | wbc_przyjecie |  |  |
|  | wiek_w_momencie_zbiorki |  |  |
|  | wrodzona_wada_serca |  |  |
|  | wyniki_fgf23_pg_ml |  |  |
|  | wyniki_klotho_pg_ml |  |  |
|  | wzrost_cm |  |  |
|  | zaburzenia_homeostazy_glukozy_6 |  |  |
|  | zaburzenia_rytmu |  |  |
|  | zapalenie_miesnia_serca |  |  |
|  | zapalenie_zyl_konczyn_dolnych_zakrzepica |  |  |
|  | zatorowosc_plucna |  |  |
| **Reasons for excluding variables** |  |  |  |
| all_missing_in_df_task / missing_gt_0.95 | 4 | 4 |  |
| triage_auc_ge_0.98 |  |  | 1 |
