## Supplementary material for "Artificial Intelligence for Cardiac Biomarkers After Myocardial Infarction: A Systematic Review and a Leakage-Aware Modeling Framework": Table S9

**Table S4.** Nested cross-validation performance summary (OOF-based)

| **Parameter** | **FULL** | **CLINICAL** | **BIOMARKERS** |
| --- | --- | --- | --- |
| Winner model | RF | RF | RF |
| n_features | 69 | 58 | 10 |
| ROC AUC (mean ± SD) | 0.994 ± 0.013 | 0.594 ± 0.115 | 0.950 ± 0.051 |
| PR AUC (mean ± SD) | 0.993 ± 0.015 | 0.616 ± 0.157 | 0.934 ± 0.074 |
| Brier score (mean ± SD) | 0.078 ± 0.057 | 0.237 ± 0.010 | 0.111 ± 0.031 |
