## Supplementary material for "Artificial Intelligence for Cardiac Biomarkers After Myocardial Infarction: A Systematic Review and a Leakage-Aware Modeling Framework": Table S10

**Table S9.** Winner model selection by feature-set variant

| **Feature-set variant** | **Winner model** | **AUC (mean ± SD)** | **MCC (mean ± SD)** | **n_features** | **RFE** | **Calibration** |
| --- | --- | --- | --- | --- | --- | --- |
| BIOMARKERS | RF | 0.95 ± 0.051 | 0.756 ± 0.164 | 10 | No | isotonic |
| CLINICAL | RF | 0.582 ± 0.157 | 0.08 ± 0.364 | 42 | Yes (top 10) | isotonic |
| FULL | RF | 0.994 ± 0.013 | 0.931 ± 0.153 | 53 | No | isotonic |
