## Supplementary material for "Artificial Intelligence for Cardiac Biomarkers After Myocardial Infarction: A Systematic Review and a Leakage-Aware Modeling Framework": Table S11

**Table S10.** Winner-model OOF performance with bootstrap confidence intervals

| **Parameter** | **FULL** | **CLINICAL** | **BIOMARKERS** |
| --- | --- | --- | --- |
| ROC AUC [95% CI] | 0.99 [0.98–1.01] | 0.59 [0.49–0.70] | 0.95 [0.91–0.99] |
| PR AUC [95% CI] | 0.99 [0.98–1.01] | 0.62 [0.48–0.75] | 0.93 [0.87–1.00] |
| Brier score [95% CI] | 0.08 [0.03–0.13] | 0.24 [0.23–0.25] | 0.11 [0.08–0.14] |
