## Supplementary material for "Artificial Intelligence for Cardiac Biomarkers After Myocardial Infarction: A Systematic Review and a Leakage-Aware Modeling Framework": Table S12

**Table S11.** Cross-fitted calibration and threshold-selection summary

| **Calibration method** | **Selected threshold** | **FULL** | **CLINICAL** | **BIOMARKERS** |
| --- | --- | --- | --- | --- |
| cal_intercept | f1 | 0.20 | -0.18 | 0.35 |
| cal_intercept | f1 | 0.21 | -0.07 | -0.04 |
| cal_intercept | f1 | 0.31 | -0.15 | -0.24 |
| cal_intercept | mcc | 0.20 | -0.18 | 0.35 |
| cal_intercept | mcc | 0.21 | -0.07 | -0.04 |
| cal_intercept | mcc | 0.31 | -0.15 | -0.24 |
| cal_intercept | youden | 0.20 | -0.18 | 0.35 |
| cal_intercept | youden | 0.21 | -0.07 | -0.04 |
| cal_intercept | youden | 0.31 | -0.15 | -0.24 |
| cal_slope | f1 | 0.37 | 0.06 | 0.15 |
| cal_slope | f1 | 1.99 | 0.70 | 1.12 |
| cal_slope | f1 | 2.68 | 0.81 | 1.76 |
| cal_slope | mcc | 0.37 | 0.06 | 0.15 |
| cal_slope | mcc | 1.99 | 0.70 | 1.12 |
| cal_slope | mcc | 2.68 | 0.81 | 1.76 |
| cal_slope | youden | 0.37 | 0.06 | 0.15 |
| cal_slope | youden | 1.99 | 0.70 | 1.12 |
| cal_slope | youden | 2.68 | 0.81 | 1.76 |
| calibration | f1 | isotonic_crossfit | isotonic_crossfit | isotonic_crossfit |
| calibration | f1 | platt_crossfit | platt_crossfit | platt_crossfit |
| calibration | f1 | uncalibrated | uncalibrated | uncalibrated |
| calibration | mcc | isotonic_crossfit | isotonic_crossfit | isotonic_crossfit |
| calibration | mcc | platt_crossfit | platt_crossfit | platt_crossfit |
| calibration | mcc | uncalibrated | uncalibrated | uncalibrated |
| calibration | youden | isotonic_crossfit | isotonic_crossfit | isotonic_crossfit |
| calibration | youden | platt_crossfit | platt_crossfit | platt_crossfit |
| calibration | youden | uncalibrated | uncalibrated | uncalibrated |
| ece | f1 | 0.03 | 0.08 | 0.08 |
| ece | f1 | 0.09 | 0.04 | 0.09 |
| ece | f1 | 0.21 | 0.03 | 0.11 |
| ece | mcc | 0.03 | 0.08 | 0.08 |
| ece | mcc | 0.09 | 0.04 | 0.09 |
| ece | mcc | 0.21 | 0.03 | 0.11 |
| ece | youden | 0.03 | 0.08 | 0.08 |
| ece | youden | 0.09 | 0.04 | 0.09 |
| ece | youden | 0.21 | 0.03 | 0.11 |
| mce | f1 | 0.35 | 0.25 | 0.41 |
| mce | f1 | 0.28 | 0.08 | 0.16 |
| mce | f1 | 0.30 | 0.04 | 0.17 |
| mce | mcc | 0.35 | 0.25 | 0.41 |
| mce | mcc | 0.28 | 0.08 | 0.16 |
| mce | mcc | 0.30 | 0.04 | 0.17 |
| mce | youden | 0.35 | 0.25 | 0.41 |
| mce | youden | 0.28 | 0.08 | 0.16 |
| mce | youden | 0.30 | 0.04 | 0.17 |
