## Supplementary material for "Artificial Intelligence for Cardiac Biomarkers After Myocardial Infarction: A Systematic Review and a Leakage-Aware Modeling Framework": Table S13

**Table S12.** Performance–complexity summary for winner models (variant with net_benefit)

| **Variant** | **Model** | **Calibration metrics** | **N** | **ROC AUC** | **PR AUC** | **Brier** | **ROC AUC (OOF)** | **PR AUC (OOF)** | **Brier (OOF)** | **DCA indicator (max net benefit)** |
| --- | --- | --- | --- | --- | --- | --- | --- | --- | --- | --- |
| BIOMARKERS | RF | isotonic_crossfit | 57 | 0.9 | 0.82 | 0.09 | 0.93 | 0.89 | 0.11 | 0.43 |
| BIOMARKERS | RF | platt_crossfit | 57 | 0.93 | 0.89 | 0.1 | 0.93 | 0.89 | 0.11 | 0.43 |
| BIOMARKERS | RF | uncalibrated | 57 | 0.93 | 0.89 | 0.11 | 0.93 | 0.89 | 0.11 | 0.43 |
| CLINICAL | RF | isotonic_crossfit | 57 | 0.53 | 0.5 | 0.26 | 0.6 | 0.58 | 0.24 | 0.43 |
| CLINICAL | RF | platt_crossfit | 57 | 0.6 | 0.58 | 0.24 | 0.6 | 0.58 | 0.24 | 0.43 |
| CLINICAL | RF | uncalibrated | 57 | 0.6 | 0.58 | 0.24 | 0.6 | 0.58 | 0.24 | 0.43 |
| FULL | RF | isotonic_crossfit | 57 | 1 | 1 | 0.02 | 1 | 1 | 0.08 | 0.43 |
| FULL | RF | platt_crossfit | 57 | 1 | 1 | 0.03 | 1 | 1 | 0.08 | 0.43 |
| FULL | RF | uncalibrated | 57 | 1 | 1 | 0.08 | 1 | 1 | 0.08 | 0.43 |
