## Supplementary material for "Artificial Intelligence for Cardiac Biomarkers After Myocardial Infarction: A Systematic Review and a Leakage-Aware Modeling Framework": Table S14

**Table S13.** Regularization-based feature reduction summary

| **Variant** | **Model** | **roc_auc** | **N** | **Calibration** |
| --- | --- | --- | --- | --- |
| BIOMARKERS | RF | 0.8975 | 57 | isotonic_crossfit |
| BIOMARKERS | RF | 0.9263 | 57 | platt_crossfit |
| BIOMARKERS | RF | 0.93 | 57 | uncalibrated |
| CLINICAL | RF | 0.5319 | 57 | isotonic_crossfit |
| CLINICAL | RF | 0.6038 | 57 | platt_crossfit |
| CLINICAL | RF | 0.6025 | 57 | uncalibrated |
| FULL | RF | 0.9975 | 57 | isotonic_crossfit |
| FULL | RF | 0.9988 | 57 | platt_crossfit |
| FULL | RF | 0.9988 | 57 | uncalibrated |
