## Supplementary material for "Artificial Intelligence for Cardiac Biomarkers After Myocardial Infarction: A Systematic Review and a Leakage-Aware Modeling Framework": Table S15

**Table S14.** Locked-model split diagnostics and feature integrity checks

| **Parameter** | **FULL** | **CLINICAL** | **BIOMARKERS** |
| --- | --- | --- | --- |
| n_train | 45 | 45 | 45 |
| n_test | 12 | 12 | 12 |
| n_features_expected | 69 | 58 | 10 |
| n_features_used | 53 | 42 | 10 |
| Dropped (empty/constant) | 16 | 16 | 0 |
| Notes | No RFE; calibration: isotonic | RFE applied, top 10 features; calibration: isotonic | No RFE; calibration: isotonic |
