## Supplementary material for "Artificial Intelligence for Cardiac Biomarkers After Myocardial Infarction: A Systematic Review and a Leakage-Aware Modeling Framework": Table S16

**Table S15.** Locked-model diagnostic test performance

| **Parameter** | **BIOMARKERS** | **CLINICAL** | **FULL** |
| --- | --- | --- | --- |
| ROC AUC (test) | 0.886 | 0.629 | 1 |
| Brier score (test) | 0.105 | 0.234 | 0 |
