## Supplementary material for "Artificial Intelligence for Cardiac Biomarkers After Myocardial Infarction: A Systematic Review and a Leakage-Aware Modeling Framework": Table S17A

**Table S16A.** Workflow-level diagnostic checks using permutation testing and simple models - Permutation diagnostic

| **Model** | **Parameter** | **FULL** | **CLINICAL** | **BIOMARKERS** |
| --- | --- | --- | --- | --- |
| LR_ELNET | Mean (± SD) | 0.501 (± 0.102) | 0.5 (± 0.1) | 0.517 (± 0.103) |
| LR_ELNET | Percentiles (95; 99) | 0.674; 0.708 | 0.695; 0.719 | 0.673; 0.711 |
| LR_ELNET | p-value | 0.02 | 0.49 | 0.02 |
| LR_L1 | Mean (± SD) | 0.503 (± 0.097) | 0.506 (± 0.092) | 0.516 (± 0.107) |
| LR_L1 | Percentiles (95; 99) | 0.687; 0.722 | 0.684; 0.706 | 0.681; 0.727 |
| LR_L1 | p-value | 0.02 | 0.333 | 0.02 |
| RF_SHALLOW | Mean (± SD) | 0.503 (± 0.104) | 0.477 (± 0.127) | 0.524 (± 0.113) |
| RF_SHALLOW | Percentiles (95; 99) | 0.665; 0.719 | 0.696; 0.742 | 0.681; 0.743 |
| RF_SHALLOW | p-value | 0.02 | 0.294 | 0.02 |
| SVM_REG | Mean (± SD) | 0.488 (± 0.112) | 0.504 (± 0.113) | 0.509 (± 0.115) |
| SVM_REG | Percentiles (95; 99) | 0.681; 0.758 | 0.675; 0.703 | 0.688; 0.715 |
| SVM_REG | p-value | 0.02 | 0.255 | 0.02 |
