## Supplementary material for "Artificial Intelligence for Cardiac Biomarkers After Myocardial Infarction: A Systematic Review and a Leakage-Aware Modeling Framework": Table S17B

**Table S16B.** Workflow-level diagnostic checks using permutation testing and simple models - Simplified models

| **Variant** | **Model** | **ROC AUC (mean ± SD)** | **Perfect AUC flag (any fold)** |
| --- | --- | --- | --- |
| BIOMARKERS | RF | 0.95 (± 0.051) | TRUE |
| BIOMARKERS | KNN | 0.909 (± 0.084) |  |
| BIOMARKERS | LR_L1 | 0.85 (± 0.089) |  |
| BIOMARKERS | LR_ELNET | 0.821 (± 0.036) |  |
| BIOMARKERS | LR_L2 | 0.814 (± 0.136) |  |
| CLINICAL | RF | 0.582 (± 0.157) | FALSE |
| CLINICAL | LR_ELNET | 0.506 (± 0.061) |  |
| CLINICAL | KNN | 0.502 (± 0.15) |  |
| CLINICAL | LR_L2 | 0.479 (± 0.083) |  |
| CLINICAL | LR_L1 | 0.473 (± 0.09) |  |
| FULL | LR_L1 | 1 (± 0) |  |
| FULL | LR_ELNET | 1 (± 0) |  |
| FULL | KNN | 1 (± 0) |  |
| FULL | RF | 0.994 (± 0.013) | TRUE |
| FULL | LR_L2 | 0.901 (± 0.13) |  |
